# Social health and change in cognitive capability among older adults: findings from four European longitudinal studies

**DOI:** 10.1101/2022.08.29.22279324

**Authors:** Jane Maddock, Federico Gallo, Frank J Wolters, Jean Stafford, Anna Marseglia, Serhiy Dekhtyar, Marta Lenart-Bugla, Eline Verspoor, Marieke Perry, Suraj Samtani, Myrra Vernooij-Dassen, Karin Wolf-Ostermann, Rene Melis, Henry Brodaty, Mohammad Arfan Ikram, Anna-Karin Welmer, Daniel Davis, George B Ploubidis, Marcus Richards, Praveetha Patalay

## Abstract

**Introduction:** In this study we examine whether social health markers measured at baseline are associated with differences in cognitive capability and in the rate of cognitive decline over an 11-to-18-year period among older adults and compare results across studies.

**Methods:** We applied an integrated data analysis approach to 16,858 participants (mean age 65 years; 56% female) from the National Survey for Health and Development (NSHD), the English Longitudinal Study of Aging (ELSA), the Swedish National Study on Aging and Care in Kungsholmen (SNAC-K), and the Rotterdam Study. We used multilevel models to examine social health in relation to cognitive capability and the rate of cognitive decline.

**Results:** Pooled estimates show distinct relationships between markers of social health and cognitive domains e.g., a large network size (≥6 people vs none) was associated with higher executive function (0.17 SD[95%CI:0.0, 0.34], I^2^=27%) but not with memory (0.08 SD[95%CI: -0.02, 0.18], I^2^=19%). We also observed pooled associations between being married or cohabiting, having a large network size and participating in social activities with slower decline in cognitive capability, however estimates were close to zero e.g., 0.01SD/year [95%CI: 0.01 to 0.02] I^2^=19% for marital status and executive function. There were clear study-specific differences: results for average processing speed were the most homogenous and results for average memory were the most heterogenous.

**Conclusion:** Overall, markers of good social health have a positive association with cognitive capability. However, we found differential associations between specific markers of social health and cognitive domains and differences between studies. These findings highlight the importance of examining between study differences and considering context specificity of findings in developing and deploying any interventions.

## Introduction

Cognitive capability, the capacity to undertake the mental tasks of daily living, is an important aspect of healthy ageing [1–3]. Numerous factors across the life course can affect cognitive capability and decline [4–7]. Most research has focused on biological and medical correlates [8]. Understanding if and how an individual’s social connections can affect cognitive capability may provide cost-effective complementary targets to support interventions to delay cognitive decline.

Terms such as social connection, social network, social functioning, and social engagement are used interchangeably. Our paper capitalises on recent conceptual advancement in this area that uses the term social health for clarity [9]. Social health is a concept covering a range of individual and environmental social markers such as social participation and appraisal of social environment [9]. Researchers have theorised that markers of social health can influence cognition through a combination of physiological, psychological, or behavioural pathways. For example, participating in social activities involves cognitive and mental stimulation [10]. Positive aspects of social support may act as a buffer against stress, which itself can impair cognition [11, 12]. Further, health-related behaviours can be influenced by the normative behaviours of a social network, a phenomenon known as the social control theory [12].

There is empirical evidence that markers of social health contribute to cognitive capability, the rate of cognitive decline and risk of dementia [13–18]. A systematic review highlighting the complexities of associations between social health and cognition [15] reported that social activity, larger social networks, and greater social support were associated with greater global cognitive capability, with mixed findings for domain-specific cognitive abilities and less evidence for associations with cognitive decline. Further systematic reviews and meta-analyses focusing on cognitive decline found that poor social relationships (for example, small social network size, low social engagement, and loneliness) were associated with faster rates of decline, but there was heterogeneity between the studies and evidence for publication bias [17, 18].

It is difficult to ascertain the exact nature of the association between social health and cognitive capability for several reasons. First, the term social health refers to a multitude of markers that have been variously defined, operationalised, assessed, and analysed. Second, cognitive capability has often been examined as a composite measure in relation to markers of social health, and differential effects between specific cognitive domains (e.g., memory, processing speed) observed in some previous studies are worth exploring [19, 15]. Third, results from previous meta-analyses may have overlooked important between study differences by combining results from different types of studies that measured and analysed data in different ways.

The current study sought to overcome some of these challenges. We used a clear framework to operationalise markers of social health and examined both specific cognitive domains (memory, executive function, and processing speed) and global or composite cognitive function where possible. Importantly, we applied integrative data analyses across four large European longitudinal studies. Integrated data analysis involves coordination of measurement and analytical protocols between independent studies which maximises comparability between results, while accounting for study specific strengths and design features [20]. The studies and countries chosen are part of the SHARED consortium (https://www.shared-dementia.eu/) which were sampled from the general population, contained the necessary exposure and outcome measures and multiple timepoints required to examine decline, and were homogeneous enough to pool findings and examine replication. This paper further builds on a recent study that examined a similar question in different studies involving fewer timepoints and greater heterogeneity of populations sampled from the COSMIC consortium [21].

Our overall aim was to examine whether social health markers measured at baseline are associated with differences in cognitive capability and in the rate of cognitive decline over an 11-to-18-year period among older adults and compare results across studies.

## Materials and Methods

### Participants

Participants in this paper came from four European population-based longitudinal studies located in the UK, Sweden and the Netherlands: The National Survey of Health and Development (NSHD) [22] and The English Longitudinal Study of Ageing (ELSA) [23] in the UK, The Swedish National Study on Aging and Care-Kungsholmen (SNAC-K) [24] in Sweden and The Rotterdam Study [25] in the Netherlands. Detailed information about these studies is provided in Supplementary File 1. Briefly, NSHD is a British birth cohort where participants of the same age have been followed up 24 times since birth in 1946. Cognitive capability was measured in 1999, 2006-2010, and 2014-2015 when participants were aged 53, 60-64, and 68-69 years. ELSA is a study of adults aged 50 years and over living in private households in England. The original sample was contacted in 2002-2003 and participants are followed up every 2 years. Cognitive capability was measured in waves 1 to 9. SNAC-K is a study of people aged 60 years and older living at home or in nursing homes in Kungsholmen (central Stockholm). Baseline assessment took place between 2001 and 2004. Those aged between 60 and 72 years were followed every six years, and those aged 78 years and older every three years, up until a maximum follow-up period of 12 years. The Rotterdam Study is a study of adults aged 45 years or older that began in 1990-1993. Participants are followed up every four years. For each of these studies, our analytical sample consisted of participants with information on at least one social health variable at baseline, and cognitive capability from at least two time points in one domain. Participants with dementia at baseline and those with missing covariate data (<9%) were excluded (see Supplementary File 1). Table 1 outlines the analytical sample size for each study.

**Table 1.** Descriptive statistics across studies

|  | **NSHD** | **ELSA** | **SNAC-K** | **Rotterdam** |
| --- | --- | --- | --- | --- |
| **N** | 2,109 | 8,460 | 1,892 | 4,397 |
| **Total follow-up time, years** | 16 | 18 | 12 | 11 |
| **Age at baseline, years (SD)** | 53.4 (0.17) | 64.3 (9.8) | 71.4 (9.6) | 72.2 (7.3) |
| **Demographics** |  |  |  |  |
| **Female, %(n)** | 53 (1,116) | 55 (4,625) | 62 (1,182) | 56 (2,466) |
| **Occupational social class, %(n)** |  |  |  |  |
| *Manual* | 33 (699) | 44 (3,732) | 15 (279) | 27 (1193) |
| *Non-manual* | 67 (1,410) | 56 (4,728) | 85 (1613) | 73 (3204) |
| **Education, %(n)** |  |  |  |  |
| *Lower* | 39 (820) | 47 (4,018) | 12 (222) | 11 (484) |
| *Secondary* | 50 (1,053) | 28 (2,390) | 49 (928) | 43 (1,909) |
| *Higher* | 11 (236) | 24 (2,052) | 39 (742) | 46 (2,004) |
| **IADL, %(n)** |  |  |  |  |
| *None* | N/A | 93 (7,841) | 91 (1722) | 55 (2,436) |
| *At least one* |  | 7 (619) | 9 (170) | 45 (1,961) |
| **Vascular-related health conditions, %(n)** |  |  |  |  |
| *None* | 96 (2,029) | 87 (7,334) | 74 (1397) | 67 (2,954) |
| *At least one* | 4 (80) | 13 (1,126) | 26 (495) | 33 (1,443) |
| **Structural social health variables** |  |  |  |  |
| **Marital/cohabitation status, %(n)** |  |  |  |  |
| *Unmarried and alone* | 9 (189) | 24 (2,011) | 48 (907) | 31 (1,386) |
| *Married or cohabiting* | 91 (1,920) | 76 (6,449) | 52 (983) | 68 (3,010) |
| *Missing* | 0 | 0 | 0.001(2) | 0.02 (1) |
| **Network size, %(n)** |  |  |  |  |
| *None* | 4 (78) | 2 (145) | 1 (21) | N/A |
| *1-2 people* | 13 (269) | 12 (975) | 14 (273) |  |
| *3-6* | 30 (626) | 39 (3,330) | 42 (799) |  |
| *≥6* | 54 (1,136) | 40 (3,443) | 37 (690) |  |
| *Missing* | 0 | 7 (567) | 6 (109) |  |
| **Contact frequency, %(n)** |  |  |  |  |
| *Never or almost never* | 0.9 (18) | 1 (49) | 0.6 (11) | N/A |
| *More than once a year* | 8 (161) | 15 (1,271) | 10 (181) |  |
| *About once to twice a month* | 12 (261) | 55 (4,618) | 45 (853) |  |
| *Weekly or more than twice a month* | 48 (1,013) | 20 (1,722) | 36 (682) |  |
| *At least two to three times per week* | 29 (620) | 4 (311) | 5 (97) |  |
| *Missing* | 2 (36) | 6 (489) | 4 (68) |  |
| Functional social health variables |  |  |  |  |
| **Participation in social activities, %(n)** |  |  |  |  |
| *Low ($\leq 1$ activity)* | N/A | 21 (1,751) | 3 (56) | N/A |
| *moderate (2-3 activities)* |  | 33.3 (2,816) | 9 (160) |  |
| *high ($\geq 4$ or more activities)* |  | 46 (3,893) | 82 (1557) |  |
| *Missing* |  | 0 | 6 (119) |  |
| **Positive support\*** |  |  |  |  |
| *Range* | 0-9 | 0-9 | 0-10 | 0-10 |
| *Median (IQR range)* | 6.0 (5.0 – 8.0) | 7.0 (5.8 – 8.0) | 10 (9 – 10) | 10 (9 – 10) |
| *Missing, %(n)* | 3 (70) | 6 (516) | 4 (91) | 0.1 (5) |
| **Negative support\*** |  |  |  |  |
| *Range* | 0-9 | 0-9 | N/A | N/A |
| *Median (IQR range)* | 8.0 (6.0 – 8.0) | 7.3 (6.3-8.0) |  |  |
| *Missing, %(n)* | 3 (70) | 6 (479) |  |  |
| Cognitive capability |  |  |  |  |
| **Memory\*** |  |  |  |  |
| *Range* | 0-15 | 0-10 | 0-16 | 0-15 |
| *At baseline, mean (SD)* | 6.0 (2.0) | 5.6 (1.7) | 7.3 (2.3) | 6.8 (2.0) |
| *\*\*Rate of decline, $\beta$ (95% CI)* | -0.025 (-0.028, -0.022) | -0.03 (-0.04, 0.03) | -0.049 (-0.054, -0.045) | -0.007 (-0.011, -0.003) |
| **Executive function\*** |  |  |  |  |
| *Range* | N/A | 0-50 | 2-50 | 2-47 |
| *At baseline, mean (SD)* |  | 19.9 (6.2) | 22.0 (6.3) | 20.6 (5.3) |
| *\*\*Rate of decline, $\beta$ (95% CI)* |  | -0.01 (-0.02, -0.01) | -0.046 (-0.050, -0.042) | -0.021 (-0.025, -0.017) |
| **Processing speed\*** |  |  |  |  |
| *Range* | 0-780 | 0-780 | 2-34 | 1-55 |
| *At baseline, mean (SD)* | 283.9 (73.9) | 309.7 (94.3) | 18.0 (4.1) | 26.8 (7.0) |
| *\*\*Rate of decline, $\beta$ (95% CI)* | -0.020 (-0.023, -0.017) | -0.03 (-0.04, -0.03) | -0.045 (-0.049, -0.041) | -0.056 (-0.059, -0.053) |

Full details on how each variable was coded are in Supplementary File 1.
\*Raw scores presented for descriptive purposes; standardised scores used in analyses. For negative support, higher scores indicate less feelings of negative support.
\*\*Based on standardised outcomes from mixed-effects multilevel models e.g., -0.03 indicates a 0.03 SD lower cognitive score per year

### Measures

We briefly describe variables used in analyses in the following sections. We harmonised variables across studies as much as possible. Full details of the questions and coding used within each study are in Supplementary File 1. Note that some studies did not include every outcome and exposure of interest (Table 1).

### Social health: structural and functional markers

We distinguish between structural and functional social health markers based primarily on work from Vernooij-Dassen and colleagues [9], complemented by the social networks model by Berkman, and social health domains identified by Huber [26, 27]. Structural characteristics represent the structure of the social network itself in determining opportunities by which the social network can influence health (e.g., marital status, social network size, and frequency of social contact). Functional characteristics denote the role of the social network structure (e.g., social engagement and social support). Structural social health markers at baseline included: marital or cohabitation status (married or cohabiting vs. unmarried and alone); social network size (1-2 people, 3-6 people or ≥6 people vs. none); contact frequency (more than once a year, about once to twice a month, weekly or more than twice a month, at least two to three times per week vs. never or almost never). Functional social relationship variables included: participation in social activities (moderate or high vs. low); perceived positive support (coded as a standardised score and categorised as -1SD to 0SD, 0 to 1SD and >1SD vs. <-1SD); and perceived negative support (coded as a standardised score and categorized as -1SD to 0SD, 0 to 1SD and >1SD vs. <-1SD). We categorised the standardised scores for support as positive support in the Rotterdam Study and SNAC-K had a skewed distribution (Table 1).

### Cognitive capabilities

We included three tests of cognitive capability that were similarly measured across studies. Memory was assessed in all cohorts using an immediate word list recall. Executive function was assessed in ELSA, SNAC-K and the Rotterdam Study using a test of semantic verbal fluency. Processing speed was assessed in NSHD and ELSA using a letter cancellation task, a digit cancellation task in SNAC-K and letter-digit substitute task in RS. We standardised each test across time-points and within studies on a common standard deviation (SD)-based scale. This enhances comparability of estimates between studies while allowing examination of changes over time within studies. Total follow-up time in each study ranged between 11 to 18 years (Supplementary File 1 provides detailed follow-up time for each cognitive outcome within each study). Study-specific global or composite cognitive scores were constructed for sensitivity analyses (full details in Supplementary File 1).

### Confounders

We defined baseline confounders *a priori*: sex (female vs. male); social class (non-manual vs. manual occupation); education (secondary or higher vs. lower); impairment in Instrumental Activities of Daily Living (IADL; at least one vs. none); and vascular-related health conditions excluding hypertension, e.g., diabetes, stroke, heart disease. Mental health may be considered a mediator or confounder, therefore we have not included it in our main models. However, we adjusted for standardised mental health scores in sensitivity analyses. For details of mental health measurement in each study, see Supplementary File 1.

### Analyses

We conducted coordinated analyses for this paper. Following harmonisation of all variables after agreement between authors (Supplementary File 1), JM wrote an exemplar script using Stata 17 and an analyst within each country (JM, FG and FJW) adapted and ran the script for their study.

Within each study, change in each cognitive domain and each individual social health marker were modelled using linear mixed-effects models with random intercepts and slopes and time centered on baseline date and coded as years. This method accounts for within-person correlation between repeated cognitive scores over time. We used two main approaches when applying these linear mixed-effects models. First, we included the social health marker and time in the model and interpreted the coefficient for social health as the association with levels of cognitive capability on average over time, hereinafter referred to as ‘average cognitive capability’. Second, we included an interaction term between the social health marker and time and interpreted the coefficient as association with rate of cognitive decline.

We applied three levels of adjustment to the specified linear mixed-effects models. After estimating associations adjusted for sex and age at baseline, we adjusted first for socio-demographic information (social class and education) and second for health-related information at baseline (IADL and vascular-related health conditions). We decided to use three levels of adjustment as the identity of key confounding variables remains uncertain. We also wanted to explicitly demonstrate the effect of health-related information above socio-demographic information on the observed associations.

It is plausible that associations between social health and cognitive capability might vary by sex [28, 29], therefore each model was repeated stratifying by sex.

Within-study analyses were conducted using Stata. Overall and stratified results from each study were sent to JM who pooled the estimates using random effects meta-analysis with restricted maximum likelihood (*meta* command in Stata 17). Tests of sex-differences were performed using the subgroup option in the *meta* command in Stata 17. We report heterogeneity using the *I*^*2*^ statistic, where 0% indicates estimates were similar across studies and values closer to 100% represent greater heterogeneity. Figures 1 and 3 were created using R.

**Fig. 1.**
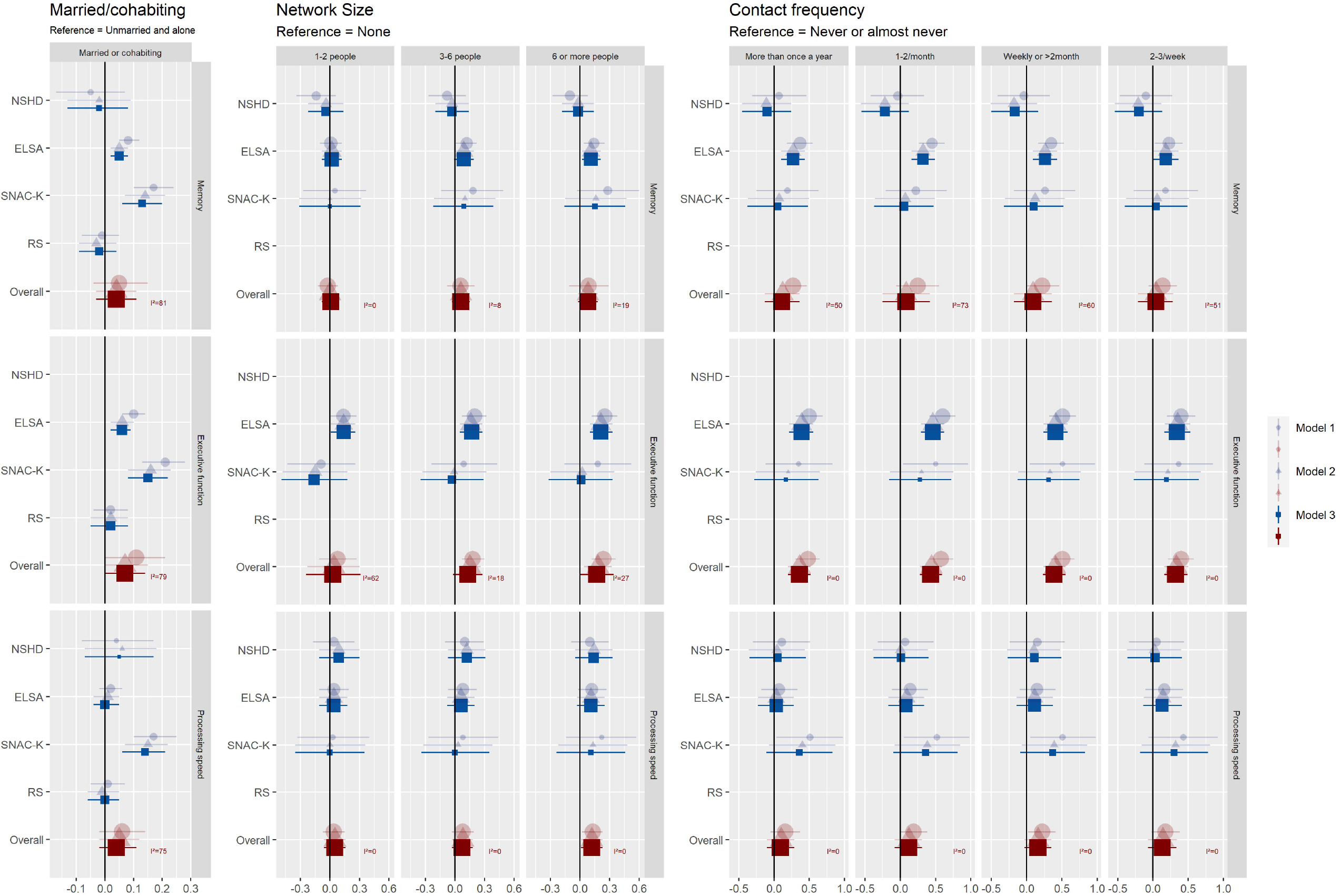
Associations between baseline structural social relationship variables and standardised cognitive capability on average over time. Size of the estimate markers represent study weight. See Supplementary File 2 for this information in table format. Model 1: sex, age at baseline. Model 2: sex, age at baseline, social class, education. Model 3: sex, age at baseline, social class, education, IADL, and vascular-related health conditions.

#### Sensitivity analyses

In sensitivity analyses we examined if associations were robust to additional adjustment for depressive symptoms. We also repeated analyses with a global or composite measure of cognitive function.

## Results

Our analyses included 16,858 participants (mean age 65.3 years; 56% female) with follow-up times ranging from 11 to 18 years across four studies (Table 1). Most participants came from a non-manual social class, particularly in SNAC-K (85%). The proportion of participants who were married or cohabiting was somewhat higher in the two UK studies (NSHD and ELSA) than in SNAC-K and Rotterdam Study, which included on average older participants. Most people had extensive social networks (>37% had more than six people in their network) and reported having high positive support (median ranged from 6 to 10 across studies where scales ranged from 0-9 and 0-10). A large proportion of SNAC-K participants (82%) had high levels of social participation. Markers of social health were weakly correlated within studies (*ρ*<0.03, Supplementary File 2). Cognitive domains were moderately correlated within studies (*r*>0.3) except for memory and processing speed in NSHD, ELSA and SNAC-K (*r*<0.3, Supplementary File 2). We observed decline in all cognitive domains across all studies ranging from -0.001 to -0.06 SD per year (Table 1).

Results for the linear mixed-effects models are discussed in the following sections. We organise results by cognitive domain. Within each subsection we report results first for structural social health markers and then functional social health markers. We focus on the fully adjusted models as we did not observe large differences in results between different adjustment levels (Fig. 1, Fig 3. and Supplementary File 2). We did not observe any statistically significant sex-differences, however when stratifying by sex, estimates tended to be stronger for married males than for married females (Supplementary File 2).

### Memory

#### Structural social health markers

Being ***married or cohabiting*** was associated with 0.05 (95% CI: 0.02, 0.08) and 0.13 (95%CI: 0.06, 0.2) SD higher average memory scores in ELSA and SNAC-K respectively. As seen in Fig. 1. and supplementary file 2, estimates were opposite in sign and confidence intervals crossed the null in NSHD and RS, and a relationship was not observed in the pooled results (0.04 [95%CI: -0.03, 0.11]). This heterogeneity was reflected in the high I^2^ statistic (81%). We observed a similar degree of heterogeneity (I^2^ =76%) when examining how being married or cohabiting associated with the rate of decline in memory. As shown in Fig. 2 and supplementary file 2, clear associations were observed in ELSA and SNAC-K where being married or cohabiting at baseline was associated with a slower rate of decline in memory, although the effect size was small (e.g., 0.02SD/year 95%CI: 0.01 to 0.02 in ELSA).

**Fig. 2.**
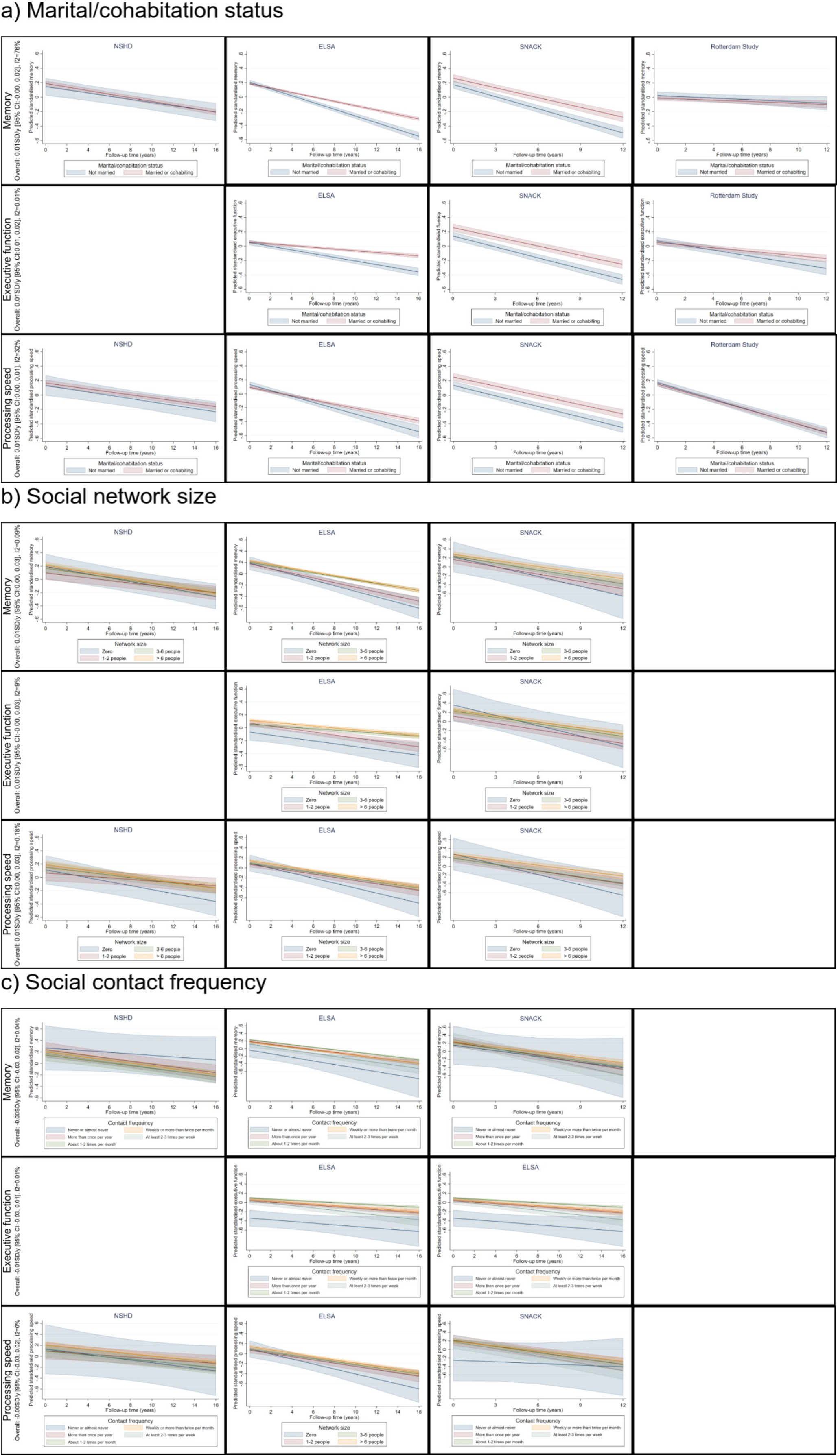
Association between a) marital and cohabitation status b) social network size c) social contact frequency and decline in cognitive function in each study. Estimates are adjusted for sex, age at baseline, social class, education, IADLs, vascular-related health conditions. Overall result reflects the meta-analysed value for decline. Where there is more than one exposure category, the overall value reflects the result from the final category.

There were no clear associations between having a large ***network size*** and average memory scores In NSHD or SNAC-K (shown in Fig. 1 and supplementary file 2). In ELSA, only a very large network size (≥6 people compared with none) was associated with 0.11 (95% CI: 0.02, 0.21) SD higher memory score. RS did not have information about network size. Overall, results across studies were less heterogeneous than for marital or cohabitation status (I^2^ =≤19%) with effect sizes being similar in ELSA and SNAC-K and opposite in sign in NSHD as shown in fig 1. and supplementary file 2. As shown in Fig 2. and supplementary file 2 results were also relatively homogeneous across studies when examining the rate of decline in memory (I^2^ =≤0.09%). Pooled results from NSHD, ELSA and SNAC-K suggest that having a larger network size was associated with a slower rate of decline in memory (e.g., 0.02 SD/year [95%I: 0.01, 0.03] for 3-6 people compared with none).

Overall pooled estimates suggest no association between ***frequent social contact*** and average memory however, results were heterogenous (I^2^ = ≥50%). In ELSA, having frequent social contact was associated with higher memory scores (Fig 1., supplementary file 2). Estimates in SNAC-K were smaller and confidence intervals crossed the null and were opposite in sign in NSHD. RS did not have information about social contact frequency. Results from analyses examining decline in memory were less heterogenous (I^2^ =≤0.08%, Fig 2. supplementary file 2). Overall, there was no evidence that frequency of social contact was associated with the rate of decline in memory.

#### Functional social health markers

Among the two studies with relevant information (ELSA and SNAC-K), ***participation in social activities*** was associated with higher average memory scores as shown in Fig 3 and supplementary file 2. Effect sizes were similar across the two studies (I^2^ = ≤8%) with pooled estimates indicating that participating in ≥4 social activities was associated with 0.32 (95%CI 0.28, 0.35) SD higher average memory score compared with not participating in any social activities. Effect sizes were also similar (I^2^ = ≤0.09%) across the studies when examining rate of decline in memory as shown in Fig. 4 and supplementary file 2. Pooled estimates suggest a slower rate of decline in memory for those participating in ≥4 social activities compared with none (0.01 SD/year 95%CI: 0.01, 0.02, I^2^ =0%).

**Fig. 3.**
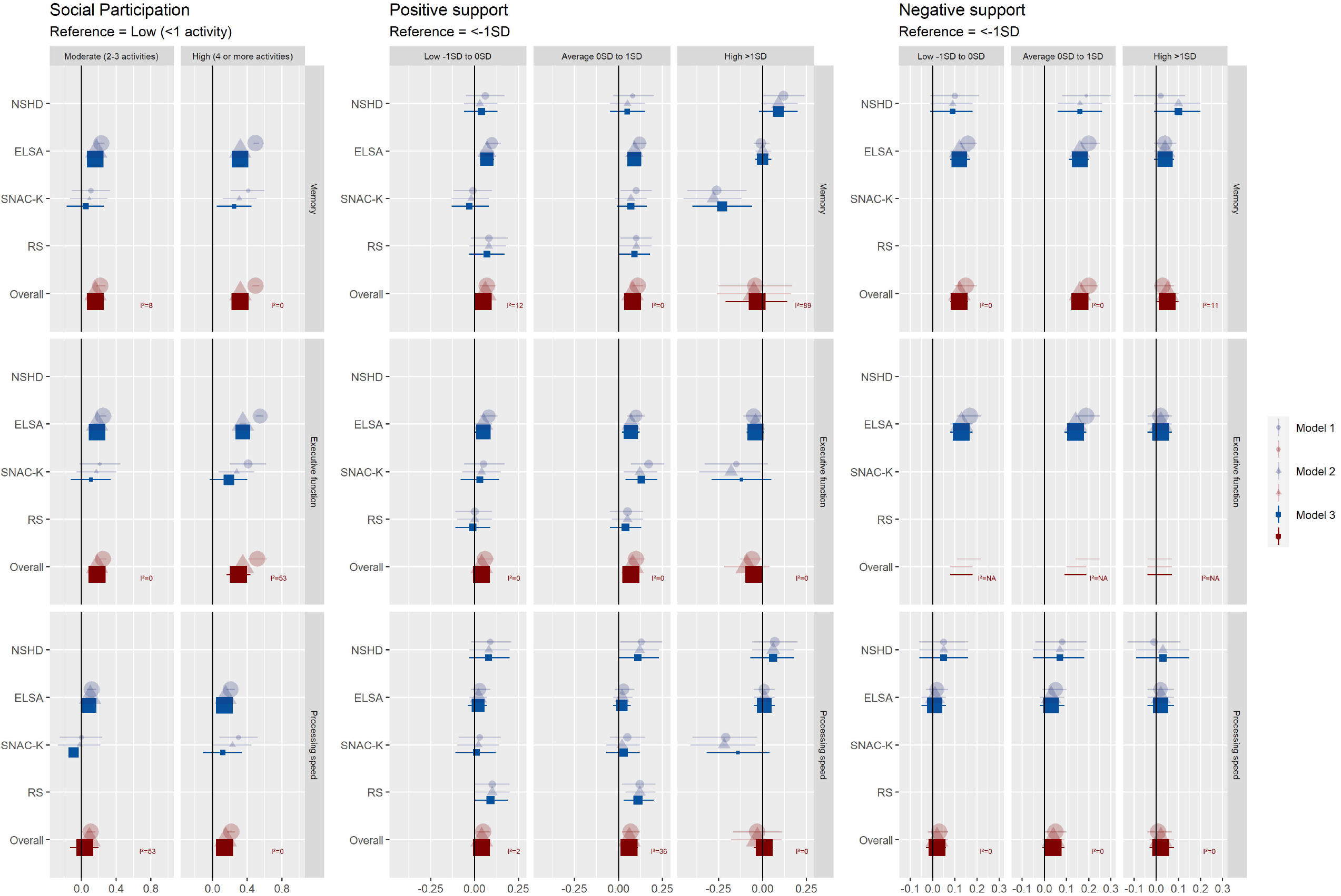
Associations between baseline functional social relationship variables and standardised cognitive capability on average over time. Size of the estimate markers represent study weight. See Supplementary File 2 for this information in table format. Model 1: sex, age at baseline. Model 2: sex, age at baseline, social class, education. Model 3: sex, age at baseline, social class, education, IADL, and vascular-related health conditions.

**Fig. 4.**
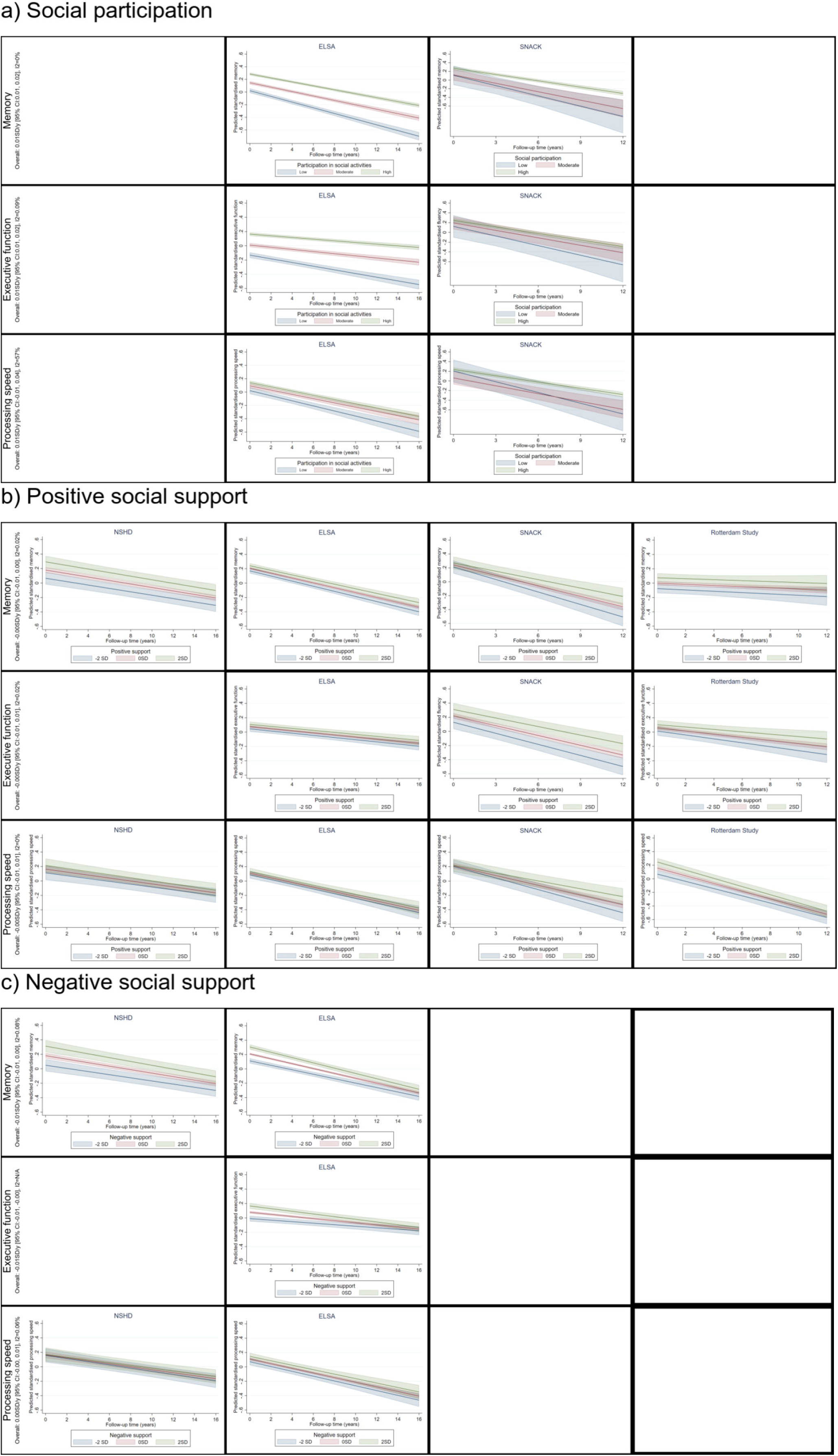
Association between a) social participation b) positive social support c) negative social support and decline in cognitive function in each study. Estimates are adjusted for sex, age at baseline, social class, education, IADLs, vascular-related health conditions. Overall result reflects the meta-analysed value for decline. Where there is more than one exposure category, the overall value reflects the result from the final category.

Pooled estimates in Fig. 3 and supplementary file 2, show that compared with participants who reported having low levels of ***positive social support*** (i.e., <-1 SD) those in the middle of the distribution (between -1 SD to 0SD and 0SD to 1SD) had higher average memory. Estimates were relatively consistent across all studies (I^2^ =≤0.02%). However, results were more heterogeneous (I^2^ =89%) when comparing individuals reporting high levels of positive support (>1SD vs. <-1SD). Participants from RS did not contribute to this due to the distribution of positive social support in the sample. Unexpectedly, estimates were smaller compared with people in the middle of the distribution for ELSA and SNAC-K. There was no strong evidence for an association between positive social support and rate of memory decline across studies (I^2^ =<0.02%, Fig. 4, supplementary file 2).

Only the UK studies (ELSA and NSHD) had information about ***perceived negative aspects*** of social support. Overall pooled results in Fig. 3 and supplementary file 2 show that those the middle of the distribution (i.e., -1SD to 0SD and 0SD to 1SD) had 0.12 (95%CI: 0.08, 0.16) and 0.16 (0.11, 0.20) SD higher average memory scores compared with participants reporting high negative social support (i.e., <-1 SD). Estimates attenuated to 0.05 (95%CI: 0.00, 0.10) for those in reporting low levels of negative social support (>1SD vs <-1SD). Estimates were similar between studies (I^2^ <11%). There was no clear evidence for an association between negative social support and rate of memory decline across studies (Fig. 4, supplementary file 2).

### Executive function

NSHD did not have information about executive function over the follow up period.

#### Structural social health markers

Being ***married or cohabiting*** was associated with higher average executive function scores in ELSA and SNAC-K, with the largest estimate found in SNAC-K (0.15 SD [95%CI: 0.08, 0.22]) as seen in Fig 1. and supplementary file 2. The smallest estimate was found in RS where the confidence interval crossed the null (0.02 [95%CI: -0.05, 0.08]). The difference in effect sizes was reflected in the high I^2^ statistic (79%). Being married or cohabiting was also associated with slower rate of decline in executive function (0.01 SD/year [95%CI:0.01, 0.02]) with similar results observed across all studies (I^2^ =0.01%) as shown in Fig 2 and supplementary file 2.

Only ELSA and SNAC-K had information on ***network size*** and executive function. A larger network size was associated with higher average executive function in ELSA but estimates were in the opposite direction and crossed the null for SNAC-K (I^2^ ≥18%, Fig 1 and supplementary file 2). There was little evidence for an association between network size and the rate of decline in executive function with results being slightly more homogenous (I^2^ =≤20.9%) than for average executive function (Fig 2, supplementary file 2).

***Frequent social contact*** was associated with higher average executive function (Fig 1, supplementary file 2). Only ELSA and SNAC-K contributed to these analyses and findings were similar across the studies (I2=0%), although estimates were larger and more precise for ELSA. The largest pooled estimate was observed for participants who had social contact once to twice a month compared with never or almost never (0.43 [95%CI: 0.28, 0.5]). We did not observe associations between frequency of social contact and the rate of decline in executive function (I2=≤0.06%).

#### Functional social health markers

Pooled results showed ***participating in social activities*** was associated with higher average executive function (e.g, 0.30 95%CI: 0.16, 0.44 for ≥4 activities vs none, Fig 3, supplementary file 2). ELSA and SNAC-K contributed to these analyses and results were homogeneous (I^2^ =0%), however estimates from ELSA tended to be larger and more precise. Fig 4 and supplementary File 2 shows that more social participation at baseline slowed the rate of decline in executive function (e.g., 0.01 (95%CI: 0.01, 0.02), I^2^ =0.09%).

Pooled results in Fig 3 showed that compared with those reporting low levels of ***perceived positive social*** support, those in the middle of the distribution (i.e., -1SD to 0SD and 0 SD to 1SD) had higher average executive function (0.04 [95%CI: 0.00 to 0.08] and 0.07 [95%CI: 0.04, 0.11], I^2^ <0.01%). The association was no longer observed when comparing those in the highest end of the distribution (e.g., >1SD) with those in the lowest (<-1SD): -0.05 (95% CI:-0.1, 0.00, I^2^ =0.01%). Similarly, those in the middle of the distribution at baseline had a slower rate of decline in executive function which was not observed among those in the top of the distribution (Fig 4, supplementary file 2).

ELSA was the only study with information on ***perceived negative social support*** and executive function. Results from this study showed that those in the middle of the distribution had higher average executive function which attenuated for those in the top end of the distribution (Fig 3). While there was no evidence for perceive negative social support at baseline and the rate of decline in executive function, those in the top end of the distribution tended to have a faster decline (−0.01SD/year 95%CI: -0.01, - 0.00, Fig 4, supplementary file 2).

### Processing speed

#### Structural social health markers

Being ***married or cohabiting*** was associated with higher average processing speed scores in SNAC-K only (0.14 SD 95%CI:0.06, 0.21, Fig. 1). Other estimates crossed the null and overall, results were heterogenous (I^2^ =75%). Results were also slightly heterogeneous for the rate of decline (I^2^ =32%), with ELSA showing the clearest associations between being married or cohabiting at baseline and 0.01SD/year (95%CI: 0.00, 0.02) slower rate of decline in processing speed with small estimates from other studies (Fig. 2, supplementary file 2).

There was some evidence for pooled associations between having large ***network size (i.e***., ***≥6 people)*** and average processing speed (Fig. 1, I^2^ =<0.01%). Confidence intervals from individual studies examining network size and decline in processing speed cross the cross the null, however, pooled analyses found having a larger network size was associated with slower rate of decline in processing speed (Fig 2. e.g, 0.01 SD/year [95%CI: 0.00, 0.03] for ≥6 people vs. none, I =0.18%)

There was no evidence for associations between ***frequency of social contact*** and average processing speed scores among included studies (Fig 1, I^2^ =0%). Similarly, there was no evidence for frequency of social contact and rate of decline in processing speed (Fig 2, I^2^ =≤0.02%)

#### Functional social health markers

Only ELSA and SNAC-K contributed to analyses examining ***social participation*** and processing speed. In ELSA, high social participation (≥4 activities) was associated with 0.14 SD (95%CI: 0.03, 0.19) higher average processing speed scores. Estimates were smaller and confidence intervals crossed the null for SNAC-K (Fig. 3, Supplementary file 2). We did not observe that social participation at baseline was associated with the rate of decline of processing speed (Fig 4, supplementary file 2), however this was heterogenous for those participating in ≥4 activities with SNAC-K showing a 0.03 SD/year (95%CI: 0.00 to 0.06) slower rate of decline that was not observed in ELSA.

While confidence intervals for estimates in all studies, expect for RS, crossed the null, pooled results indicated that those in the middle of the distribution for ***perceived positive social support*** had higher average processing speed (Fig 3, supplementary file 2, I^2^ <36%)). Like memory and executive function, this attenuated for those in the highest end of the distribution. There was no clear evidence for associations between perceived social support and rate of decline in processing speed, however for those in the middle of the distribution, results were heterogenous (I^2^ ≥59%, Fig 3, Supplementary file 2).

We did not observe evidence for associations between ***perceived negative social support*** and average processing speed which included the two UK studies only (fig 3, supplementary file 2). Those in the middle of the distribution tended to have slower decline in processing speed, but this was not observed for those in the highest (Fig 4, supplementary file 2).

### Sensitivity analyses

Overall, additional adjustment for depressive symptoms slightly attenuated estimates but did not change general interpretation (Supplementary File 2).

Estimates for the global or composite measures of cognitive capability were of a similar magnitude as the domain-specific estimates and showed similarly heterogeneity between studies (Supplementary File 2). Being married or cohabiting was associated with 0.07 SD (95% CI: 0.01 to 0.15, *I*^2^ =81%) higher average composite or global score and slower rate of decline (0.01 SD/year [95% CI: 0.00, 0.02], *I*^2^ =84%) decline per year), with the average association being stronger for males (0.15 SD [95%CI: 0.06 to 0.25]) than for females (0.03 SD [95%CI:-0.05 to 0.11]; *p*-value for subgroup difference=0.05). Having a larger network size (≥6 people vs. none) was associated with a 0.15 SD (95%CI: 0.05 0.24, *I*^2^ =0%) higher average composite or global score and a slower rate of decline (0.02 SD/year [95%CI: 0.01 to 0.03], *I*^2^ =0.02%). We observed no associations between frequency of contact and the composite or global score of cognitive capability. More frequent participation in social activities was associated with 0.37 SD (0.32 to 0.41, *I*^2^ =0.01%) higher average composite score and a slower rate of decline (0.02 SD /year [95%CI: 0.01 to 0.02], *I*^2^ =0.09%). Compared with participants who reported having low levels of *positive* social support or high levels of *negative* support, those in the middle of the distribution (between -1SD to 1SD) had a higher average composite or global score (e.g. 0.09 [95%CI: 0.05 to 1.12, *I*^2^=0.2%] for positive support and 0.14 [95%CI: 0.09 to 0.18, *I*^2^=0.01%] for negative support), but we did not observe associations with high positive support and low negative support, nor with rate of decline for either of the support measures.

## Discussion

We applied coordinated analyses to data from participants aged 50 years and older from four longitudinal European studies and, overall, found that markers of good social health were associated with higher average cognitive capability and slower rate of cognitive decline. However, despite harmonisation of measures and application of the same analytic protocol, associations varied across studies, exposure type and cognitive domains.

When focusing on pooled estimates from our study, findings support conclusions from previous research suggesting distinct relationships between markers of social health and cognitive domains [15, 21]. Findings for structural markers of social health (marital or cohabitation status, social network size, contact frequency) were mixed — all were associated with executive function, and network size was also associated with processing speed. We did not observe associations between structural markers of social health and memory. We did find associations between functional aspects of social health (social participation, positive and negative social support) across all cognitive domains (except for negative social support and processing speed). This result may reflect distinct mechanisms underlying the associations between social health and cognitive capability. Active engagement and quality of interpersonal relationships are key components in our definition of functional aspects of social health. While engagement may be beneficial for cognitive capability through cognitive stimulation [30], positive interpersonal relationships may act as a buffer against stressful life events [11, 12]. This implies that functional aspects of social health may affect different aspects of cognitive capability through several pathways. One specific result requires further discussion. For both positive and negative aspects of social support, we observed non-linear relationships. Compared to those in the lower end of the distribution (<-1SD), those in the middle scored higher on cognitive tests, but this effect attenuated for the higher part of the distribution (>1 SD). There are several potential explanations for this. There may be a threshold for the beneficial effects of social support on cognitive capability. Alternativity, this could reflect reverse causality; participants with lower cognitive capability at baseline may already receive higher social support at baseline.

We also observed pooled associations between marital or cohabitation status, network size and social participation with the rate of decline in cognitive capability, however estimates were close to zero. It is possible that, although the follow-up time in our study ranged from 11 to 18 years, it may take longer for many participants to experience the effect of social health on cognitive decline. Alternatively, certain social health markers may convey a protective effect on the baseline level of cognitive capability, namely by affecting its peak levels during the life course, but less on the rate of its decline. Researchers from the United States observed a similar association between education and initial levels of global cognition but not with the rate of cognitive change among older participants [31].

The goal of pooling estimates from different studies is to provide a unified and precise estimate to support clear conclusions [32]. There are several reasons why estimates may not be the same between studies; variables may be measured differently, statistical analyses might be different, characteristics of study populations may be different and there might be a true difference in the effect of interest between studies. A random-effects meta-analysis aims to incorporate the underlying between-study variation [32]. With the co-ordinated approach taken, we minimised heterogeneity due to measurement and analytical approaches and focused our interpretation on heterogeneity between studies as well as the pooled estimate. Overall, results for average processing speed were the most homogenous and results for average memory were the most heterogenous. We found heterogenous estimates across studies for marital or cohabitation status and all cognitive domains. Being married or cohabiting was associated with higher average cognitive capability in ELSA and SNAC-K. This was not observed in NSHD or RS. Conversely, we observed relatively consistent associations between positive social support and average cognitive capability. Similarly, we observed associations participating in social activities and higher average cognitive capability across all included studies, although this may be because NSHD and RS did not contribute data to these analyses. Results for social health and decline in cognitive capability were relatively homogeneous across studies, except for marital and cohabitation status.

A major strength of our study was the inclusion of four European studies which resulted in a large sample size and the ability to examine replication of findings across datasets. We applied a single framework and coordinated analyses to multiple datasets, selected outcomes that were similarly measured across studies, and harmonised variables as far as possible. This approach has the advantage of reducing conceptual and analytical heterogeneity. Associations that are consistently observed across all studies support the existence of a true effect (or lack thereof). This approach also highlights the importance of not relying on single datasets. Where findings diverge between studies, further exploration of study-specific or context-specific consideration is needed, and caution is required when interpreting the overall pooled effect. However, there are limitations to our study. Assuming that attrition predominantly affected people with poor social health and poor cognition, this could have led to an underestimation of the relationships of interest. Despite our efforts to harmonise measures, some of the heterogeneity in findings between studies observed could be due to differences in the wording of social health questions, and differences in the distribution of social relationship variables. Importantly, while we examine social health markers at baseline and cognitive capability over a long follow-up, a bi-directional association between social health and cognitive capability cannot be ruled out and further studies examining the direction of this relationship are warranted.

In conclusion, we provide evidence that markers of good social health have a positive association with cognitive capability in later life. However, we found differential associations between specific markers of social health and cognitive domains, and not all findings replicated consistently across datasets, highlighting the importance of examining between study differences and considering context specificity of findings in developing and deploying any interventions. Social contact is recognised as a key component in dementia prevention, intervention, and care [33]. Understanding the mechanisms through which markers of social health can affect cognitive capability is a key next step to identify effective policy-level interventions that target social health. As such, our findings may guide future studies to determine if promoting social health at old age may delay cognitive decline.

## Data Availability

NSHD data used in this study are available to bona fide researchers upon request to the NSHD Data Sharing Committee via a standard application procedure. Further details can be found at http://www.nshd.mrc.ac.uk/data doi: 10.5522/NSHD/Q101, 10.5522/NSHD/Q102 and doi: 10.5522/NSHD/Q103. ELSA data used in this study are available to download through the UK data service. doi.org/10.5255/UKDA-SN-5050-16. SNAC-K data used in this study are available to researchers upon approval by the SNAC-K data management and maintenance committee. Applications for accessing these data can be submitted to Maria Wahlberg at the Aging Research Center, Karolinska Institutet, Stockholm, Sweden. Requests for access to the data from Rotterdam Study reported in this paper can be addressed to the data management team of the Rotterdam Study.

## Statements

## Acknowledgment

The contributing studies have been made possible because of the tireless dedication, commitment and enthusiasm of the many people who have taken part. We would like to thank the participants and the numerous team members involved in the studies including interviewers, technicians, researchers, administrators, managers, health professionals and volunteers. We are additionally grateful to our funders for their financial input and support in making this research happen.

## Statement of Ethics

Ethical approval for the MRC National Survey of Health and Development study was obtained from Research Ethics Committees. Written, informed consent was obtained from the study member for each component of data collection. ELSA has received ethical approval from a Research Ethics Committee. SNAC-K was approved by the Ethical Review Board in Stockholm and written informed consent was obtained from participants or their next of kin. The Rotterdam Study has been approved by an Ethics Committee. The Rotterdam Study has been entered into the Netherlands National Trial Register (NTR; www.trialregister.nl) and into the WHO International Clinical Trials Registry Platform (ICTRP; www.who.int/ictrp/network/primary/en/) under shared catalogue number NTR6831. Written informed consent was obtained from all participants.

## Conflict of Interest Statement

Authors report no conflicts of interest.

## Funding Sources

This work was supported by the ‘Social Health And Reserve in the Dementia patient journey (SHARED) SHARED Consortium, an EU Joint Programme-Neurodegenerative Disease Research (JPND). The project is supported by Alzheimer’s Society (Ref:469) in the UK, by ZonMw/JPND (733051082) in the Netherlands, by NCBiR (National Center for Research and Development in Poland, project number (JPND/06/2020) in Poland, by the NHMRC (National Health and Medical Research Council (APP1169489) in Australia. FG received additional support from the Basic Research Program at the National Research University Higher School of Economics. NSHD is funded and supported by the UK Medical Research Council (MC_UU_00019/2). The English Longitudinal Study of Ageing was developed by a team of researchers based at University College London, NatCen Social Research, the Institute for Fiscal Studies, the University of Manchester and the University of East Anglia. The data were collected by NatCen Social Research. The funding is currently provided by the National Institute on Aging (Ref: R01AG017644) and by a consortium of UK government departments: Department for Health and Social Care; Department for Transport; Department for Work and Pensions, which is coordinated by the National Institute for Health Research (NIHR, Ref: 198-1074). Funding has also been provided by the Economic and Social Research Council (ESRC). SNAC-K (http://www.snac.org) is financially supported by the Swedish Ministry of Health and Social Affairs; participating County Councils and Municipalities; the Swedish Research Council; and Swedish Research Council for Health, Working Life and Welfare. This project was funded by the Swedish Research Council for Health, Working Life, and Welfare (FORTE grant no.: 2018-01888 to AK Welmer).

## Author Contributions

JM, PP, AM, SD, AKW conceptualised the study. JM, FG, FJW, AM, SD, AKW were involved in data curation. JM, FG, FJW conducted formal analyses. KWO, RM, HB, MAI, AKW, DD were involved in funding acquisition. JM, PP designed the methodology for this study. KWO and MAI were involved in SHARED project administration. PP supervised this study. JM was responsible for visualization. JM wrote original draft. All authors provided critical review and editing of the original manuscript.

## Data Availability Statement

NSHD data used in this study are available to bona fide researchers upon request to the NSHD Data Sharing Committee via a standard application procedure. Further details can be found at http://www.nshd.mrc.ac.uk/data doi: 10.5522/NSHD/Q101, 10.5522/NSHD/Q102 and doi: 10.5522/NSHD/Q103. ELSA data used in this study are available to download through the UK data service. doi.org/10.5255/UKDA-SN-5050-16. SNAC-K data used in this study are available to researchers upon approval by the SNAC-K data management and maintenance committee. Applications for accessing these data can be submitted to Maria Wahlberg at the Aging Research Center, Karolinska Institutet, Stockholm, Sweden. Requests for access to the Rotterdam Study data reported in this paper can be addressed to the data management team of the Rotterdam Study.

## Notes

### Competing Interest Statement

The authors have declared no competing interest.

### Funding Statement

This project is part of the Social Health And Reserve in the Dementia patient journey (SHARED) SHARED Consortium, an EU Joint Programme-Neurodegenerative Disease Research (JPND). The project is supported by Alzheimer's Society (Ref:469) in the UK, by ZonMw/JPND (733051082) in the Netherlands, by NCBiR (National Center for Research and Development in Poland, project number (JPND/06/2020) in Poland, by the NHMRC (National Health and Medical Research Council (APP1169489) in Australia. FG received additional support from the Basic Research Program at the National Research University Higher School of Economics. NSHD is funded and supported by the UK Medical Research Council (MC_UU_00019/2). The English Longitudinal Study of Ageing was developed by a team of researchers based at University College London, NatCen Social Research, the Institute for Fiscal Studies, the University of Manchester and the University of East Anglia. The data were collected by NatCen Social Research. The funding is currently provided by the National Institute on Aging (Ref: R01AG017644) and by a consortium of UK government departments: Department for Health and Social Care; Department for Transport; Department for Work and Pensions, which is coordinated by the National Institute for Health Research (NIHR, Ref: 198-1074). Funding has also been provided by the Economic and Social Research Council (ESRC). SNAC-K (http://www.snac.org) is financially supported by the Swedish Ministry of Health and Social Affairs; participating County Councils and Municipalities; the Swedish Research Council; and Swedish Research Council for Health, Working Life and Welfare. This project was funded by the Swedish Research Council for Health, Working Life, and Welfare (FORTE grant no.: 2018-01888 to AK Welmer).

### Author Declarations

Ethical approval for the NSHD was obtained from the Greater Manchester Local Research Ethics Committee and the Scotland A Research Ethics Committee. Written, informed consent was obtained from the study member for each component of data collection. ELSA has received ethical approval from the South Central Berkshire Research Ethics Committee (21/SC/0030, 22nd March 2021). SNAC-K was approved by the Regional Ethical Review Board in Stockholm and written informed consent was obtained from participants or their next of kin. The Rotterdam Study has been approved by the Medical Ethics Committee of the Erasmus MC (registration number MEC 02.1015) and by the Dutch Ministry of Health, Welfare and Sport (Population Screening Act WBO, license number 1071272-159521-PG). The Rotterdam Study has been entered into the Netherlands National Trial Register (NTR; www.trialregister.nl) and into the WHO International Clinical Trials Registry Platform (ICTRP; www.who.int/ictrp/network/primary/en/) under shared catalogue number NTR6831. Written informed consent was obtained from all participants.

### Summary of Updates

This version has been revised to correct table 1 and to re-format and elaborate on the results.

