## Supplementary file 1 for "Social health and change in cognitive capability among older adults: findings from four European longitudinal studies"

Supplementary File 1: Study details

Social health and change in cognitive capability among older adults: findings from four European longitudinal studies.

### Table of tables

### Cohort information

MRC National Survey of Health and Development

##### Participants

The Medical Research Council (MRC) National Survey of Health and Development (NSHD) is a British birth cohort. The original NSHD sample consisted of 5,362 singleton babies born in one week in March 1946 to married parents in England, Scotland or Wales, stratified by social class [1]. This sample has been followed up twenty-four times since birth [2, 3]. In 1999 at the 53 year sweep, data were collected from 3,035 participants who compared to the national census data, were somewhat advantaged but still broadly representative of UK-born individuals of the same age [4]. Cognitive capability was measured in 1999, 2006-2010 and 2014-2015 when participants were aged 53, 60-64 and 68-69 years. The analytical sample for this study included 2,981 participants with information on at least one social health variable at baseline and cognitive capability from at least two time points in one domain. Supplementary Table 1 outlines the number of participants with cognitive measures at each time point among analytical sample and Supplementary Table 5 outlines missing data among the analytical sample.

**Supplementary Table 1:** Overview of outcomes in NSHD

|  | **1999**  **(53y)** | **2006-2010**  **(60-64y)** | **2014-2015**  **(68-69y)** |
| --- | --- | --- | --- |
| **Memory** | 2,291 | 2,043 | 1,968 |
| **Processing speed** | 1,998 | 2,071 | 2,308 |

##### Ethics

Ethical approval for the study was obtained from Research Ethics Committees. Written, informed consent was obtained from the study member for each component of data collection.

##### Data access

NSHD data used in this study are available to bona fide researchers upon request to the NSHD Data Sharing Committee via a standard application procedure. Further details can be found at <http://www.nshd.mrc.ac.uk/data> doi: 10.5522/NSHD/Q101, 10.5522/NSHD/Q102 and doi: 10.5522/NSHD/Q103.

##### Study specific statistical analyses

NSHD is a birth cohort, therefore participants were generally the same age when completing cognitive assessments. Age at each wave was used as the time metric and centered to age at baseline.

#### English Longitudinal Study of Ageing

##### Participants

The English Longitudinal Study of Ageing (ELSA) is a study of adults aged ≥50 years living in private households in England [5]. In 2002/2003 the original sample was drawn from households that had previously responded to the Health Survey for England (*n*=11,931). Comparisons of the sociodemographic characteristics of participants against results from the national census indicate that the sample was broadly representative of the English population [5]. Participants are contacted every two years and data collection consists of a face-to-face interview and self-completion questionnaire. The analytical sample for this study included 9,179 core sample members aged ≥50 in wave 1 who had information on at least one social health variable at baseline and cognitive capability from at least two time points in one domain, and who did not have dementia at baseline (*n*=74 with dementia at wave 1). Supplementary Table 2 outlines the number of participants with cognitive measures at each time point among analytical sample and Supplementary Table 5 outlines missing data among the analytical sample.

**Supplementary Table 2**: Overview of outcomes in ELSA

| Wave | 1 | 2 | 3 | 4 | 5 | 6 | 7 | 8 | 9 |
| --- | --- | --- | --- | --- | --- | --- | --- | --- | --- |
| Year | 2002/3 | 2004/5 | 2006/7 | 2008/9 | 2010/11 | 2012/13 | 2014/15 | 2016/17 | 2018/19 |
| Memory | 9,079 | 8,578 | 7,316 | 6,354 | 5,921 | 5,365 | 4,636 | 3,979 | 3,429 |
| Executive function | 9,078 | 8,589 | 7,312 | 6,355 | 5,903 |  | 4,635 | 4,011 | 3,455 |
| Processing speed | 8,831 | 8,327 | 6,960 | 5,685 | 5,477 |  |  |  |  |

##### Ethics

ELSA has received ethical approval from a Research Ethics Committee.

##### Study specific statistical analyses

We used cluster and stratification variables to account for the complex sample design used in ELSA [6]. Non-response weights were also used as outlined in the manuscript. The date of individual interview (month/year) at each wave was used as the time metric. This was centered to date at wave 1 and coded as yearly intervals.

##### Data access

ELSA data used in this study are available to download through the UK data service. doi.org/10.5255/UKDA-SN-5050-16.

#### The Swedish National study on Aging and Care in Kungsholmen

##### Participants

The Swedish National Study on Aging and Care-Kungsholmen (SNAC-K) is an ongoing population-based longitudinal study [7]. Between 2001 and 2004, *n*=5,111 people, aged ≥60 years, living at home or in institutions in Kungsholmen (central Stockholm), were invited to participate in the baseline assessment. Of these 5,111, 262 had no contact information, 200 died before the baseline assessment, and 59 were deaf, moved away, or were not Swedish speakers. Of the 4590 alive and eligible older adults, 1227 declined to participate, leaving a study population of 3363 (participation rate = 73.3%). Of these 3363, *n*=2848 participants completed cognitive tests (n=390 refused to participate in the psychological testing, n=106 had MMSE score<10, n=10 died before the testing, and n=9 were not included for other reasons) [8]. The younger age cohorts (60, 66, and 72 years) were followed every 6 years (2007-2010 and 2013-2016; the 72 age-cohort was further assessed in 2010-2013) and the older age cohorts (≥78 years) every 3 years (2004-2007, 2007-2010, 2010-2013, and 2013-2016). The relevant cognitive function and social health variables to be included in this study were measured at baseline and each follow-up. For the current study, we further excluded participants with dementia (*n*=121), Parkinson’s disease (*n*=19) or schizophrenia (*n*=4) at baseline, as well as those who had cognitive capability information for less than two time points in any one domain (n=749). Thus, 1955 dementia-free participants were included in the analytical sample for this study. Description of study design and data collection protocol are available at https://www.snac-k.se/for-researchers/. Supplementary Table 3 outlines the number of participants with cognitive measures at each time point among analytical sample and Supplementary Table 5 outlines missing data among the analytical sample.

**Supplementary Table 3:** Overview of outcomes in SNAC-K

| Wave | 1 | 2 | 3 | 4 | 5 |
| --- | --- | --- | --- | --- | --- |
| Year | 2001/4 | 2004/7 | 2007/10 | 2010/13 | 2013/16 |
| Memory | 1,934 | 647 | 1,673 | 507 | 1,024 |
| Executive function | 1,955 | 672 | 1,693 | 528 | 1,062 |
| Processing speed | 1,903 | 595 | 1,623 | 479 | 1,008 |

##### Ethics

SNAC-K was approved by the Ethical Review Board in Stockholm and written informed consent was obtained from participants or their next of kin.

##### Data access

SNAC-K data used in this study are available to researchers upon approval by the SNAC-K data management and maintenance committee. Applications for accessing these data can be submitted to Maria Wahlberg at the Aging Research Center, Karolinska Institutet, Stockholm, Sweden.

#### Rotterdam Study

##### Participants

The Rotterdam Study (RS) is a prospective population-based cohort study comprising 14,926 subjects aged 45 years or older [9]. Baseline data from 7,983 participants were collected between 1990 and 1993 (response 78%), with subsequent cohort expansions in 2000 (3,011 individuals, 67%) and 2006 (3,236 individuals, 65%). Participants are interviewed at home and re-examined at a dedicated research centre once every 4 years. In addition, the entire cohort is continuously under surveillance for disease outcomes through linkage of electronic medical records with the study database. For the current study we excluded participants with dementia at baseline (n=37) as well as those who had cognitive capability information for less than two time points in any one domain. The analytical sample for this study was n=5,168. Supplementary Table 4 outlines the number of participants with cognitive measures at each time point among analytical sample and Supplementary Table 5 outlines missing data among the analytical sample.

**Supplementary Table 4:** Overview of outcomes in Rotterdam Study

| Wave | 1 | 2 | 3 |
| --- | --- | --- | --- |
| Year | 2002-2006 | 2009-2014 | 2014-2016 |
| Memory | 4000 | 2405 | 892 |
| Executive function | 4080 | 2666 | 929 |
| Processing speed | 4282 | 2574 | 916 |

##### Ethics

The Rotterdam Study has been approved by an Ethics Committee. The Rotterdam Study has been entered into the Netherlands National Trial Register (NTR; www.trialregister.nl) and into the WHO International Clinical Trials Registry Platform (ICTRP; www.who.int/ictrp/network/primary/en/) under shared catalogue number NTR6831. Written informed consent was obtained from all participants.

##### Data access

Requests for access to the data reported in this paper can be addressed to the data management team of the Rotterdam Study.

**Supplementary Table 5**: Missing data among analytical sample

|  | **NSHD** | **ELSA** | **SNAC-K** | **Rotterdam** |
| --- | --- | --- | --- | --- |
| *Analytical N* | *2318* | *9179* | *1955* | *5168* |
| Marital/cohabitation status | 1 (0.04) | 0 | 2 (0.1) | 102 (2.0) |
| Network size | 1 (0.04) | 702 (7.6) | 112 (5.7) | n/a |
| Contact frequency | 44 (1.9) | 612 (6.7) | 69 (3.5) | n/a |
| Social participation | n/a | 35 (0.38) | 127 (6.5) | n/a |
| Positive support | 82 (3.5) | 587 (6.4) | 94 (4.8) | 111 (2.1) |
| Negative support | 82 (3.5) | 602 (6.6) | n/a | n/a |
| Sex | 0 (0) | 0 (0) | 0 (0) | 0 (0) |
| Social class | 37 (1.6) | 694 (7.6) | 16 (0.8) | 255 (4.9) |
| Education | 125 (5.4) | 10 (0.11) | 0 (0) | 77 (1.5) |
| Instrumental activities of daily living | n/a | 34 (0.37) | 48 (2.5) | 108 (2.1) |
| Vascular-related health conditions | 57 (2.4) | 4 (0.04) | 0 (0) | 445 (8.6) |
| Depressive symptoms | 3 (0.13) | 99 (1.08) | 52 (2.7) | 130 (2.5) |
| Values are n (%) |  |  |  |  |

### Variables included

Variables were recoded to facilitate comparison across cohorts as outlined in supplementary tables 7 to 10.

#### Outcomes

We included three tests of cognitive capability in the main analyses (Supplementary Table 7). Memory was assessed in all cohorts using an immediate word list recall. Executive function was assessed in ELSA, SNAC-K and Rotterdam using a test of semantic verbal fluency. Processing speed was assessed in NSHD and ELSA using a letter cancellation task and in SNAC-K using a digit cancellation task.

In sensitivity analyses we examined measures of composite cognitive function. Supplementary Table 6 outlines how this measure was constructed in each study.

**Supplementary Table 6**: Composite cognitive function

| **Study** | **Details** |
| --- | --- |
| NSHD | Standardised tests for memory and processing speed were summed and re-standardised. |
| ELSA | Standardised tests for memory, verbal fluency and processing speed were summed and re-standardised. This was restricted to the waves that had information on all three cognitive tests (waves 1-5). |
| SNAC-K | A composite index of cognitive performance was computed as the average of z-scores for the domains of memory, processing speed and executive function. |
| RS | Test scores on verbal fluency, delayed recall, Stroop interference, letter-digit substitution, and Purdue pegboard tasks were entered into a principal component analysis, to calculate a standardised compound score (g-factor) that captured 51% of the variance in test performance in the population. |

#### Exposures

To have a consistent conceptualisation of the social health markers across cohorts, we used information from discussion within the Social Health And Reserve in the Dementia patient journey (SHARED) consortium [10] as well as previous literature [11, 12] and applied it to the data available. We distinguish between structural characteristics (e.g., marital status, social network size, and contact frequency) and functional characteristics (e.g., social engagement and social support). All cohorts included in this study have information on at least one social health variable (Supplementary Table 8 and Supplementary Table 9).

#### Covariates

Unless stated, covariates at baseline were included in this study (Supplementary Table 10).

**Supplementary Table 7:** Description of cognitive tests within each study.

| **Study** | **Description of test** | **N follow-up*** | **Follow-up time (years)** |
| --- | --- | --- | --- |
| **Memory (standardized immediate word recall)** | | | |
| NSHD | Participants were shown 15 words and were asked to write down as many as possible from memory, in any order. Different word lists were used to minimise practice effects. This test was conducted three times. Scores from the first test were included in this study [13]. | 3 | 16 |
| ELSA | Participants were presented orally (using a taped voice) with a wordlist containing ten words. They were asked to recall as many words as possible immediately after the reading of the list. Different word lists were used to minimise practice effects. | 9 | 18 |
| SNAC-K | Participants were presented orally and visually with 16 unrelated nouns (5s pace). Immediately after presentation, participants were given 2 minutes to recall the nouns. Number of words correctly remembered was recorded [8]. | 5 | 12 |
| Rotterdam | Participants were shown 15 words and were asked to write down as many as possible from memory, in any order. Number of words correctly remembered was recorded [14]. | 3 | 11 |
| **Executive function (Standardised verbal fluency)** | | | |
| NSHD | NA |  |  |
| ELSA | Participants were asked to name as many different animals as possible in one minute. | 8 | 18 |
| SNAC-K | Participants were asked to name as many different animals as possible in one minute. The final score includes the number of animal identified in one minute [8, 15]. | 5 | 12 |
| Rotterdam | Participants were asked to name as many different animals as possible in one minute [14]. | 3 | 11 |
| **Processing speed (standardized processing score)** | | | |
| NSHD | Letter cancellation is a test of attention, mental speed and visual scanning.  The participant was given a page of random letters of the alphabet, set out in 26 rows and 30 columns, and is asked to cross out as many “Ps” and “Ws” as possible within one minute (65 target letters in total). Respondents were instructed to work across each row from left-to right as if they were reading a page and they were asked to perform the task as quickly and accurately as possible. When the allotted time is over the respondent is asked to underline the last letter that their eye has reached. The total number of letters searched provides a measure of processing speed [13]. | 3 | 16 |
| ELSA | Participants completed the letter cancellation task as outlined in NSHD. | 5 | 10 |
| SNAC-K | Participants are presented with 11 rows of random digits (1-9) and asked to cross out every 4 they encountered as quickly as possible within 30 seconds. The final score includes the number of correct patterns crossed within 30s [8]. | 5 | 12 |
| Rotterdam | Letter digit substitution task. | 3 | 11 |
| *includes baseline assessment | | | |

**Supplementary Table 8:** Structural social health markers.

| **Study** | **Question** | **Response** |
| --- | --- | --- |
| **Married or cohabiting (Unmarried and alone=0; Married or cohabiting=1)** | | |
| NSHD | 1) What is your current marital status?  2) How many people in total live in this household, including yourself (including those temporarily absent)? | 1) single, never married; married; separated; divorced; widowed.  2) 1; 2; 3; 4; 5; 6 or over. |
| ELSA | 1) What is your current legal marital status?  2) Number of people in the household | 1) single, never married; married; remarried; legally separated; divorced; widowed.  2) Numeric response. |
| SNAC-K | 1) What is your current marital status?  2) Who lives with the participant? | 1) married (or equivalent); widow/er also in case of civil/common law union); unmarried; divorced.  2) Single; spouse/common-law partner; daughter; son; grandchild; sibling; sister/brother-in-law. |
| Rotterdam | 1) What is your current marital status?  2) Do you live alone, or do you share your household with someone? | 1) never married; married or living with partner; widower; divorced.  2) Single; with partner; with other(s); with son or daughter. |
| **Network size (None=0; 1-2 people=1; 3-6=2; ≥6=3)** | | |
| NSHD | How many friends/relatives do you see once a month or more? | None;1-2; 3-5; 6-10; More than 10. |
| ELSA | For each of the following: children, other family, friends: how many do you have a close relationship with? | Numeric response (*Created a summed score across relationship type).* |
| SNAC-K | How many people do you feel you know well and can talk to about most things (e.g., relatives, friends, neighbours, and/or colleagues)? | None; 1-2 people; 3 people; 4-6 people; 7-9 people; 10-15 people; 16-30 people; more than 30 people. |
| Rotterdam | Not assessed |  |
| **Contact frequency (Never or almost never=0; More than once a year=1; About once to twice a month=2; Weekly or more than twice a month=3; At least two to three times per week=4; (*rounded to nearest category for summed scores*)** | | |
| NSHD | How often do you regularly visit or are visited by friends and relatives? | Never or almost never; once every few months; about once a month; about once a week; almost daily. |
| ELSA | For each of the following: children, friends, family:  Q1. How often do you meet up with them?  Q2. How often do you speak on the phone with them?  Q3. How often do you write or email them? | Three or more a week; once or twice a week; one or twice a month; every few months; once or twice a year; less than once a year or never.  *Recode to:*  *Less than once a year or never=0; once or twice a year=1; every few months=1; one or twice a month=2; once or twice a week=3; Three or more a week=4.* |
| SNAC-K | For each of the following: parents, children, son or daughter-in-law, grandchildren, siblings, other relative, neighbour, friend:  Q1. How often do you meet them in person?  Q2. How often are you in touch via telephone, letters or email? | Never; less often; quarterly more than once/year; monthly more than six times per year; weekly more than twice per month; daily more than twice per week.  *Recode to:*  *Never or less often=0; quarterly more than once/year=1; monthly more than six times per year=2; weekly more than twice per month=3; daily more than twice per week=4.* |
| Rotterdam | Not assessed |  |

**Supplementary Table 9**: Functional social health markers

| **Study** | **Question** | **Response** |
| --- | --- | --- |
| **Participation in social activities** Low (≤1)=0;moderate (2-3 activities)=1; high (≥4 or more activities)=2. | | |
| NSHD | Not assessed. |  |
| ELSA | 1) Are you a member of the following?  Political party, trade union of environmental group; tenants' or residents group or Neighbourhood Watch; church or other religious group; charitable association; education, arts or music group or evening class; social club; sports group, gym or exercise class, not a member  2) How often do you do any of the following: go to the cinema; eat out of the house; go to an art gallery or museum; go to the theatre, concert or the opera | 1) Yes; no.  2) Twice a month or more; about once a month; every few months; about once or twice a year; less than once a year; never  *Recode question 2 to yes; no.* |
| SNAC-K | Have you participated in any of these 12 months?  Cinema/theatre/concert; sporting events; museum/art exhibit; Go to restaurant/pub/café; bingo; dancing; attend church/revival meeting; participate in study circle or a course; participate in volunteer work; participate in association/club work; travel. | Yes, to the same degree; yes to a higher degree; yes to a lesser degree; no |
| Rotterdam | Not assessed |  |
| **Perceived positive support (Standardised score categorised as <-1SD=0; -1SD to 0SD=1; 0SD to 1SD=2; >1SD=3)** | | |
| NSHD | Based on the person you felt closest to in the last 12 months:  1) How much did you confide in the person you felt closest to?  2) How much did they make you feel good about yourself?  3) How much did you share interests, hobbies and fun with the person you felt closest to? | Not at all; a little; quite a lot; a great deal.  *Sum and standardise scores across three questions* |
| ELSA | For each of the following: spouse/partner, children, immediate family, friends:  1) How much do they really understand the way you feel about things?  2) How much can you rely on them if you have a serious problem?  3) How much can you open up to them if you need to talk about your worries? | A lot; some; a little; not at all.  *Reverse code so not at all=0. Compute mean of each question across relationship type i.e., mean of Q1 for partner, children, family and friend. Then sum across questions and standardise.* |
| SNAC-K | 1) Do you feel that you know one or a few people who could give you proper personal/emotional support to manage the stress and troubles of life?  2) Do you know someone with whom you can be yourself, who accepts you for all your good and bad qualities? | Yes, without a doubt; yes, probably; no, probably not; no, not at all.  *Reverse code so not at all=0. Sum and standardise scores across questions* |
| Rotterdam | 1) I know people whom I can count on always  2) I know people who give me a sense of importance  3) I know people who would help me if needed  4) I know people who give me a sense of importance  5) I know people who accept me the way I am | No; somewhat; yes.  *Sum and standardise scores across* |
| **Perceived negative support (Standardised score categorised as <-1SD=0; -1SD to 0SD =1; 0SD to 1SD=2; >1SD=3. Higher scores indicating less negative social support)** | | |
| NSHD | Based on the person you felt closest to in the last 12 months:  1) How much did they give you worries, problems and stress?  2) How much in the last 12 months did talking to the person you feel closest to make things worse? | Not at all; a little; quite a lot; a great deal.  *Reverse code. Sum and standardise scores across questions.* |
| ELSA | For each of the following: spouse/partner, children, immediate family, friends:  1) How much do they criticise you?  2) How much do they let you down when you are counting on them?  3) How much do they get on your nerves? | A lot; some; a little; not at all.  *Compute mean of each question across relationship type i.e., mean of Q1 for partner, children, family and friend. Then sum across questions and standardise.* |
| SNAC-K | Not assessed |  |
| Rotterdam | Not assessed |  |

**Supplementary Table 10:** Covariates measured at baseline

| **Study** | **Description** | **Response** |
| --- | --- | --- |
| **Social class (manual=0; non-manual=1)** | | |
| NSHD | Social class of head of household at 53 years. | Registrar General classification:  1 Professional; II intermediate; IIIINM skilled non-manual; IIIM skilled manual; IV partly skilled; V unskilled |
| ELSA | Occupational class | Three-class National Statistics – Socioeconomic Classification Scheme:  Managerial and professional occupations; intermediate occupations; semi-routine occupations. |
| SNAC-K | Socioeconomic Index based on type of last/longest-held occupation | Manual; non-manual |
| Rotterdam | Current or last occupation for those who are not working any more. | RIASEC classification, Realistic; non-realistic |
| **Education (Lower (lower than secondary education or no education)=0; Secondary (secondary education or equivalent)=1; Higher (university/other post-secondary)= 2)** | | |
| NSHD | Highest educational attainment up to 26 years | None; vocational; Sub GCE or Burnham C; GCSE O-Level or Burnham C; GCSE A-Level or Burnham B; Burnham A2; 1st Degree; Higher degree; Masters; Higher degree; doctorate |
| ELSA | Highest educational attainment | NVQ4/NVQ5/Degree or equiv; Higher ed below degree; NVQ3/GCE A Level equiv; NVQ2/GCE O Level equiv; NVQ1/CSE other grade equiv; Foreign/other; No qualification |
| SNAC-K | Highest educational attainment | Unfinished primary education; Primary school ("folkskola", ca 6 yrs.); Elementary school/Secondary "realskola"/girls' school; High school/Upper secondary/"Gymnasium"; "Folkhögskola" Folk high school/Vocational/Trade school; Education of at least one year after high school graduation; College/University degree |
| Rotterdam | Highest educational attainment | Primary school only; lower/intermediate general education / lower vocational; intermediate vocational or higher general education, Higher vocational or university. |
| **Instrumental activities of daily living (none=0; at least one=1)** | | |
| NSHD | Not assessed at baseline |  |
| ELSA | Because of physical, mental, emotional or memory problems do you have any difficulty with:  Using a map, preparing a hot meal, shopping for groceries, making telephone calls, taking medications, doing work around the house or garden, managing money. | |
| SNAC-K | Participants were asked whether they could: independently manage their daily activities (e.g. cooking, cleaning, and running errands); buy grocery; prepare a meal; manage household chores (light and heavy); manage laundry; manage household economy; use the telephone; use public transports; drive a car (if participant’s had access to a car). | |
| Rotterdam | Running errands, household chores, doing work around the house or garden, use the telephone, prepare a meal, manage laundry, manage household economy, manage own medication, get in and out of a car, travel independently, cycle. Questions are answered on a 4-point scale (from ‘without difficulty’ to ‘unable to perform by myself’). Items were scored positively if participant indicated that they were unable to perform independently [16]. | |
| **Vascular-related health conditions (0=None; 1=At least one)** | | |
| NSHD | Self-reported diabetes in the last 10 years, stroke in the last 10 years. Rose Angina scale at 53 years. |  |
| ELSA | Self-report doctor diagnosed: angina, heart attack/myocardial infarction, congestive heart failure, heart murmur, abnormal heart rhythm, diabetes, stroke/cerebral vascular disease, other heart trouble. |  |
| SNAC-K | TYPE 2 DIABETES: self-reported medical history, glucose-lowering medication use, medical records from the NPR (ICD-10 code E11), or glycated haemoglobin ≥6.5%.  HEART DISEASES (CVD) including: 1) atrial fibrillation (ICD-10 code I48 and/or discrete P-wave undetectable and irregular ventricular rate), 2) bradycardias and conduction diseases (presence of a cardiac pacemaker and ICD-10 codes I441-I443, I453, I455, Z950), 3) ischemic heart disease (ICD-10 codes I20-I22, I24-I25, Z951,Z955, and/or use of organic nitrates [Anatomical Therapeutic Chemical; ATC, code C01DA] or ranolazine [ATC code C01EB18]), 4) cardiac valve disease (ICD-10 codes I05-I08, I091, I098, I34-I38, I390-I394, Q22-Q23, Z952-Z954), and 5) heart failure (ICD-10 codes I110, I130, I132, I27, I280, I42-I43, I50, I515, I517, I528, Z941, Z943).  CEREBROVASCULAR DISEASES included stroke/TIA and other cerebrovascular syndromes. Diseases were identified according to the ICD-10 codes G45-G46, I60-I64, I67, and I69. |  |
| Rotterdam | Cardiovascular disease includes myocardial infarction, coronary revascularisation procedures, atrial fibrillation, heart failure, and stroke, self-reported with validation in medical records. Diabetes was defined as a fasting serum glucose ≥7.0 or use of antidiabetic medication. |  |
| **Mental Health (Standardised score where higher score indicates more depressive symptoms)** | | |
| NSHD | 28-item general heath questionnaire. | Each item scores 1-4. All items summed and standardised. If an item is missing, impute using individual mean of non-missing items. |
| ELSA | 8-item Center for Epidemiologic Studies Depression Scale. | Yes; no for each item. Reverse code items so that higher score indicated more depressive symptoms. All items summed and standardised. If an item is missing, impute using individual mean of non-missing items. |
| SNAC-K | Montgomery-Åsberg Depression Rating Scale (MADRS) |  |
| Rotterdam | Structured interview to screen for depressive symptoms using the Center for Epidemiologic Studies Depression Scale (CES-D) | 20-item yes/no. Reverse code items so that higher score indicates more depressive symptoms. If an item is missing, impute suing individual mean of non-missing items. |
