## Supplementary File 2 for "Social health and change in cognitive capability among older adults: findings from four European longitudinal studies"

Supplementary File 2: Additional tables and figures

Social health and change in cognitive capability among older adults: findings from four European longitudinal studies.

### Supplementary Table 1 Correlations between markers of social health

|  | Marital/cohabitation status | Network size | Contact frequency | Participation in social activities | Positive support | Negative support |
| --- | --- | --- | --- | --- | --- | --- |
| **NSHD** | | | | | | |
| Marital/cohabitation status | 1 |  |  |  |  |  |
| Network size | -0.04 | 1 |  |  |  |  |
| Participation in social activities | - | - | - | - |  |  |
| Contact frequency | -0.12 | 0.32 | 1 | - |  |  |
| Positive support | 0.09 | 0.08 | 0.06 | - | 1 |  |
| Negative support | -0.02 | 0.00 | 0.03 | - | 0.26 | 1 |
| **ELSA** | | | | | | |
| Marital/cohabitation status | 1 |  |  |  |  |  |
| Network size | 0.03 | 1 |  |  |  |  |
| Contact frequency | -0.07 | 0.17 | 1 |  |  |  |
| Participation in social activities | 0.07 | 0.10 | 0.02 | 1 |  |  |
| Positive support | -0.01 | 0.25 | 0.32 | 0.07 | 1 |  |
| Negative support | -0.15 | 0.10 | 0.04 | 0.00 | 0.29 | 1 |
| **SNAC-K** | | | | | | |
| Marital/cohabitation status | 1 |  |  |  |  |  |
| Network size | 0.15 | 1 |  |  |  |  |
| Contact frequency | -0.10 | 0.13 | 1 |  |  |  |
| Participation in social activities | 0.12 | 0.17 | 0.03 | 1 |  |  |
| Positive support | 0.14 | 0.35 | 0.16 | 0.09 | 1 |  |
| Negative support | - | - | - | - | - | - |
| **RS** | | | | | | |
| Marital/cohabitation status | 1 |  |  |  |  |  |
| Network size | - | - |  |  |  |  |
| Contact frequency | - | - | - |  |  |  |
| Participation in social activities | - | - | - | - |  |  |
| Positive support | 0.05 | - | - | - | 1 |  |
| Negative support | - | - | - | - | - | - |

### Supplementary Table 2: Correlations between cognitive measures at baseline

|  | Memory | Executive Functioning | Processing speed |
| --- | --- | --- | --- |
| **NSHD** | | | |
| Memory | 1 |  |  |
| Executive Functioning | - | - |  |
| Processing speed | 0.11 | - | 1 |
| **ELSA** | | | |
| Memory | 1 |  |  |
| Executive Functioning | 0.38 | 1 |  |
| Processing speed | 0.13 | 0.17 | 1 |
| **SNAC-K** | | | |
| Memory | 1 |  |  |
| Executive Functioning | 0.39 | 1 |  |
| Processing speed | 0.28 | 0.36 | 1 |
| **RS** | | | |
| Memory | 1 |  |  |
| Executive Functioning | 0.40 | 1 |  |
| Processing speed | 0.44 | 0.46 | 1 |

### Supplementary Table 3: Social health at baseline and standardised memory score on average over time

*Adjustment 1= sex, age at baseline*

*Adjustment 2= sex, age at baseline, social class, education.*

*Adjustment 3= sex, age at baseline, social class, education, IADL, and vascular-related health conditions.*

*Blank cells indicate no data available*

|  |  | Adjustment 1 | | Adjustment 2 | | Adjustment 3 | | Additional adjustment for depression | |
| --- | --- | --- | --- | --- | --- | --- | --- | --- | --- |
| Study | Exposure | Coef (95% CI) | Weight/I^2^ | Coef (95% CI) | Weight/I^2^ | Coef (95% CI) | Weight/I^2^ | Coef (95% CI) | Weight/I^2^ |
| **Marital/cohabitation status (reference unmarried and alone)** | | | | | | | | | |
| NSHD | Married or cohabiting | -0.05 (-0.17, 0.07) | 20.16 | -0.02 (-0.13, 0.09) | 19.03 | -0.02 (-0.13, 0.08) | 18.71 | -0.02 (-0.13, 0.08) | 17.10 |
| ELSA | Married or cohabiting | 0.08 (0.05, 0.12) | 28.35 | 0.05 (0.02, 0.08) | 30.14 | 0.05 (0.02, 0.08) | 30.45 | 0.02 (-0.01, 0.05) | 32.56 |
| SNAC-K | Married or cohabiting | 0.17 (0.1, 0.24) | 25.42 | 0.14 (0.07, 0.21) | 24.81 | 0.13 (0.06, 0.2) | 24.80 | 0.11 (0.04, 0.18) | 24.20 |
| RS | Married or cohabiting | -0.01 (-0.08, 0.05) | 26.07 | -0.03 (-0.09, 0.04) | 26.01 | -0.02 (-0.09, 0.04) | 26.04 | -0.03 (-0.09, 0.03) | 26.14 |
| **Overall** | **Married or cohabiting** | **0.05 (-0.04, 0.15)** | **88.51** | **0.04 (-0.03, 0.11)** | **81.76** | **0.04 (-0.03, 0.11)** | **80.67** | **0.02 (-0.04, 0.08)** | **73.46** |
| **Network size (reference: none)** | | | | | | | | | |
| NSHD | 1-2 | -0.14 (-0.34, 0.06) | 24.25 | -0.04 (-0.22, 0.14) | 22.74 | -0.04 (-0.22, 0.14) | 22.97 | -0.04 (-0.22, 0.14) | 23.50 |
| ELSA | 1-2 | 0.01 (-0.1, 0.12) | 65.81 | 0.02 (-0.08, 0.12) | 69.95 | 0.02 (-0.08, 0.12) | 69.60 | 0.02 (-0.09, 0.12) | 69.24 |
| SNAC-K | 1-2 | 0.05 (-0.27, 0.37) | 9.95 | 0 (-0.31, 0.32) | 7.30 | 0 (-0.31, 0.31) | 7.43 | -0.02 (-0.34, 0.3) | 7.25 |
| RS | 1-2 |  |  |  |  |  |  |  |  |
| **Overall** | **1-2** | **-0.02 (-0.12, 0.08)** | **8.95** | **0 (-0.08, 0.09)** | **0.00** | **0.01 (-0.08, 0.09)** | **0.00** | **0 (-0.09, 0.09)** | **0.00** |
| NSHD | 3-6 | -0.08 (-0.27, 0.11) | 32.13 | -0.03 (-0.2, 0.14) | 25.72 | -0.03 (-0.2, 0.14) | 26.13 | -0.04 (-0.2, 0.13) | 26.43 |
| ELSA | 3-6 | 0.12 (0.01, 0.22) | 51.96 | 0.09 (-0.01, 0.19) | 66.33 | 0.09 (-0.01, 0.19) | 65.69 | 0.08 (-0.02, 0.18) | 65.74 |
| SNAC-K | 3-6 | 0.18 (-0.14, 0.49) | 15.91 | 0.1 (-0.21, 0.41) | 7.95 | 0.09 (-0.22, 0.39) | 8.17 | 0.05 (-0.26, 0.36) | 7.83 |
| RS | 3-6 |  |  |  |  |  |  |  |  |
| **Overall** | **3-6** | **0.06 (-0.08, 0.2)** | **44.28** | **0.06 (-0.03, 0.15)** | **7.56** | **0.06 (-0.03, 0.15)** | **8.22** | **0.05 (-0.04, 0.14)** | **7.24** |
| NSHD | ≥6 | -0.1 (-0.28, 0.08) | 34.82 | -0.02 (-0.18, 0.14) | 29.44 | -0.02 (-0.18, 0.14) | 29.65 | -0.03 (-0.19, 0.13) | 28.55 |
| ELSA | ≥6 | 0.14 (0.04, 0.25) | 42.97 | 0.11 (0.02, 0.21) | 61.11 | 0.11 (0.02, 0.21) | 60.75 | 0.09 (-0.01, 0.19) | 63.13 |
| SNAC-K | ≥6 | 0.28 (-0.03, 0.6) | 22.21 | 0.16 (-0.15, 0.47) | 9.45 | 0.15 (-0.16, 0.46) | 9.60 | 0.12 (-0.19, 0.44) | 8.31 |
| RS |  |  |  |  |  |  |  |  |  |
| **Overall** | **≥6** | **0.09 (-0.11, 0.29)** | **71.35** | **0.08 (-0.02, 0.18)** | **19.09** | **0.08 (-0.02, 0.18)** | **19.24** | **0.06 (-0.03, 0.15)** | **12.31** |
| **Contact frequency (reference: never/almost never)** | | | | | | | | | |
| NSHD | >1/year | 0.07 (-0.31, 0.46) | 21.28 | -0.11 (-0.45, 0.24) | 28.67 | -0.1 (-0.45, 0.24) | 28.65 | -0.12 (-0.46, 0.23) | 28.75 |
| ELSA | >1/year | 0.37 (0.18, 0.55) | 61.87 | 0.27 (0.1, 0.44) | 49.66 | 0.27 (0.1, 0.44) | 49.55 | 0.26 (0.09, 0.43) | 49.22 |
| SNAC-K | >1/year | 0.19 (-0.25, 0.63) | 16.86 | 0.07 (-0.37, 0.5) | 21.67 | 0.05 (-0.38, 0.48) | 21.80 | 0.02 (-0.41, 0.45) | 22.03 |
| RS | >1/year |  |  |  |  |  |  |  |  |
| **Overall** | **>1/year** | **0.27 (0.08, 0.47)** | **19.41** | **0.12 (-0.13, 0.36)** | **50.12** | **0.11 (-0.13, 0.36)** | **50.33** | **0.1 (-0.15, 0.35)** | **51.72** |
| NSHD | 1-2/month | -0.04 (-0.42, 0.34) | 29.22 | -0.22 (-0.56, 0.12) | 31.63 | -0.22 (-0.55, 0.12) | 31.59 | -0.22 (-0.56, 0.11) | 31.47 |
| ELSA | 1-2/month | 0.45 (0.26, 0.63) | 45.15 | 0.32 (0.16, 0.49) | 41.68 | 0.32 (0.16, 0.49) | 41.68 | 0.3 (0.14, 0.47) | 41.95 |
| SNAC-K | 1-2/month | 0.22 (-0.21, 0.66) | 25.63 | 0.07 (-0.36, 0.49) | 26.69 | 0.05 (-0.37, 0.47) | 26.73 | 0.02 (-0.4, 0.44) | 26.57 |
| RS | 1-2/month |  |  |  |  |  |  |  |  |
| **Overall** | **1-2/month** | **0.25 (-0.06, 0.55)** | **61.83** | **0.08 (-0.25, 0.42)** | **72.98** | **0.08 (-0.25, 0.42)** | **72.90** | **0.06 (-0.27, 0.39)** | **72.33** |
| NSHD | 2-4/month | -0.04 (-0.41, 0.33) | 26.65 | -0.18 (-0.51, 0.15) | 30.64 | -0.17 (-0.5, 0.16) | 30.56 | -0.18 (-0.51, 0.15) | 30.33 |
| ELSA | 2-4/month | 0.35 (0.16, 0.53) | 51.92 | 0.26 (0.09, 0.43) | 45.76 | 0.26 (0.09, 0.43) | 45.85 | 0.24 (0.07, 0.4) | 46.36 |
| SNAC-K | 2-4/month | 0.26 (-0.18, 0.69) | 21.43 | 0.12 (-0.31, 0.54) | 23.60 | 0.1 (-0.32, 0.52) | 23.59 | 0.07 (-0.35, 0.5) | 23.31 |
| RS | 2-4/month |  |  |  |  |  |  |  |  |
| **Overall** | **2-4/month** | **0.22 (-0.01, 0.46)** | **42.04** | **0.09 (-0.18, 0.36)** | **60.20** | **0.09 (-0.18, 0.36)** | **59.83** | **0.07 (-0.19, 0.34)** | **58.74** |
| NSHD | 2-3/week | -0.1 (-0.47, 0.27) | 23.77 | -0.21 (-0.54, 0.12) | 30.31 | -0.2 (-0.54, 0.13) | 30.20 | -0.22 (-0.55, 0.11) | 30.42 |
| ELSA | 2-3/week | 0.23 (0.03, 0.42) | 59.30 | 0.18 (0, 0.36) | 48.40 | 0.18 (0, 0.36) | 48.54 | 0.17 (0, 0.35) | 47.85 |
| SNAC-K | 2-3/week | 0.18 (-0.27, 0.64) | 16.93 | 0.06 (-0.38, 0.51) | 21.29 | 0.04 (-0.4, 0.49) | 21.26 | 0.01 (-0.43, 0.45) | 21.74 |
| RS | 2-3/week |  |  |  |  |  |  |  |  |
| **Overall** | **2-3/week** | **0.14 (-0.06, 0.34)** | **21.19** | **0.04 (-0.21, 0.29)** | **51.03** | **0.04 (-0.21, 0.28)** | **50.52** | **0.02 (-0.24, 0.27)** | **53.12** |
| **Social participation (reference: low <1 activity)** | | | | | | | | | |
| NSHD | moderate (2-3 activities) |  |  |  |  |  |  |  |  |
| ELSA | moderate (2-3 activities) | 0.23 (0.19, 0.27) | 94.54 | 0.17 (0.13, 0.2) | 97.01 | 0.16 (0.13, 0.2) | 93.48 | 0.14 (0.1, 0.18) | 79.38 |
| SNAC-K | moderate (2-3 activities) | 0.11 (-0.11, 0.33) | 5.46 | 0.09 (-0.13, 0.3) | 2.99 | 0.05 (-0.17, 0.26) | 6.52 | -0.01 (-0.23, 0.22) | 20.62 |
| RS | moderate (2-3 activities) |  |  |  |  |  |  |  |  |
| **Overall** | **moderate (2-3 activities)** | **0.22 (0.17, 0.28)** | **5.14** | **0.16 (0.13, 0.2)** | **0.01** | **0.16 (0.1, 0.21)** | **7.68** | **0.11 (-0.01, 0.23)** | **37.86** |
| NSHD | high (≥4 or more activities) |  |  |  |  |  |  |  |  |
| ELSA | high (≥4 or more activities) | 0.5 (0.47, 0.54) | 96.60 | 0.32 (0.29, 0.36) | 96.32 | 0.32 (0.28, 0.36) | 96.52 | 0.29 (0.25, 0.32) | 96.67 |
| SNAC-K | high (≥4 or more activities) | 0.41 (0.21, 0.6) | 3.40 | 0.31 (0.12, 0.51) | 3.68 | 0.25 (0.05, 0.45) | 3.48 | 0.22 (0.01, 0.42) | 3.33 |
| RS | high (≥4 or more activities) |  |  |  |  |  |  |  |  |
| **Overall** | **high (≥4 or more activities)** | **0.5 (0.46, 0.54)** | **0.00** | **0.32 (0.29, 0.36)** | **0.00** | **0.32 (0.28, 0.35)** | **0.00** | **0.28 (0.25, 0.32)** | **0.00** |
| **Positive social support (reference: low <-1SD)** | | | | | | | | | |
| NSHD | -1SD to 0SD | 0.03 (-0.06, 0.13) | 13.08 | 0.04 (-0.06, 0.13) | 14.49 | 0.03 (-0.07, 0.13) | 11.83 | 0.08 (-0.03, 0.2) | 9.29 |
| ELSA | -1SD to 0SD | 0.07 (0.03, 0.11) | 64.02 | 0.07 (0.03, 0.11) | 59.81 | 0.04 (0, 0.08) | 67.73 | 0.12 (0.08, 0.16) | 63.32 |
| SNAC-K | -1SD to 0SD | -0.02 (-0.13, 0.08) | 11.07 | -0.03 (-0.13, 0.08) | 12.45 | -0.05 (-0.15, 0.06) | 9.74 | 0.1 (0.01, 0.19) | 14.55 |
| RS | -1SD to 0SD | 0.08 (-0.03, 0.18) | 11.83 | 0.07 (-0.03, 0.17) | 13.25 | 0.06 (-0.04, 0.17) | 10.71 | 0.1 (0.01, 0.19) | 12.84 |
| **Overall** | **-1SD to 0SD** | **0.06 (0.02, 0.09)** | **5.28** | **0.05 (0.02, 0.09)** | **11.62** | **0.03 (0, 0.07)** | **12** | **0.11 (0.08, 0.14)** | **0.00** |
| NSHD | 0SD to 1SD | 0.08 (-0.03, 0.2) | 9.29 | 0.05 (-0.05, 0.15) | 10.39 | 0.05 (-0.05, 0.15) | 10.37 | 0.04 (-0.06, 0.14) | 10.59 |
| ELSA | 0SD to 1SD | 0.12 (0.08, 0.16) | 63.32 | 0.09 (0.05, 0.13) | 64.34 | 0.09 (0.05, 0.13) | 64.23 | 0.05 (0.01, 0.09) | 64.24 |
| SNAC-K | 0SD to 1SD | 0.1 (0.01, 0.19) | 14.55 | 0.07 (-0.02, 0.16) | 13.27 | 0.07 (-0.01, 0.16) | 13.35 | 0.05 (-0.04, 0.14) | 12.91 |
| RS | 0SD to 1SD | 0.1 (0.01, 0.19) | 12.84 | 0.1 (0, 0.19) | 12.00 | 0.09 (0, 0.18) | 12.06 | 0.08 (-0.01, 0.17) | 12.26 |
| **Overall** | **0SD to 1SD** | **0.11 (0.08, 0.14)** | **0.00** | **0.08 (0.05, 0.12)** | **0.01** | **0.08 (0.05, 0.11)** | **0.00** | **0.05 (0.02, 0.08)** | **0.00** |
| NSHD | >1 SD | 0.12 (0, 0.24) | 33.24 | 0.09 (-0.01, 0.2) | 33.66 | 0.09 (-0.02, 0.2) | 33.73 | 0.08 (-0.03, 0.18) | 33.75 |
| ELSA | >1 SD | -0.01 (-0.05, 0.04) | 36.75 | 0 (-0.04, 0.05) | 36.31 | 0 (-0.04, 0.05) | 37.76 | -0.03 (-0.08, 0.01) | 40.24 |
| SNAC-K | >1 SD | -0.26 (-0.43, -0.09) | 30.01 | -0.28 (-0.45, -0.12) | 30.03 | -0.23 (-0.4, -0.06) | 28.51 | -0.2 (-0.38, -0.03) | 26.01 |
| RS | >1 SD |  |  |  |  |  |  |  |  |
| **Overall** | **>1 SD** | **-0.04 (-0.25, 0.17)** | **90.88** | **-0.05 (-0.26, 0.16)** | **92.50** | **-0.03 (-0.21, 0.14)** | **88.93** | **-0.04 (-0.18, 0.1)** | **82.96** |
| **Negative social support (reference: low <-1SD)** | | | | | | | | | |
| NSHD | -1SD to 0SD | 0.1 (-0.01, 0.21) | 17.56 | 0.09 (-0.01, 0.18) | 15.43 | 0.09 (-0.01, 0.18) | 15.41 | 0.08 (-0.02, 0.17) | 15.32 |
| ELSA | -1SD to 0SD | 0.16 (0.12, 0.2) | 82.44 | 0.12 (0.08, 0.17) | 84.57 | 0.12 (0.08, 0.17) | 84.59 | 0.09 (0.05, 0.14) | 84.68 |
| SNAC-K | -1SD to 0SD |  |  |  |  |  |  |  |  |
| RS | -1SD to 0SD |  |  |  |  |  |  |  |  |
| **Overall** | **-1SD to 0SD** | **0.15 (0.1, 0.2)** | **9.11** | **0.12 (0.08, 0.16)** | **0.02** | **0.12 (0.08, 0.16)** | **0.01** | **0.09 (0.05, 0.13)** | **0.02** |
| NSHD | 0SD to 1SD | 0.19 (0.08, 0.3) | 13.74 | 0.16 (0.06, 0.26) | 14.82 | 0.16 (0.06, 0.26) | 14.79 | 0.14 (0.04, 0.24) | 14.55 |
| ELSA | 0SD to 1SD | 0.2 (0.16, 0.25) | 86.26 | 0.16 (0.12, 0.2) | 85.18 | 0.16 (0.11, 0.2) | 85.21 | 0.11 (0.07, 0.15) | 85.45 |
| SNAC-K | 0SD to 1SD |  |  |  |  |  |  |  |  |
| RS | 0SD to 1SD |  |  |  |  |  |  |  |  |
| **Overall** | **0SD to 1SD** | **0.2 (0.16, 0.24)** | **0.01** | **0.16 (0.12, 0.2)** | **0.01** | **0.16 (0.12, 0.19)** | **0.02** | **0.12 (0.08, 0.16)** | **0.00** |
| NSHD | >1 SD | 0.02 (-0.1, 0.13) | 16.42 | 0.1 (-0.01, 0.2) | 20.36 | 0.1 (-0.01, 0.2) | 21.24 | 0.07 (-0.03, 0.17) | 29.79 |
| ELSA | >1 SD | 0.04 (-0.01, 0.09) | 83.58 | 0.04 (-0.01, 0.08) | 79.64 | 0.04 (-0.01, 0.08) | 78.76 | 0 (-0.05, 0.04) | 70.21 |
| SNAC-K | >1 SD |  |  |  |  |  |  |  |  |
| RS | >1 SD |  |  |  |  |  |  |  |  |
| **Overall** | **>1 SD** | **0.03 (-0.01, 0.08)** | **0.03** | **0.05 (0, 0.1)** | **8.56** | **0.05 (0, 0.1)** | **11.30** | **0.02 (-0.05, 0.08)** | **38.82** |

### Supplementary Table 4: Social health at baseline and standardised executive function score on average over time

*Adjustment 1= sex, age at baseline*

*Adjustment 2= sex, age at baseline, social class, education.*

*Adjustment 3= sex, age at baseline, social class, education, IADL, and vascular-related health conditions.*

*Blank cells indicate no data available*

|  |  | Adjustment 1 | | Adjustment 2 | | Adjustment 3 | | Additional adjustment for depression | |
| --- | --- | --- | --- | --- | --- | --- | --- | --- | --- |
| Study | Exposure | Coef (95% CI) | Weight/I^2^ | Coef (95% CI) | Weight/I^2^ | Coef (95% CI) | Weight/I^2^ | Coef (95% CI) | Weight/I^2^ |
| **Marital/cohabitation status (reference unmarried and alone)** | | | | | | | | | |
| NSHD | Married or cohabiting |  |  |  |  |  |  |  |  |
| ELSA | Married or cohabiting | 0.1 (0.06, 0.14) | 35.73 | 0.06 (0.02, 0.1) | 37.73 | 0.06 (0.02, 0.09) | 38.24 | 0.03 (0, 0.07) | 39.49 |
| SNAC-K | Married or cohabiting | 0.21 (0.13, 0.28) | 31.38 | 0.16 (0.08, 0.23) | 29.93 | 0.15 (0.08, 0.22) | 29.59 | 0.13 (0.06, 0.21) | 28.56 |
| RS | Married or cohabiting | 0.02 (-0.04, 0.08) | 32.90 | 0.02 (-0.05, 0.08) | 32.33 | 0.02 (-0.05, 0.08) | 32.17 | 0.01 (-0.05, 0.08) | 31.95 |
| **Overall** | **Married or cohabiting** | **0.11 (0, 0.21)** | **89.19** | **0.07 (0, 0.15)** | **81.50** | **0.07 (0, 0.14)** | **79.32** | **0.06 (-0.01, 0.12)** | **74.62** |
| **Network size (references: none)** | | | | | | | | | |
| NSHD | 1-2 |  |  |  |  |  |  |  |  |
| ELSA | 1-2 | 0.14 (0, 0.27) | 76.10 | 0.14 (0.01, 0.26) | 65.33 | 0.14 (0.01, 0.26) | 64.46 | 0.13 (0.01, 0.26) | 68.77 |
| SNAC-K |  | -0.09 (-0.43, 0.26) | 23.90 | -0.15 (-0.48, 0.18) | 34.67 | -0.16 (-0.49, 0.18) | 35.54 | -0.13 (-0.47, 0.21) | 31.23 |
| RS | 1-2 |  |  |  |  |  |  |  |  |
| **Overall** | **1-2** | **0.08 (-0.11, 0.27)** | **29.59** | **0.04 (-0.23, 0.3)** | **59.82** | **0.03 (-0.24, 0.31)** | **62.00** | **0.05 (-0.19, 0.29)** | **50.98** |
| NSHD | 3-6 |  |  |  |  |  |  |  |  |
| ELSA | 3-6 | 0.2 (0.07, 0.32) | 87.47 | 0.16 (0.05, 0.28) | 88.60 | 0.17 (0.05, 0.28) | 81.42 | 0.16 (0.04, 0.27) | 88.74 |
| SNAC-K |  | 0.09 (-0.24, 0.43) | 12.53 | -0.01 (-0.33, 0.32) | 11.40 | -0.03 (-0.35, 0.29) | 18.58 | -0.02 (-0.35, 0.31) | 11.26 |
| RS | 3-6 |  |  |  |  |  |  |  |  |
| **Overall** | **3-6** | **0.18 (0.07, 0.3)** | **0.00** | **0.15 (0.04, 0.26)** | **0.00** | **0.13 (-0.02, 0.28)** | **18.39** | **0.14 (0.03, 0.25)** | **0.00** |
| NSHD | ≥6 |  |  |  |  |  |  |  |  |
| ELSA | ≥6 | 0.25 (0.12, 0.38) | 87.60 | 0.21 (0.1, 0.33) | 83.58 | 0.21 (0.1, 0.33) | 78.05 | 0.19 (0.08, 0.31) | 88.94 |
| SNAC-K | ≥6 | 0.18 (-0.16, 0.52) | 12.40 | 0.02 (-0.3, 0.35) | 16.42 | 0.01 (-0.32, 0.33) | 21.95 | 0.01 (-0.32, 0.35) | 11.06 |
| RS |  |  |  |  |  |  |  |  |  |
| **Overall** | **≥6** | **0.24 (0.12, 0.36)** | **0.00** | **0.18 (0.04, 0.32)** | **13.42** | **0.17 (0, 0.34)** | **27.47** | **0.17 (0.06, 0.29)** | **0.00** |
| **Contact frequency (reference: never/almost never)** | | | | | | | | | |
| NSHD | >1/year |  |  |  |  |  |  |  |  |
| ELSA | >1/year | 0.5 (0.31, 0.69) | 86.19 | 0.39 (0.21, 0.56) | 87.67 | 0.39 (0.21, 0.56) | 87.43 | 0.37 (0.21, 0.54) | 88.20 |
| SNAC-K | >1/year | 0.35 (-0.12, 0.83) | 13.81 | 0.2 (-0.26, 0.65) | 12.33 | 0.17 (-0.28, 0.63) | 12.57 | 0.16 (-0.29, 0.61) | 11.80 |
| RS | >1/year |  |  |  |  |  |  |  |  |
| **Overall** | **>1/year** | **0.48 (0.3, 0.65)** | **0.00** | **0.36 (0.2, 0.52)** | **0.00** | **0.36 (0.2, 0.52)** | **0.00** | **0.35 (0.19, 0.5)** | **0.00** |
| NSHD | 1-2/month |  |  |  |  |  |  |  |  |
| ELSA | 1-2/month | 0.6 (0.41, 0.78) | 86.08 | 0.46 (0.29, 0.62) | 87.63 | 0.46 (0.29, 0.62) | 87.38 | 0.44 (0.28, 0.6) | 88.16 |
| SNAC-K | 1-2/month | 0.5 (0.04, 0.96) | 13.92 | 0.3 (-0.15, 0.75) | 12.37 | 0.28 (-0.16, 0.72) | 12.62 | 0.25 (-0.19, 0.69) | 11.84 |
| RS | 1-2/month |  |  |  |  |  |  |  |  |
| **Overall** | **1-2/month** | **0.58 (0.41, 0.75)** | **0.00** | **0.44 (0.28, 0.59)** | **0.00** | **0.43 (0.28, 0.59)** | **0.00** | **0.42 (0.26, 0.57)** | **0.00** |
| NSHD | 2-4/month |  |  |  |  |  |  |  |  |
| ELSA | 2-4/month | 0.51 (0.32, 0.7) | 85.84 | 0.41 (0.24, 0.58) | 87.42 | 0.41 (0.24, 0.58) | 87.17 | 0.39 (0.22, 0.55) | 87.93 |
| SNAC-K | 2-4/month | 0.51 (0.04, 0.97) | 14.16 | 0.33 (-0.12, 0.77) | 12.58 | 0.31 (-0.13, 0.75) | 12.83 | 0.29 (-0.15, 0.73) | 12.07 |
| RS | 2-4/month |  |  |  |  |  |  |  |  |
| **Overall** | **2-4/month** | **0.51 (0.34, 0.68)** | **0.00** | **0.4 (0.24, 0.55)** | **0.00** | **0.39 (0.23, 0.55)** | **0.00** | **0.38 (0.22, 0.53)** | **0.00** |
| NSHD | 2-3/week |  |  |  |  |  |  |  |  |
| ELSA | 2-3/week | 0.4 (0.2, 0.6) | 85.47 | 0.35 (0.17, 0.53) | 87.02 | 0.34 (0.16, 0.53) | 86.77 | 0.34 (0.16, 0.51) | 87.52 |
| SNAC-K | 2-3/week | 0.36 (-0.12, 0.85) | 14.53 | 0.21 (-0.26, 0.68) | 12.98 | 0.19 (-0.27, 0.65) | 13.23 | 0.15 (-0.31, 0.61) | 12.48 |
| RS | 2-3/week |  |  |  |  |  |  |  |  |
| **Overall** | **2-3/week** | **0.4 (0.21, 0.58)** | **0.00** | **0.33 (0.16, 0.5)** | **0.00** | **0.32 (0.16, 0.49)** | **0.00** | **0.32 (0.15, 0.48)** | **0.00** |
| **Social participation (reference: low <1 activity)** | | | | | | | | | |
| NSHD | moderate (2-3 activities) |  |  |  |  |  |  |  |  |
| ELSA | moderate (2-3 activities) | 0.25 (0.21, 0.29) | 96.90 | 0.18 (0.14, 0.22) | 96.80 | 0.18 (0.14, 0.22) | 96.87 | 0.16 (0.11, 0.2) | 97.01 |
| SNAC-K | moderate (2-3 activities) | 0.21 (-0.02, 0.45) | 3.10 | 0.17 (-0.06, 0.4) | 3.20 | 0.11 (-0.12, 0.34) | 3.13 | 0.11 (-0.13, 0.34) | 2.99 |
| RS | moderate (2-3 activities) |  |  |  |  |  |  |  |  |
| **Overall** | **moderate (2-3 activities)** | **0.25 (0.21, 0.29)** | **0.00** | **0.18 (0.14, 0.22)** | **0.01** | **0.18 (0.14, 0.22)** | **0.00** | **0.16 (0.11, 0.2)** | **0.00** |
| NSHD | high (≥4 or more activities) |  |  |  |  |  |  |  |  |
| ELSA | high (≥4 or more activities) | 0.55 (0.5, 0.59) | 80.17 | 0.35 (0.31, 0.39) | 95.82 | 0.35 (0.3, 0.39) | 71.41 | 0.32 (0.27, 0.36) | 85.03 |
| SNAC-K | high (≥4 or more activities) | 0.41 (0.2, 0.62) | 19.83 | 0.28 (0.07, 0.48) | 4.18 | 0.19 (-0.03, 0.4) | 28.59 | 0.19 (-0.03, 0.41) | 14.97 |
| RS | high (≥4 or more activities) |  |  |  |  |  |  |  |  |
| **Overall** | **high (≥4 or more activities)** | **0.52 (0.41, 0.63)** | **34.88** | **0.35 (0.31, 0.39)** | **0.00** | **0.3 (0.16, 0.44)** | **53** | **0.3 (0.21, 0.39)** | **24.10** |
| **Positive social support (reference: low <-1SD)** | | | | | | | | | |
| NSHD | -1SD to 0SD |  |  |  |  |  |  |  |  |
| ELSA | -1SD to 0SD | 0.08 (0.03, 0.13) | 64.92 | 0.05 (0, 0.1) | 71.51 | 0.05 (0, 0.09) | 71.29 | 0.02 (-0.03, 0.06) | 71.36 |
| SNAC-K | -1SD to 0SD | 0.05 (-0.06, 0.17) | 16.02 | 0.04 (-0.07, 0.15) | 13.09 | 0.03 (-0.08, 0.14) | 13.22 | 0.02 (-0.09, 0.13) | 12.99 |
| RS | -1SD to 0SD | 0 (-0.11, 0.1) | 19.06 | 0 (-0.1, 0.1) | 15.40 | -0.01 (-0.11, 0.09) | 15.49 | -0.01 (-0.11, 0.09) | 15.65 |
| **Overall** | **-1SD to 0SD** | **0.06 (0.01, 0.11)** | **10.36** | **0.04 (0, 0.08)** | **0.03** | **0.04 (0, 0.08)** | **0.01** | **0.01 (-0.03, 0.05)** | **0.02** |
| NSHD | 0SD to 1SD |  |  |  |  |  |  |  |  |
| ELSA | 0SD to 1SD | 0.1 (0.05, 0.15) | 55.83 | 0.07 (0.03, 0.12) | 66.57 | 0.07 (0.02, 0.12) | 66.31 | 0.03 (-0.02, 0.08) | 61.85 |
| SNAC-K | 0SD to 1SD | 0.17 (0.07, 0.26) | 21.98 | 0.12 (0.03, 0.22) | 16.74 | 0.13 (0.04, 0.22) | 16.91 | 0.11 (0.02, 0.21) | 18.70 |
| RS | 0SD to 1SD | 0.05 (-0.05, 0.14) | 22.19 | 0.05 (-0.04, 0.14) | 16.68 | 0.04 (-0.05, 0.13) | 16.78 | 0.03 (-0.06, 0.13) | 19.45 |
| **Overall** | **0SD to 1SD** | **0.1 (0.06, 0.15)** | **22.95** | **0.08 (0.04, 0.11)** | **0.00** | **0.07 (0.04, 0.11)** | **0.00** | **0.05 (0.01, 0.09)** | **10.64** |
| NSHD | >1 SD |  |  |  |  |  |  |  |  |
| ELSA | >1 SD | -0.05 (-0.11, 0) | 88.11 | -0.04 (-0.09, 0.01) | 67.99 | -0.04 (-0.09, 0.01) | 91.88 | -0.08 (-0.13, -0.03) | 92.19 |
| SNAC-K | >1 SD | -0.15 (-0.33, 0.03) | 11.89 | -0.18 (-0.36, -0.01) | 32.01 | -0.12 (-0.29, 0.05) | 8.12 | -0.1 (-0.28, 0.08) | 7.81 |
| RS | >1 SD |  |  |  |  |  |  |  |  |
| **Overall** | **>1 SD** | **-0.06 (-0.13, 0)** | **7.41** | **-0.09 (-0.22, 0.04)** | **56.93** | **-0.05 (-0.1, 0)** | **0.01** | **-0.08 (-0.13, -0.03)** | **0.00** |
| **Negative social support (reference: low <-1SD)** | | | | | | | | | |
| NSHD | -1SD to 0SD |  |  |  |  |  |  |  |  |
| ELSA | -1SD to 0SD | 0.17 (0.11, 0.22) | 100 | 0.13 (0.08, 0.18) | 100 | 0.13 (0.08, 0.18) | 100 | 0.1 (0.05, 0.15) | 100 |
| SNAC-K | -1SD to 0SD |  |  |  |  |  |  |  |  |
| RS | -1SD to 0SD |  |  |  |  |  |  |  |  |
| Overall | -1SD to 0SD |  |  |  |  |  |  |  |  |
| NSHD | 0SD to 1SD |  |  |  |  |  |  |  |  |
| ELSA | 0SD to 1SD | 0.19 (0.14, 0.25) | 100 | 0.14 (0.1, 0.19) | 100 | 0.14 (0.09, 0.19) | 100 | 0.11 (0.06, 0.16) | 100 |
| SNAC-K | 0SD to 1SD |  |  |  |  |  |  |  |  |
| RS | 0SD to 1SD |  |  |  |  |  |  |  |  |
| Overall | 0SD to 1SD |  |  |  |  |  |  |  |  |
| NSHD | >1 SD |  |  |  |  |  |  |  |  |
| ELSA | >1 SD | 0.02 (-0.04, 0.07) | 100 | 0.02 (-0.04, 0.07) | 100 | 0.02 (-0.04, 0.07) | 100 | -0.02 (-0.07, 0.04) | 100 |
| SNAC-K | >1 SD |  |  |  |  |  |  |  |  |
| RS | >1 SD |  |  |  |  |  |  |  |  |
| Overall | >1 SD |  |  |  |  |  |  |  |  |

### Supplementary Table 5: Social health at baseline and standardised processing speed score on average over time

*Adjustment 1= sex, age at baseline*

*Adjustment 2= sex, age at baseline, social class, education.*

*Adjustment 3= sex, age at baseline, social class, education, IADL, and vascular-related health conditions.*

*Blank cells indicate no data available*

|  |  | Adjustment 1 | | Adjustment 2 | | Adjustment 3 | | Additional adjustment for depression | |
| --- | --- | --- | --- | --- | --- | --- | --- | --- | --- |
| Study | Exposure | Coef (95% CI) | Weight/I^2^ | Coef (95% CI) | Weight/I^2^ | Coef (95% CI) | Weight/I^2^ | Coef (95% CI) | Weight/I^2^ |
| **Marital/cohabitation status (reference unmarried and alone)** | | | | | | | | | |
| NSHD | Married or cohabiting | 0.04 (-0.08, 0.17) | 17.83 | 0.06 (-0.07, 0.18) | 16.98 | 0.05 (-0.07, 0.17) | 16.39 | 0.05 (-0.07, 0.17) | 15.56 |
| ELSA | Married or cohabiting | 0.02 (-0.02, 0.06) | 29.87 | 0.01 (-0.04, 0.05) | 30.44 | 0 (-0.04, 0.05) | 30.85 | -0.01 (-0.06, 0.03) | 31.68 |
| SNAC-K | Married or cohabiting | 0.17 (0.1, 0.25) | 25.09 | 0.15 (0.07, 0.22) | 24.70 | 0.14 (0.06, 0.21) | 24.70 | 0.12 (0.04, 0.19) | 24.27 |
| RS | Married or cohabiting | 0.01 (-0.05, 0.07) | 27.21 | -0.01 (-0.06, 0.05) | 27.89 | 0 (-0.06, 0.05) | 28.06 | -0.01 (-0.07, 0.04) | 28.48 |
| **Overall** | **Married or cohabiting** | **0.06 (-0.02, 0.14)** | **79.80** | **0.05 (-0.02, 0.12)** | **76.90** | **0.04 (-0.02, 0.11)** | **74.87** | **0.03 (-0.03, 0.09)** | **70.64** |
| **Network size (references: none)** | | | | | | | | | |
| NSHD | 1-2 | 0.04 (-0.17, 0.25) | 29.98 | 0.09 (-0.11, 0.3) | 30.22 | 0.09 (-0.11, 0.3) | 30.28 | 0.09 (-0.11, 0.3) | 30.64 |
| ELSA | 1-2 | 0.04 (-0.11, 0.19) | 59.97 | 0.04 (-0.11, 0.18) | 59.82 | 0.04 (-0.11, 0.18) | 59.51 | 0.03 (-0.12, 0.18) | 59.51 |
| SNAC-K | 1-2 | 0.03 (-0.33, 0.4) | 10.05 | 0 (-0.35, 0.36) | 9.96 | 0 (-0.35, 0.35) | 10.22 | 0.04 (-0.33, 0.4) | 9.85 |
| RS | 1-2 |  |  |  |  |  |  |  |  |
| **Overall** | **1-2** | **0.04 (-0.07, 0.15)** | **0.00** | **0.05 (-0.06, 0.16)** | **0.00** | **0.05 (-0.06, 0.16)** | **0.00** | **0.05 (-0.06, 0.16)** | **0.00** |
| NSHD | 3-6 | 0.1 (-0.1, 0.29) | 31.27 | 0.12 (-0.07, 0.31) | 31.57 | 0.12 (-0.07, 0.31) | 31.64 | 0.11 (-0.08, 0.31) | 32.02 |
| ELSA | 3-6 | 0.08 (-0.07, 0.22) | 59.14 | 0.06 (-0.08, 0.2) | 58.95 | 0.06 (-0.08, 0.2) |  | 0.06 (-0.08, 0.2) | 58.64 |
| SNAC-K | 3-6 | 0.08 (-0.27, 0.44) | 9.58 | 0.03 (-0.32, 0.38) | 9.48 | 0 (-0.34, 0.35) | 9.72 | 0 (-0.35, 0.36) | 9.35 |
| RS | 3-6 |  |  |  |  |  |  |  |  |
| **Overall** | **3-6** | **0.08 (-0.03, 0.19)** | **0.00** | **0.08 (-0.03, 0.18)** | **0.00** | **0.07 (-0.03, 0.18)** | **0.00** | **0.07 (-0.04, 0.18)** | **0.00** |
| NSHD | ≥6 | 0.1 (-0.09, 0.29) | 32.38 | 0.14 (-0.05, 0.33) | 32.67 | 0.14 (-0.05, 0.33) | 32.74 | 0.13 (-0.06, 0.32) | 33.09 |
| ELSA | ≥6 | 0.12 (-0.02, 0.27) | 58.25 | 0.11 (-0.03, 0.25) | 58.09 | 0.11 (-0.03, 0.25) | 57.78 | 0.1 (-0.05, 0.24) | 57.82 |
| SNAC-K | ≥6 | 0.22 (-0.14, 0.57) | 9.37 | 0.13 (-0.22, 0.48) | 9.25 | 0.11 (-0.24, 0.46) | 9.48 | 0.1 (-0.26, 0.46) | 9.10 |
| RS | ≥6 |  |  |  |  |  |  |  |  |
| **Overall** | **≥6** | **0.13 (0.02, 0.23)** | **0.01** | **0.12 (0.01, 0.23)** | **0.00** | **0.12 (0.01, 0.23)** | **0.00** | **0.11 (0, 0.22)** | **0.01** |
| **Contact frequency (reference: never/almost never)** | | | | | | | | | |
| NSHD | >1/year | 0.11 (-0.3, 0.51) | 25.05 | 0.04 (-0.36, 0.44) | 24.28 | 0.05 (-0.35, 0.45) | 24.21 | 0.04 (-0.36, 0.44) | 24.25 |
| ELSA | >1/year | 0.07 (-0.18, 0.33) | 56.57 | 0.03 (-0.23, 0.28) | 58.52 | 0.03 (-0.23, 0.28) | 58.25 | 0.02 (-0.24, 0.27) | 58.04 |
| SNAC-K | >1/year | 0.51 (0.03, 0.98) | 18.38 | 0.4 (-0.07, 0.87) | 17.20 | 0.36 (-0.11, 0.83) | 17.55 | 0.35 (-0.11, 0.82) | 17.71 |
| RS | >1/year |  |  |  |  |  |  |  |  |
| **Overall** | **>1/year** | **0.16 (-0.05, 0.37)** | **0.00** | **0.1 (-0.1, 0.29)** | **0.00** | **0.09 (-0.1, 0.29)** | **0.00** | **0.08 (-0.11, 0.28)** | **0.00** |
| NSHD | 1-2/month | 0.07 (-0.32, 0.47) | 23.96 | 0 (-0.39, 0.39) | 24.61 | 0.01 (-0.38, 0.4) | 24.55 | 0.01 (-0.38, 0.39) | 24.59 |
| ELSA | 1-2/month | 0.14 (-0.12, 0.39) | 58.75 | 0.09 (-0.17, 0.34) | 57.93 | 0.08 (-0.17, 0.34) | 57.65 | 0.07 (-0.18, 0.33) | 57.43 |
| SNAC-K | 1-2/month | 0.52 (0.05, 0.98) | 17.29 | 0.38 (-0.08, 0.85) | 17.45 | 0.36 (-0.1, 0.81) | 17.81 | 0.34 (-0.12, 0.79) | 17.98 |
| RS | 1-2/month |  |  |  |  |  |  |  |  |
| **Overall** | **1-2/month** | **0.19 (-0.01, 0.38)** | **0.00** | **0.12 (-0.08, 0.31)** | **0.00** | **0.12 (-0.08, 0.31)** | **0.00** | **0.1 (-0.09, 0.3)** | **0.00** |
| NSHD | 2-4/month | 0.15 (-0.24, 0.54) | 25.15 | 0.1 (-0.28, 0.48) | 25.81 | 0.11 (-0.27, 0.49) | 25.74 | 0.1 (-0.28, 0.48) | 25.78 |
| ELSA | 2-4/month | 0.15 (-0.1, 0.41) | 57.66 | 0.11 (-0.14, 0.37) | 56.84 | 0.11 (-0.14, 0.37) | 56.56 | 0.1 (-0.15, 0.36) | 56.35 |
| SNAC-K | 2-4/month | 0.51 (0.05, 0.98) | 17.19 | 0.39 (-0.07, 0.86) | 17.35 | 0.37 (-0.09, 0.83) | 17.70 | 0.34 (-0.12, 0.79) | 17.87 |
| RS | 2-4/month |  |  |  |  |  |  |  |  |
| **Overall** | **2-4/month** | **0.22 (0.02, 0.41)** | **0.00** | **0.16 (-0.03, 0.35)** | **0.00** | **0.16 (-0.03, 0.35)** | **0.00** | **0.15 (-0.05, 0.34)** | **0.00** |
| NSHD | 2-3/week | 0.05 (-0.34, 0.44) | 27.18 | 0.02 (-0.37, 0.4) | 27.82 | 0.03 (-0.36, 0.41) | 27.75 | 0.01 (-0.37, 0.4) | 27.79 |
| ELSA | 2-3/week | 0.16 (-0.11, 0.43) | 55.68 | 0.14 (-0.13, 0.41) | 54.90 | 0.13 (-0.14, 0.41) | 54.63 | 0.13 (-0.14, 0.4) | 54.44 |
| SNAC-K | 2-3/week | 0.43 (-0.06, 0.92) | 17.14 | 0.32 (-0.16, 0.81) | 17.27 | 0.3 (-0.18, 0.78) | 17.62 | 0.27 (-0.2, 0.75) | 17.76 |
| RS | 2-3/week |  |  |  |  |  |  |  |  |
| **Overall** | **2-3/week** | **0.18 (-0.02, 0.38)** | **0.00** | **0.14 (-0.07, 0.34)** | **0.00** | **0.13 (-0.07, 0.33)** | **0.00** | **0.12 (-0.08, 0.33)** | **0.00** |
| **Social participation (reference: low <1 activity)** | | | | | | | | | |
| NSHD | moderate (2-3 activities) |  |  |  |  |  |  |  |  |
| ELSA | moderate (2-3 activities) | 0.12 (0.06, 0.17) | 95.94 | 0.1 (0.05, 0.15) | 95.83 | 0.09 (0.04, 0.14) | 71.50 | 0.08 (0.02, 0.13) | 75.50 |
| SNAC-K | moderate (2-3 activities) | 0 (-0.25, 0.24) | 4.06 | -0.02 (-0.27, 0.22) | 4.17 | -0.09 (-0.34, 0.15) | 28.50 | -0.1 (-0.35, 0.15) | 24.50 |
| RS | moderate (2-3 activities) |  |  |  |  |  |  |  |  |
| **Overall** | **moderate (2-3 activities)** | **0.11 (0.06, 0.16)** | **0.00** | **0.09 (0.04, 0.14)** | **0.00** | **0.04 (-0.13, 0.2)** | **53.13** | **0.03 (-0.12, 0.18)** | **44.62** |
| NSHD | high (≥4 or more activities) |  |  |  |  |  |  |  |  |
| ELSA | high (≥4 or more activities) | 0.21 (0.16, 0.26) | 95.51 | 0.15 (0.1, 0.2) | 94.87 | 0.14 (0.09, 0.19) | 94.99 | 0.12 (0.07, 0.17) | 95.19 |
| SNAC-K | high (≥4 or more activities) | 0.3 (0.08, 0.52) | 4.49 | 0.23 (0.01, 0.45) | 5.13 | 0.12 (-0.11, 0.34) | 5.01 | 0.1 (-0.13, 0.33) | 4.81 |
| RS | high (≥4 or more activities) |  |  |  |  |  |  |  |  |
| **Overall** | **high (≥4 or more activities)** | **0.22 (0.17, 0.26)** | **0.00** | **0.15 (0.1, 0.2)** | **0.01** | **0.14 (0.09, 0.19)** | **0.00** | **0.12 (0.07, 0.17)** | **0.00** |
| **Positive social support (reference: low <-1SD)** | | | | | | | | | |
| NSHD | -1SD to 0SD | 0.09 (-0.02, 0.21) | 13.37 | 0.08 (-0.03, 0.2) | 14.23 | 0.08 (-0.03, 0.2) | 13.59 | 0.08 (-0.03, 0.19) | 15.78 |
| ELSA | -1SD to 0SD | 0.03 (-0.02, 0.09) | 57.71 | 0.02 (-0.03, 0.07) | 53.65 | 0.02 (-0.04, 0.07) | 55.08 | 0 (-0.05, 0.06) | 48.82 |
| SNAC-K | -1SD to 0SD | 0.03 (-0.09, 0.15) | 12.64 | 0.02 (-0.1, 0.14) | 13.25 | 0.01 (-0.11, 0.12) | 12.94 | -0.02 (-0.14, 0.09) | 14.73 |
| RS | -1SD to 0SD | 0.1 (0, 0.2) | 16.27 | 0.1 (0, 0.2) | 18.87 | 0.09 (0, 0.19) | 18.39 | 0.08 (-0.01, 0.18) | 20.67 |
| **Overall** | **-1SD to 0SD** | **0.05 (0.01, 0.09)** | **0.00** | **0.04 (0, 0.09)** | **5.56** | **0.04 (0, 0.08)** | **1.92** | **0.03 (-0.02, 0.08)** | **16.19** |
| NSHD | 0SD to 1SD | 0.13 (0.01, 0.25) | 14.95 | 0.12 (0, 0.23) | 16.84 | 0.11 (0, 0.23) | 15.77 | 0.11 (-0.01, 0.22) | 17.91 |
| ELSA | 0SD to 1SD | 0.03 (-0.02, 0.09) | 43.79 | 0.02 (-0.03, 0.08) | 38.11 | 0.02 (-0.03, 0.07) | 40.19 | 0 (-0.05, 0.05) | 35.90 |
| SNAC-K | 0SD to 1SD | 0.05 (-0.05, 0.15) | 20.02 | 0.02 (-0.07, 0.12) | 21.14 | 0.03 (-0.07, 0.12) | 20.61 | -0.02 (-0.12, 0.08) | 21.60 |
| RS | 0SD to 1SD | 0.12 (0.02, 0.21) | 21.24 | 0.12 (0.04, 0.21) | 23.91 | 0.11 (0.03, 0.2) | 23.44 | 0.1 (0.01, 0.18) | 24.59 |
| **Overall** | **0SD to 1SD** | **0.07 (0.02, 0.12)** | **27.50** | **0.06 (0.01, 0.12)** | **43.79** | **0.06 (0.01, 0.11)** | **36.18** | **0.04 (-0.02, 0.1)** | **51.56** |
| NSHD | >1 SD | 0.07 (-0.06, 0.2) | 33.12 | 0.06 (-0.06, 0.18) | 33.33 | 0.06 (-0.07, 0.18) | 17.72 | 0.04 (-0.08, 0.17) | 17.80 |
| ELSA | >1 SD | 0.01 (-0.05, 0.07) | 41.39 | 0.01 (-0.05, 0.07) | 40.61 | 0.01 (-0.05, 0.07) | 74.18 | -0.01 (-0.07, 0.05) | 74.37 |
| SNAC-K | >1 SD | -0.21 (-0.4, -0.03) | 25.49 | -0.22 (-0.41, -0.04) | 26.06 | -0.14 (-0.32, 0.04) | 8.10 | -0.16 (-0.34, 0.03) | 7.83 |
| RS | >1 SD |  |  |  |  |  |  |  |  |
| **Overall** | **>1 SD** | **-0.03 (-0.17, 0.11)** | **76.86** | **-0.03 (-0.18, 0.11)** | **79.07** | **0.01 (-0.05, 0.06)** | **0.00** | **-0.01 (-0.06, 0.04)** | **0.00** |
| **Negative social support (reference: low <-1SD)** | | | | | | | | | |
| NSHD | -1SD to 0SD | 0.05 (-0.06, 0.16) | 19.14 | 0.05 (-0.06, 0.16) | 19.13 | 0.04 (-0.07, 0.16) | 19.33 | 0.08 (-0.04, 0.19) | 18.20 |
| ELSA | -1SD to 0SD | 0.01 (-0.05, 0.06) | 80.86 | 0.01 (-0.05, 0.06) | 80.87 | -0.01 (-0.06, 0.05) | 80.67 | 0.05 (-0.01, 0.1) | 81.80 |
| SNAC-K | -1SD to 0SD |  |  |  |  |  |  |  |  |
| RS | -1SD to 0SD |  |  |  |  |  |  |  |  |
| **Overall** | **-1SD to 0SD** | **0.02 (-0.03, 0.07)** | **0.02** | **0.02 (-0.03, 0.06)** | **0.01** | **0 (-0.04, 0.05)** | **0.04** | **0.05 (0, 0.1)** | **0.01** |
| NSHD | 0SD to 1SD | 0.07 (-0.05, 0.18) | 18.45 | 0.07 (-0.05, 0.18) | 18.45 | 0.05 (-0.07, 0.16) | 18.49 | -0.01 (-0.13, 0.11) | 21.09 |
| ELSA | 0SD to 1SD | 0.03 (-0.02, 0.09) | 81.55 | 0.03 (-0.02, 0.09) | 81.55 | 0.01 (-0.04, 0.07) | 81.51 | 0.02 (-0.04, 0.08) | 78.91 |
| SNAC-K | 0SD to 1SD |  |  |  |  |  |  |  |  |
| RS | 0SD to 1SD |  |  |  |  |  |  |  |  |
| **Overall** | **0SD to 1SD** | **0.04 (-0.01, 0.09)** | **0.00** | **0.04 (-0.01, 0.09)** | **0.01** | **0.02 (-0.03, 0.07)** | **0.02** | **0.01 (-0.04, 0.07)** | **0.00** |
| NSHD | >1 SD | -0.01 (-0.13, 0.11) | 21.09 | 0.03 (-0.09, 0.15) | 21.37 | 0.03 (-0.09, 0.15) | 21.37 | 0.01 (-0.11, 0.12) | 21.02 |
| ELSA | >1 SD | 0.02 (-0.04, 0.08) | 78.91 | 0.02 (-0.04, 0.08) | 78.63 | 0.02 (-0.04, 0.08) | 78.63 | 0 (-0.06, 0.06) | 78.98 |
| SNAC-K | >1 SD |  |  |  |  |  |  |  |  |
| RS | >1 SD |  |  |  |  |  |  |  |  |
| **Overall** | **>1 SD** | **0.01 (-0.04, 0.07)** | **0.00** | **0.02 (-0.03, 0.08)** | **0.01** | **0.02 (-0.03, 0.08)** | **0.01** | **0 (-0.05, 0.06)** | **0.01** |

### Supplementary Table 6: Social health at baseline and global/composite cognition on average over time

*Adjustment 1= sex, age at baseline*

*Adjustment 2= sex, age at baseline, social class, education.*

*Adjustment 3= sex, age at baseline, social class, education, IADL, and vascular-related health conditions.*

*Blank cells indicate no data available*

|  |  | Adjustment 1 | | Adjustment 2 | | Adjustment 3 | | Additional adjustment for depression | |
| --- | --- | --- | --- | --- | --- | --- | --- | --- | --- |
| Study | Exposure | Coef (95% CI) | Weight/I^2^ | Coef (95% CI) | Weight/I^2^ | Coef (95% CI) | Weight/I^2^ | Coef (95% CI) | Weight/I^2^ |
| **Marital/cohabitation status (reference unmarried and alone)** | | | | | | | | | |
| NSHD | Married or cohabiting | -0.01 (-0.13, 0.11) | 20.41 | 0.02 (-0.09, 0.13) | 18.85 | 0.01 (-0.1, 0.13) | 18.33 | 0.01 (-0.1, 0.12) | 17.35 |
| ELSA | Married or cohabiting | 0.1 (0.05, 0.14) | 27.71 | 0.06 (0.02, 0.1) | 29.22 | 0.05 (0.01, 0.09) | 29.54 | 0.02 (-0.02, 0.06) | 30.51 |
| SNAC-K | Married or cohabiting | 0.24 (0.17, 0.31) | 25.48 | 0.19 (0.12, 0.26) | 25.09 | 0.18 (0.12, 0.25) | 25.15 | 0.16 (0.09, 0.23) | 24.85 |
| RS | Married or cohabiting | 0.05 (-0.02, 0.11) | 26.40 | 0.03 (-0.02, 0.09) | 26.84 | 0.04 (-0.02, 0.09) | 26.98 | 0.03 (-0.03, 0.09) | 27.29 |
| **Overall** | **Married or cohabiting** | **0.1 (0, 0.2)** | **89.31** | **0.08 (0, 0.15)** | **82.67** | **0.07 (0, 0.15)** | **81.17** | **0.06 (-0.01, 0.12)** | **77.02** |
| **Network size (reference: none)** | | | | | | | | | |
| NSHD | 1-2 | -0.06 (-0.27, 0.15) | 28.97 | 0.03 (-0.16, 0.22) | 29.52 | 0.03 (-0.15, 0.22) | 29.61 | 0.03 (-0.16, 0.22) | 30.30 |
| ELSA | 1-2 | 0.09 (-0.06, 0.23) | 58.95 | 0.09 (-0.05, 0.22) | 59.64 | 0.09 (-0.05, 0.22) | 59.21 | 0.08 (-0.05, 0.21) | 58.73 |
| SNAC-K | 1-2 | -0.02 (-0.34, 0.3) | 12.07 | -0.08 (-0.39, 0.23) |  | -0.09 (-0.4, 0.22) | 11.18 | -0.07 (-0.38, 0.24) | 10.97 |
| RS | 1-2 |  |  |  |  |  |  |  |  |
| **Overall** | **1-2** | **0.03 (-0.08, 0.14)** | **0.83** | **0.05 (-0.05, 0.15)** | **0.00** | **0.05 (-0.05, 0.15)** | **0.00** | **0.05 (-0.05, 0.15)** | **0.01** |
| NSHD | 3-6 | 0.01 (-0.18, 0.21) | 31.45 | 0.06 (-0.12, 0.23) | 30.89 | 0.06 (-0.12, 0.23) | 31.00 | 0.05 (-0.13, 0.23) | 31.75 |
| ELSA | 3-6 | 0.17 (0.03, 0.31) | 55.88 | 0.13 (0.01, 0.26) | 58.79 | 0.13 (0.01, 0.26) | 58.35 | 0.12 (-0.01, 0.25) | 57.81 |
| SNAC-K | 3-6 | 0.13 (-0.19, 0.44) | 12.68 | 0.03 (-0.28, 0.33) | 10.32 | 0 (-0.3, 0.3) |  | -0.01 (-0.32, 0.3) | 10.44 |
| RS | 3-6 |  |  |  |  |  |  |  |  |
| **Overall** | **3-6** | **0.11 (0, 0.23)** | **8.78** | **0.1 (0, 0.2)** | **0.00** | **0.09 (0, 0.19)** | **0.00** | **0.09 (-0.01, 0.18)** | **0.00** |
| NSHD | ≥6 | 0 (-0.19, 0.19) | 35.60 | 0.08 (-0.09, 0.25) | 31.99 | 0.08 (-0.09, 0.25) | 32.12 | 0.07 (-0.11, 0.24) | 32.81 |
| ELSA | ≥6 | 0.24 (0.1, 0.38) | 44.44 | 0.2 (0.07, 0.32) | 57.97 | 0.19 (0.07, 0.32) | 57.52 | 0.17 (0.04, 0.3) | 57.06 |
| SNAC-K | ≥6 | 0.27 (-0.05, 0.58) | 19.96 | 0.11 (-0.2, 0.41) | 10.04 | 0.08 (-0.22, 0.39) | 10.37 | 0.08 (-0.23, 0.38) | 10.13 |
| RS | ≥6 |  |  |  |  |  |  |  |  |
| **Overall** | **≥6** | **0.16 (-0.01, 0.33)** | **53.63** | **0.15 (0.05, 0.25)** | **0.00** | **0.15 (0.05, 0.24)** | **0.00** | **0.13 (0.03, 0.23)** | **0.00** |
| **Contact frequency (reference: never/almost never)** | | | | | | | | | |
| NSHD | >1/year | 0.11 (-0.29, 0.51) | 20.63 | -0.06 (-0.42, 0.3) | 27.87 | -0.05 (-0.41, 0.31) | 26.93 | -0.06 (-0.42, 0.3) | 26.65 |
| ELSA | >1/year | 0.43 (0.2, 0.66) | 62.77 | 0.31 (0.09, 0.52) | 50.23 | 0.31 (0.09, 0.52) | 51.77 | 0.29 (0.08, 0.5) | 52.13 |
| SNAC-K | >1/year | 0.48 (0.03, 0.93) | 16.60 | 0.32 (-0.11, 0.75) | 21.91 | 0.29 (-0.13, 0.71) | 21.30 | 0.26 (-0.16, 0.68) | 21.22 |
| RS |  |  |  |  |  |  |  |  |  |
| **Overall** | **>1/year** | **0.37 (0.19, 0.55)** | **0.47** | **0.21 (-0.02, 0.44)** | **35.98** | **0.21 (-0.01, 0.43)** | **30.21** | **0.19 (-0.03, 0.41)** | **29.59** |
| NSHD | 1-2/month | 0.01 (-0.39, 0.4) | 30.49 | -0.16 (-0.51, 0.2) | 31.71 | -0.15 (-0.5, 0.21) | 31.46 | -0.16 (-0.51, 0.2) | 31.24 |
| ELSA | 1-2/month | 0.54 (0.32, 0.77) | 41.70 | 0.39 (0.18, 0.61) | 40.28 | 0.39 (0.18, 0.61) | 40.59 | 0.37 (0.16, 0.58) | 41.01 |
| SNAC-K | 1-2/month | 0.56 (0.12, 0.99) | 27.81 | 0.36 (-0.06, 0.78) | 28.02 | 0.33 (-0.08, 0.74) | 27.95 | 0.29 (-0.12, 0.7) | 27.75 |
| RS | 1-2/month |  |  |  |  |  |  |  |  |
| Overall | 1-2/month | 0.38 (0.04, 0.73) | 65.85 | 0.21 (-0.14, 0.56) | 71.15 | 0.21 (-0.13, 0.54) | 69.48 | 0.18 (-0.14, 0.51) | 68.03 |
| NSHD | 2-4/month | 0.06 (-0.32, 0.45) | 29.24 | -0.06 (-0.41, 0.29) | 30.80 | -0.05 (-0.4, 0.3) | 30.33 | -0.06 (-0.41, 0.29) | 29.91 |
| ELSA | 2-4/month | 0.47 (0.25, 0.7) | 45.86 | 0.36 (0.15, 0.58) | 44.23 | 0.36 (0.14, 0.57) | 45.00 | 0.34 (0.13, 0.55) | 45.80 |
| SNAC-K | 2-4/month | 0.57 (0.14, 1.01) | 24.90 | 0.39 (-0.03, 0.82) | 24.97 | 0.37 (-0.05, 0.78) | 24.68 | 0.33 (-0.08, 0.75) | 24.29 |
| **Overall** | **2-4/month** | **0.38 (0.1, 0.66)** | **49.32** | **0.24 (-0.04, 0.52)** | **54.93** | **0.24 (-0.03, 0.5)** | **51.40** | **0.22 (-0.04, 0.47)** | **48.75** |
| NSHD | 2-3/week | -0.04 (-0.42, 0.35) | 29.33 | -0.14 (-0.49, 0.21) | 31.56 | -0.13 (-0.48, 0.22) | 31.17 | -0.14 (-0.49, 0.21) | 31.18 |
| ELSA | 2-3/week | 0.36 (0.12, 0.6) | 47.46 | 0.3 (0.07, 0.53) | 44.02 | 0.3 (0.07, 0.52) | 44.63 | 0.29 (0.07, 0.52) | 44.30 |
| SNAC-K | 2-3/week | 0.43 (-0.03, 0.89) | 23.21 | 0.27 (-0.17, 0.72) | 24.42 | 0.24 (-0.19, 0.68) | 24.20 | 0.19 (-0.24, 0.63) | 24.52 |
| RS | 2-3/week |  |  |  |  |  |  |  |  |
| **Overall** | **2-3/week** | **0.26 (0, 0.52)** | **41.01** | **0.16 (-0.12, 0.44)** | **53.74** | **0.15 (-0.12, 0.42)** | **50.60** | **0.13 (-0.14, 0.41)** | **52.36** |
| **Social participation (reference: low <1 activity)** | | | | | | | | | |
| NSHD | moderate (2-3 activities) |  |  |  |  |  |  |  |  |
| ELSA | moderate (2-3 activities) | 0.27 (0.22, 0.31) | 95.75 | 0.2 (0.15, 0.24) | 95.65 | 0.19 (0.15, 0.24) | 85.70 | 0.17 (0.12, 0.21) | 84.14 |
| SNAC-K | moderate (2-3 activities) | 0.19 (-0.03, 0.41) | 4.25 | 0.15 (-0.06, 0.36) | 4.35 | 0.07 (-0.15, 0.28) | 14.30 | 0.03 (-0.19, 0.25) | 15.86 |
| RS | moderate (2-3 activities) |  |  |  |  |  |  |  |  |
| **Overall** | **moderate (2-3 activities)** | **0.26 (0.22, 0.31)** | **0.00** | **0.19 (0.15, 0.24)** | **0.00** | **0.17 (0.09, 0.26)** | **21.90** | **0.14 (0.05, 0.24)** | **25.65** |
| NSHD | high (≥4 or more activities) |  |  |  |  |  |  |  |  |
| ELSA | high (≥4 or more activities) | 0.59 (0.55, 0.63) | 95.19 | 0.38 (0.33, 0.42) | 94.55 | 0.37 (0.32, 0.42) | 94.77 | 0.33 (0.29, 0.38) | 94.97 |
| SNAC-K | high (≥4 or more activities) | 0.52 (0.33, 0.72) | 4.81 | 0.4 (0.21, 0.59) | 5.45 | 0.27 (0.08, 0.47) | 5.23 | 0.25 (0.05, 0.45) | 5.03 |
| RS | high (≥4 or more activities) |  |  |  |  |  |  |  |  |
| **Overall** | **high (≥4 or more activities)** | **0.59 (0.54, 0.63)** | **0.00** | **0.38 (0.33, 0.42)** | **0.00** | **0.37 (0.32, 0.41)** | **0.01** | **0.33 (0.28, 0.37)** | **0.00** |
| **Positive social support (reference: low <-1SD)** | | | | | | | | | |
| NSHD | -1SD to 0SD | 0.11 (-0.01, 0.22) | 12.87 | 0.08 (-0.02, 0.18) | 13.71 | 0.08 (-0.02, 0.19) | 13.62 | 0.08 (-0.02, 0.18) | 13.77 |
| ELSA | -1SD to 0SD | 0.09 (0.04, 0.14) | 57.41 | 0.06 (0.01, 0.11) | 57.91 | 0.06 (0.01, 0.11) | 57.52 | 0.02 (-0.03, 0.07) | 57.40 |
| SNAC-K | -1SD to 0SD | 0.02 (-0.09, 0.13) | 13.62 | 0 (-0.11, 0.11) | 12.77 | -0.01 (-0.11, 0.1) | 13.06 | -0.03 (-0.14, 0.07) | 12.87 |
| RS | -1SD to 0SD | 0.07 (-0.03, 0.17) | 16.11 | 0.07 (-0.03, 0.17) | 15.61 | 0.06 (-0.03, 0.16) | 15.79 | 0.06 (-0.04, 0.15) | 15.95 |
| **Overall** | **-1SD to 0SD** | **0.08 (0.04, 0.12)** | **0.04** | **0.05 (0.02, 0.09)** | **0.01** | **0.05 (0.01, 0.09)** | **0.01** | **0.03 (-0.01, 0.07)** | **0.00** |
| NSHD | 0SD to 1SD | 0.14 (0.03, 0.26) | 10.95 | 0.11 (0, 0.21) | 11.79 | 0.11 (0, 0.21) | 11.71 | 0.1 (-0.01, 0.2) | 11.96 |
| ELSA | 0SD to 1SD | 0.11 (0.06, 0.17) | 53.83 | 0.08 (0.03, 0.13) | 54.40 | 0.08 (0.03, 0.13) | 53.93 | 0.03 (-0.01, 0.08) | 53.79 |
| SNAC-K | 0SD to 1SD | 0.12 (0.03, 0.21) | 17.74 | 0.07 (-0.01, 0.16) | 16.69 | 0.08 (-0.01, 0.17) | 17.05 | 0.05 (-0.04, 0.14) | 16.58 |
| RS | 0SD to 1SD | 0.11 (0.02, 0.2) | 17.48 | 0.11 (0.02, 0.2) | 17.12 | 0.1 (0.01, 0.19) | 17.31 | 0.09 (0, 0.17) | 17.67 |
| **Overall** | **0SD to 1SD** | **0.12 (0.08, 0.16)** | **0.02** | **0.09 (0.05, 0.12)** | **0.02** | **0.09 (0.05, 0.12)** | **0.02** | **0.05 (0.02, 0.09)** | **0.01** |
| NSHD | >1 SD | 0.13 (0, 0.25) | 33.27 | 0.1 (-0.01, 0.21) | 33.51 | 0.1 (-0.01, 0.21) | 33.47 | 0.08 (-0.03, 0.2) | 33.42 |
| ELSA | >1 SD | -0.02 (-0.07, 0.04) | 36.16 | -0.01 (-0.06, 0.05) | 35.68 | -0.01 (-0.06, 0.05) | 37.72 | -0.05 (-0.1, 0.01) | 39.73 |
| SNAC-K | >1 SD | -0.28 (-0.45, -0.11) | 30.58 | -0.31 (-0.48, -0.15) | 30.80 | -0.22 (-0.38, -0.06) | 28.81 | -0.2 (-0.37, -0.03) | 26.84 |
| RS | >1 SD |  |  |  |  |  |  |  |  |
| **Overall** | **>1 SD** | **-0.05 (-0.27, 0.17)** | **91.46** | **-0.06 (-0.3, 0.17)** | **93.16** | **-0.03 (-0.2, 0.14)** | **87.23** | **-0.04 (-0.19, 0.1)** | **81.67** |
| **Negative social support (reference: low <-1SD)** | | | | | | | | | |
| NSHD | -1SD to 0SD | 0.1 (-0.01, 0.22) | 18.64 | 0.1 (-0.01, 0.2) | 19.59 | 0.1 (0, 0.2) | 19.60 | 0.09 (-0.02, 0.19) | 19.75 |
| ELSA | -1SD to 0SD | 0.15 (0.1, 0.2) | 81.36 | 0.11 (0.06, 0.16) | 80.41 | 0.11 (0.06, 0.16) | 80.40 | 0.08 (0.03, 0.13) | 80.25 |
| SNAC-K | -1SD to 0SD |  |  |  |  |  |  |  |  |
| RS | -1SD to 0SD |  |  |  |  |  |  |  |  |
| **Overall** | **-1SD to 0SD** | **0.14 (0.09, 0.19)** | **0.02** | **0.11 (0.06, 0.15)** | **0.02** | **0.11 (0.06, 0.15)** | **0.01** | **0.08 (0.03, 0.12)** | **0.02** |
| NSHD | 0SD to 1SD | 0.18 (0.07, 0.29) | 18.29 | 0.15 (0.05, 0.26) | 19.04 | 0.15 (0.05, 0.26) | 19.04 | 0.13 (0.02, 0.23) | 19.02 |
| ELSA | 0SD to 1SD | 0.19 (0.13, 0.24) | 81.71 | 0.14 (0.09, 0.19) | 80.96 | 0.13 (0.08, 0.18) | 80.96 | 0.09 (0.04, 0.14) | 80.98 |
| SNAC-K | 0SD to 1SD |  |  |  |  |  |  |  |  |
| RS | 0SD to 1SD |  |  |  |  |  |  |  |  |
| **Overall** | **0SD to 1SD** | **0.19 (0.14, 0.24)** | **0.00** | **0.14 (0.09, 0.19)** | **0.02** | **0.14 (0.09, 0.18)** | **0.01** | **0.1 (0.05, 0.14)** | **0.03** |
| NSHD | >1 SD | 0.01 (-0.11, 0.12) | 20.94 | 0.08 (-0.02, 0.19) | 21.83 | 0.08 (-0.02, 0.19) | 21.82 | 0.05 (-0.05, 0.16) | 25.53 |
| ELSA | >1 SD | 0.03 (-0.03, 0.09) | 79.06 | 0.03 (-0.03, 0.08) | 78.17 | 0.03 (-0.03, 0.08) | 78.18 | -0.01 (-0.07, 0.04) | 74.47 |
| SNAC-K | >1 SD |  |  |  |  |  |  |  |  |
| RS | >1 SD |  |  |  |  |  |  |  |  |
| **Overall** | **>1 SD** | **0.02 (-0.03, 0.08)** | **0.02** | **0.04 (-0.01, 0.09)** | **0.01** | **0.04 (-0.01, 0.09)** | **0.00** | **0 (-0.05, 0.06)** | **14.63** |

### Supplementary figure 1: Sex-stratified marital/cohabitation status and average cognitive capability

*Adjusted for age at baseline, social class, education, IADL, and vascular-related health conditions*

#### Memory

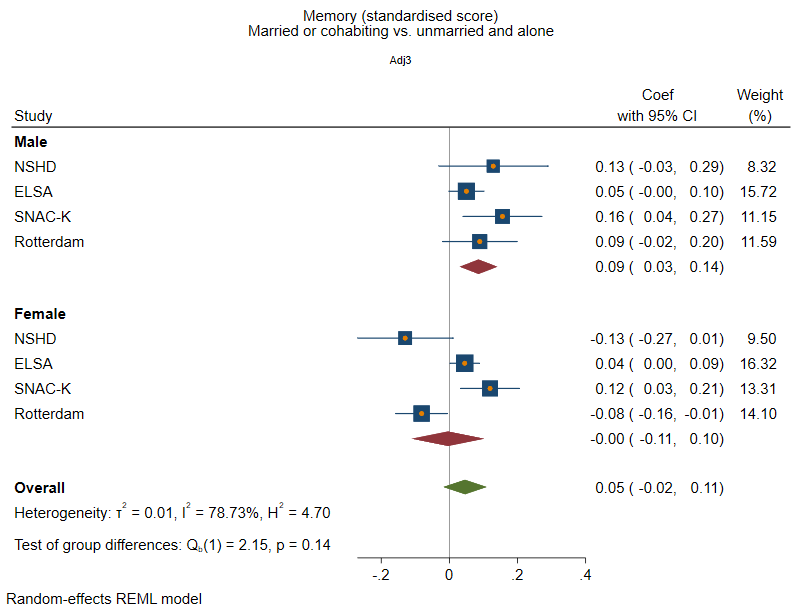

#### Executive Function

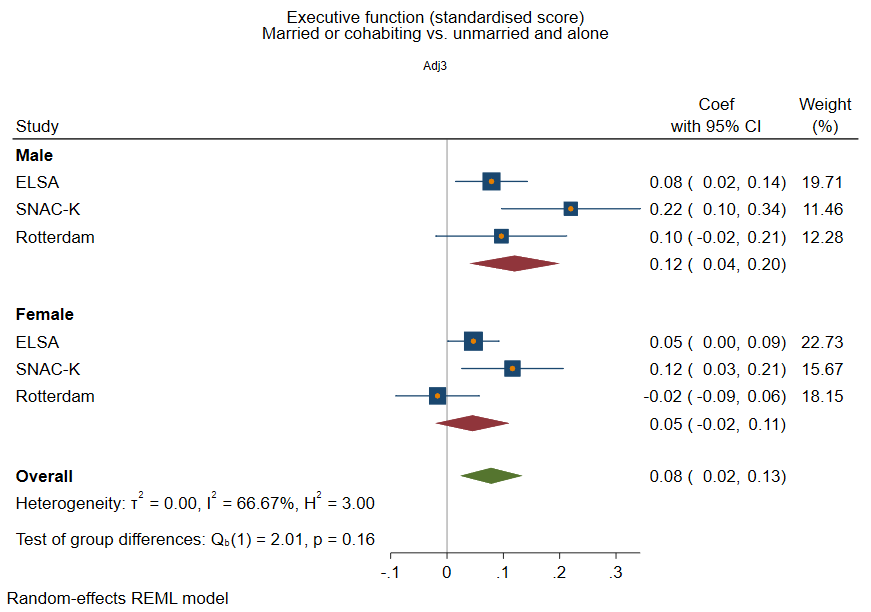

#### Processing Speed

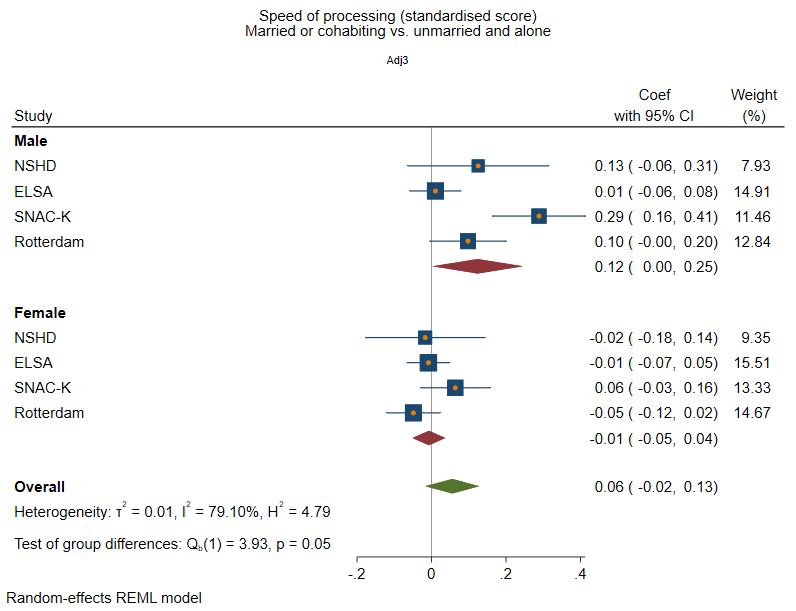

#### Global/composite cognition

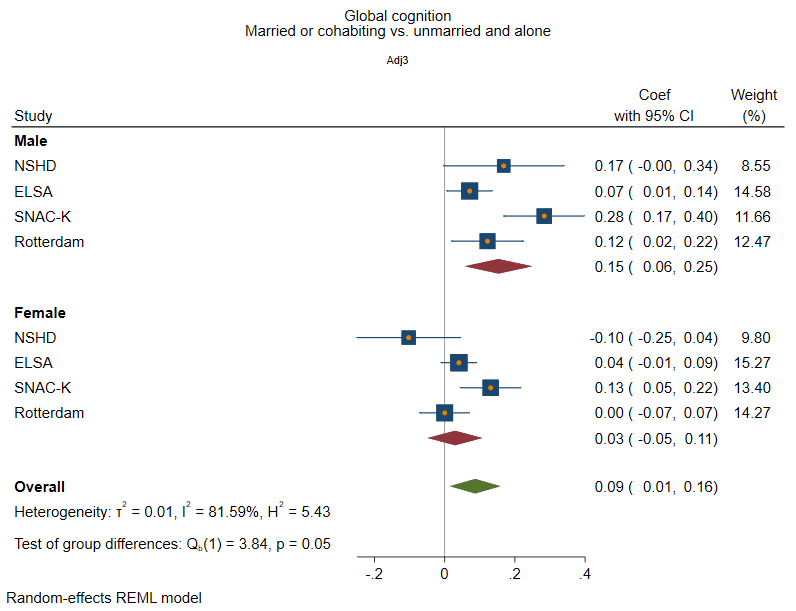

### Supplementary figure 2: Structural social health at baseline and rate of decline in memory

*Adjusted for sex and age at baseline, social class, education, IADL, and vascular-related health conditions.*

#### Marital/cohabitation status

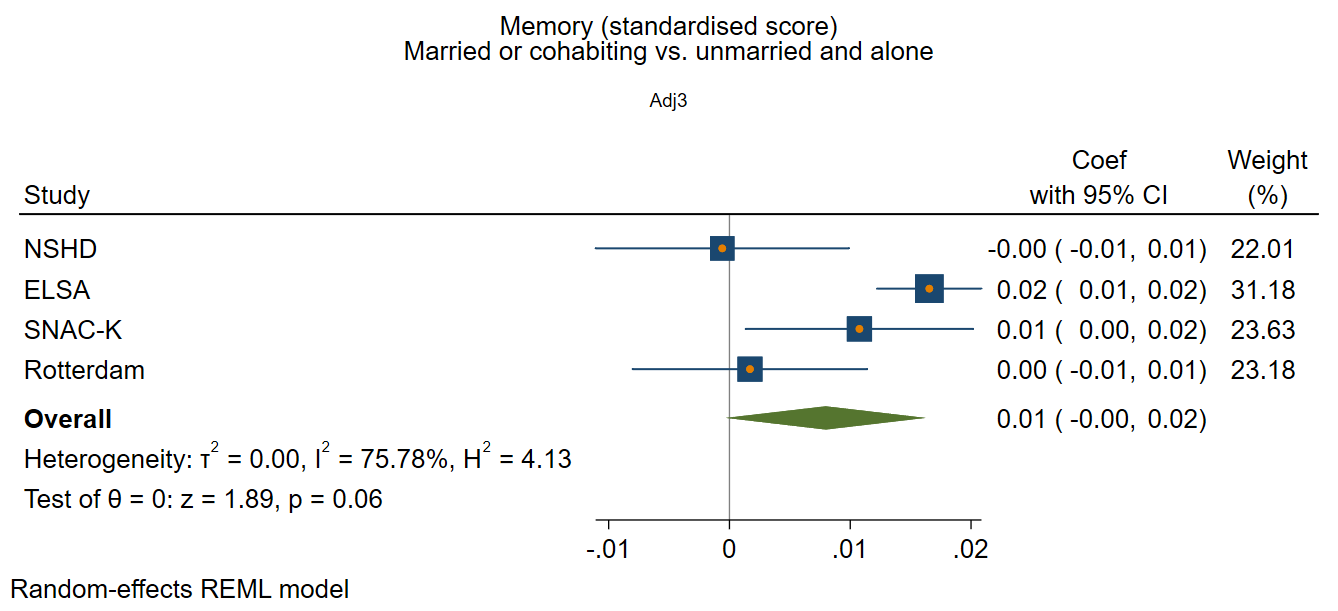

#### Network size

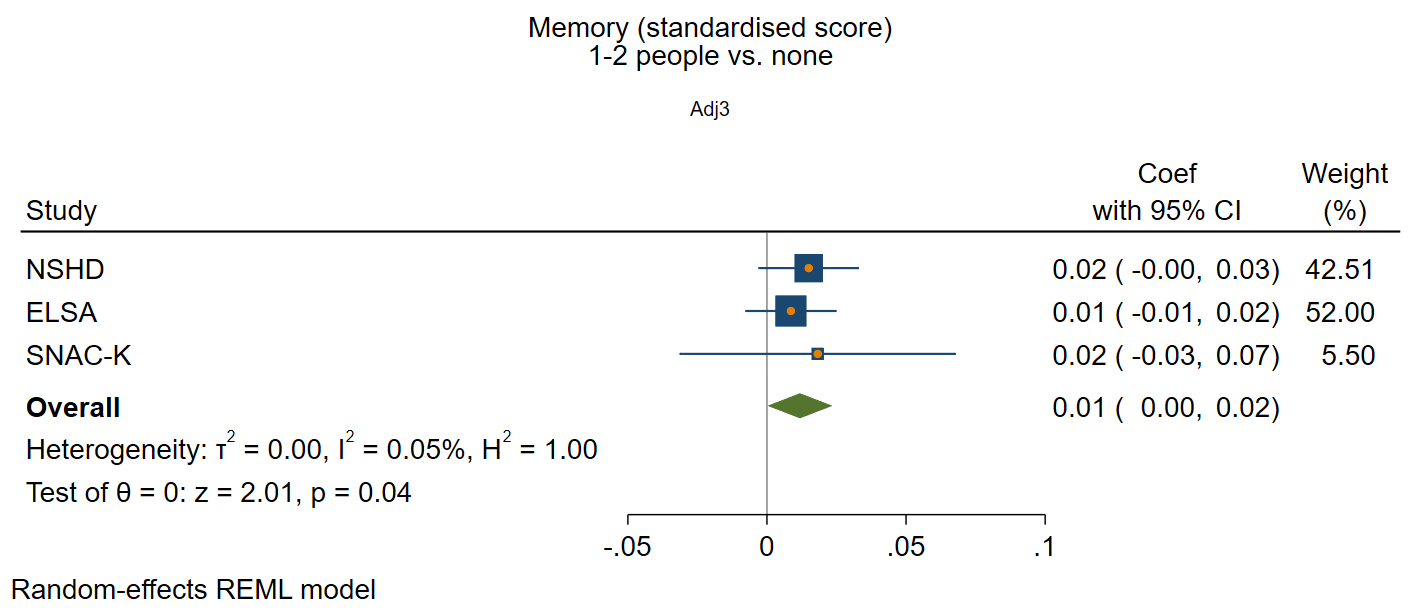

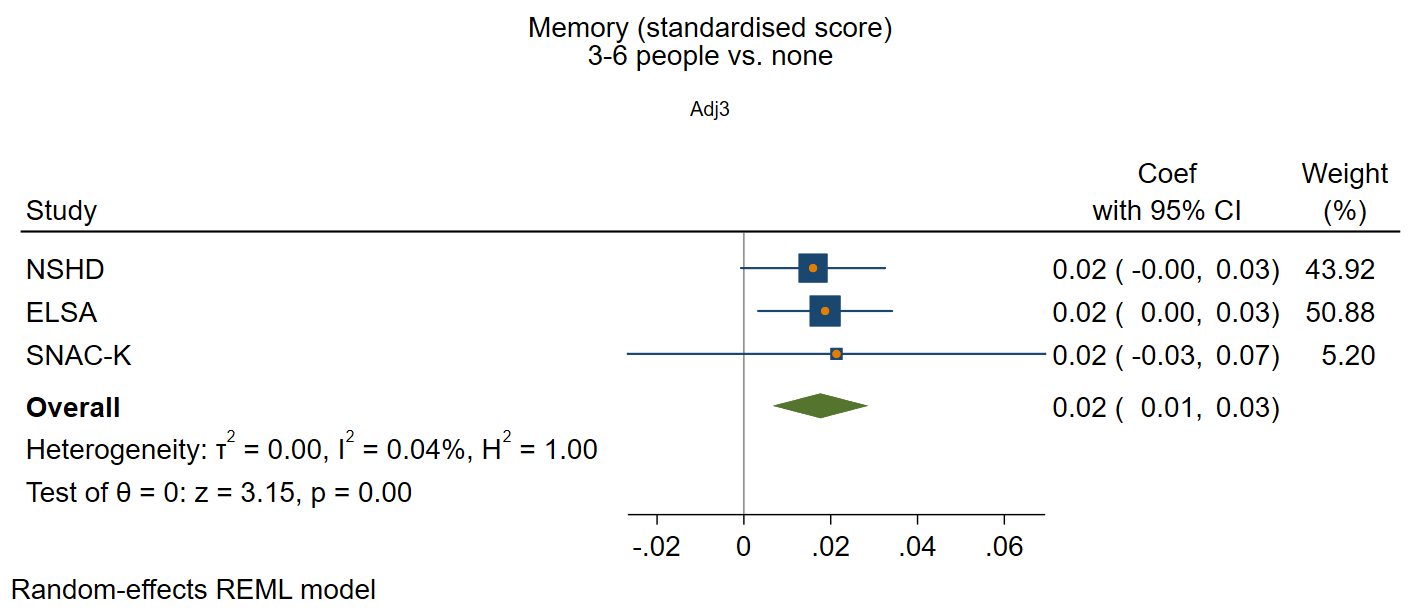

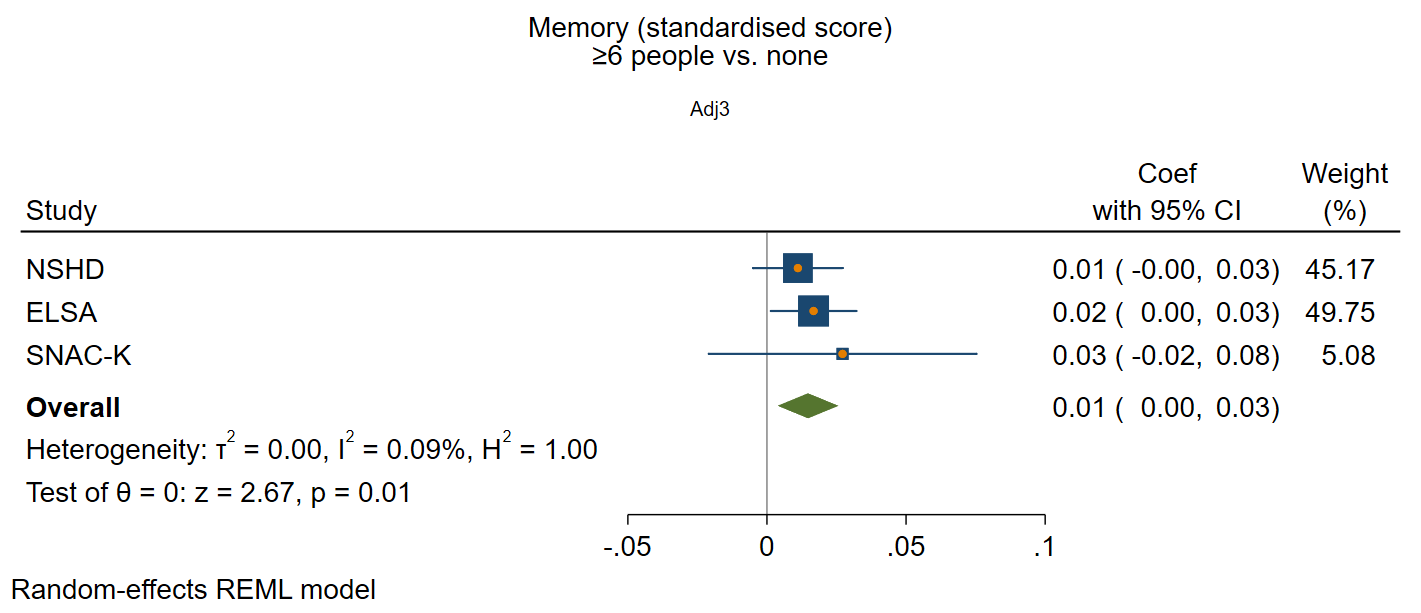

#### Frequency of contact

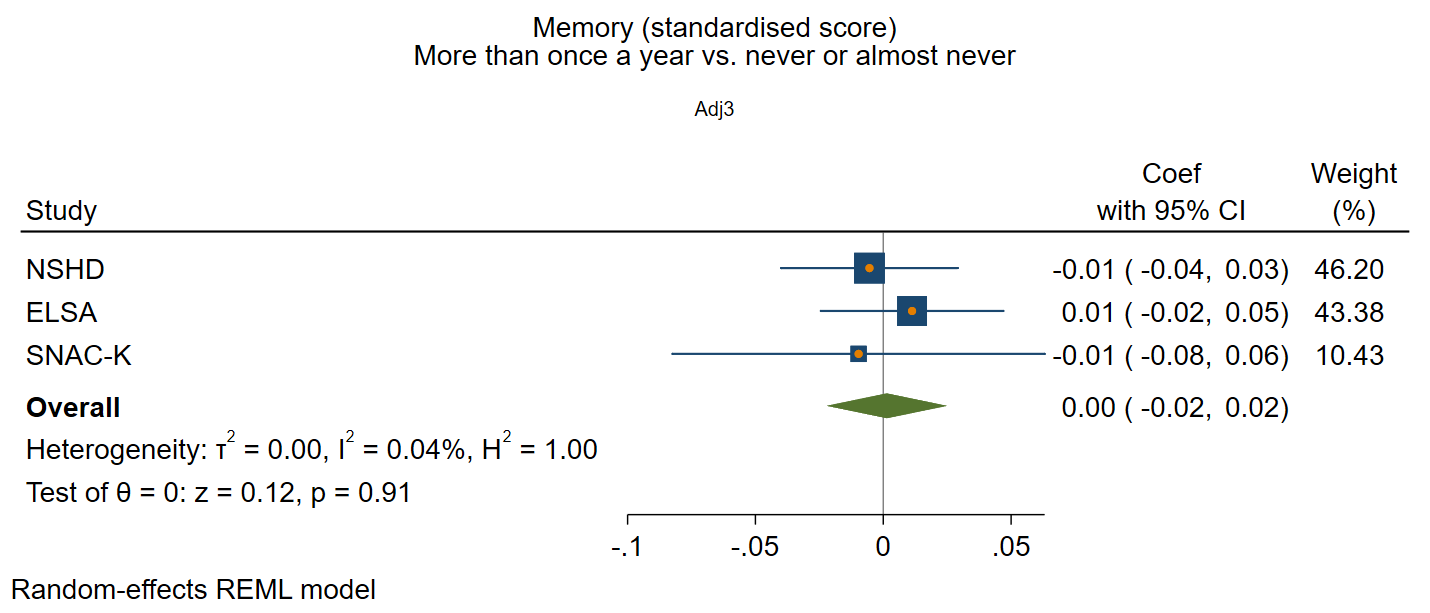

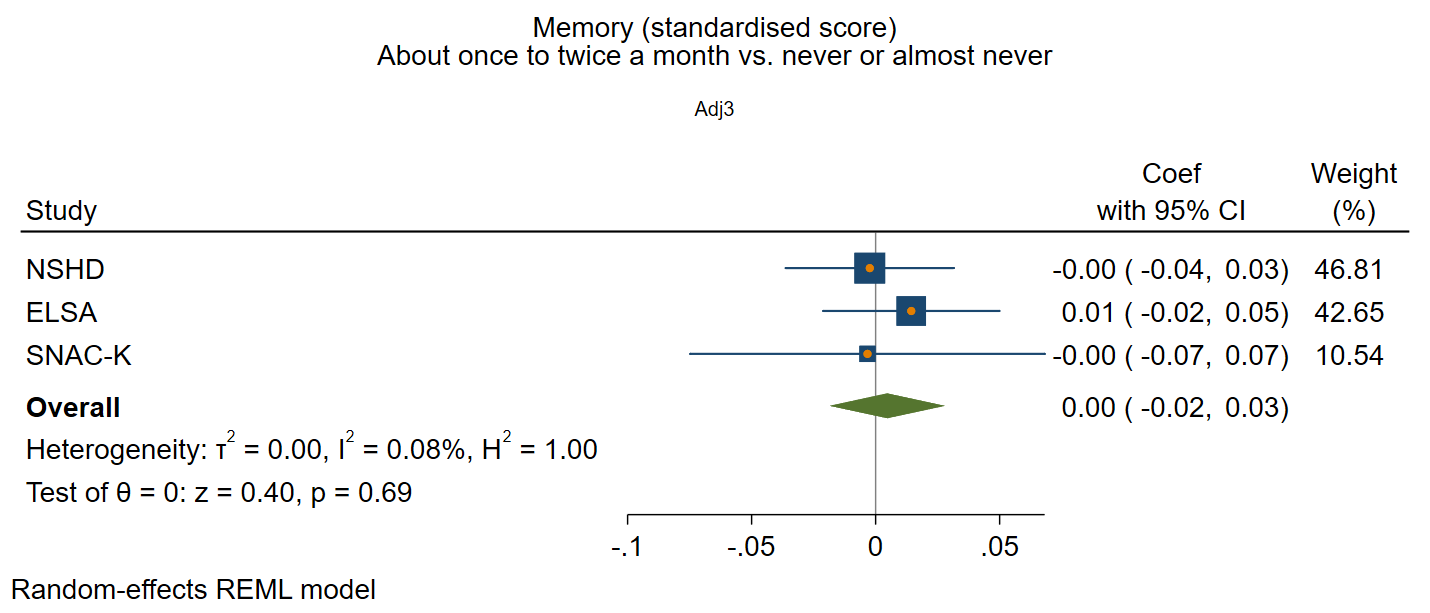

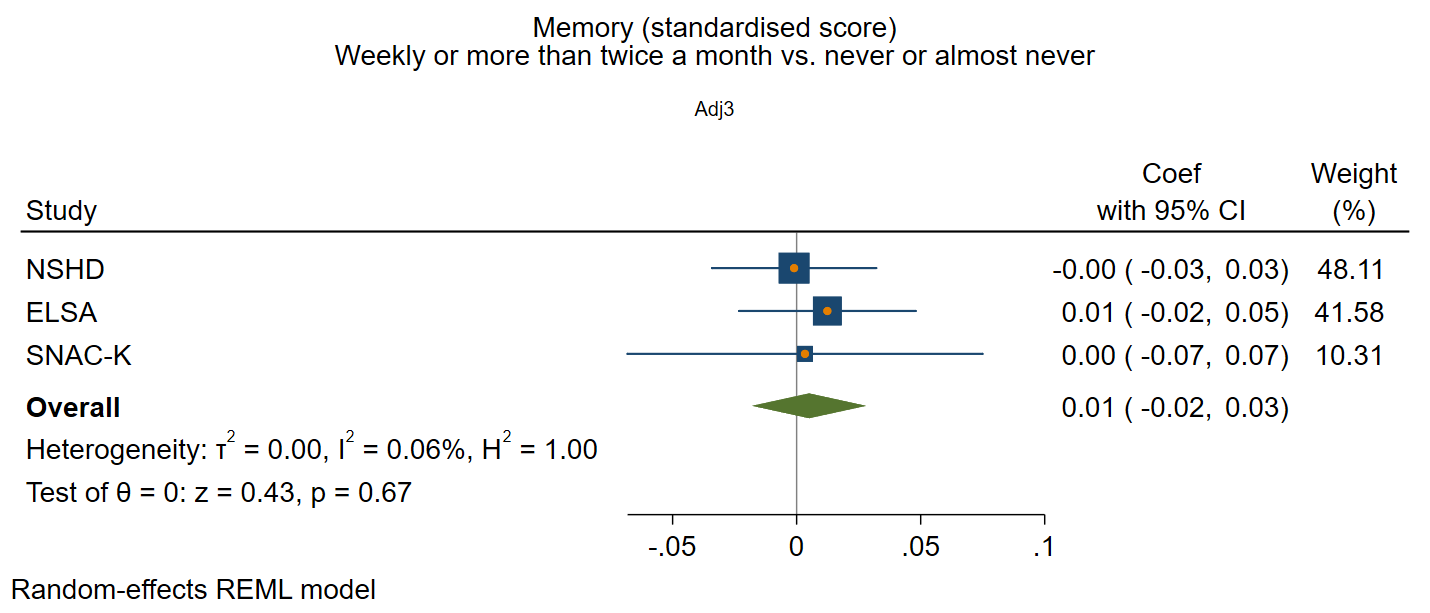

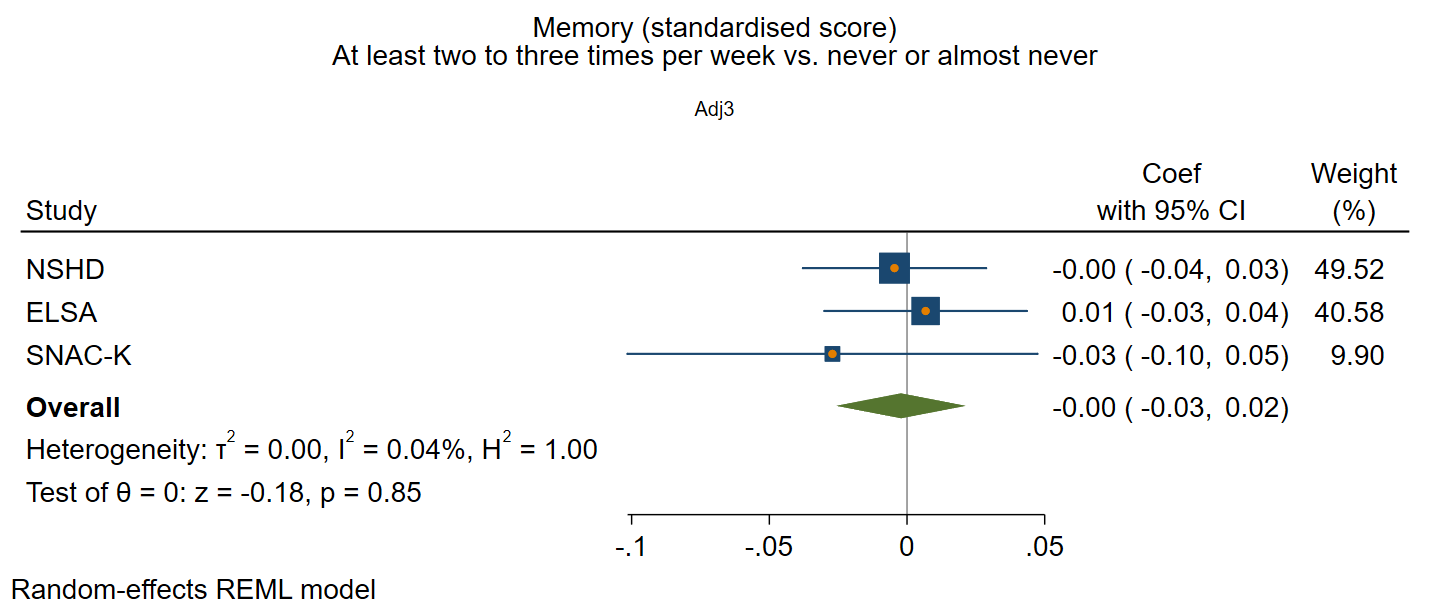

### Supplementary figure 3: Functional social health at baseline and rate of decline in memory

*Adjusted for sex and age at baseline, social class, education, IADL, and vascular-related health conditions*

#### Participation in social activities

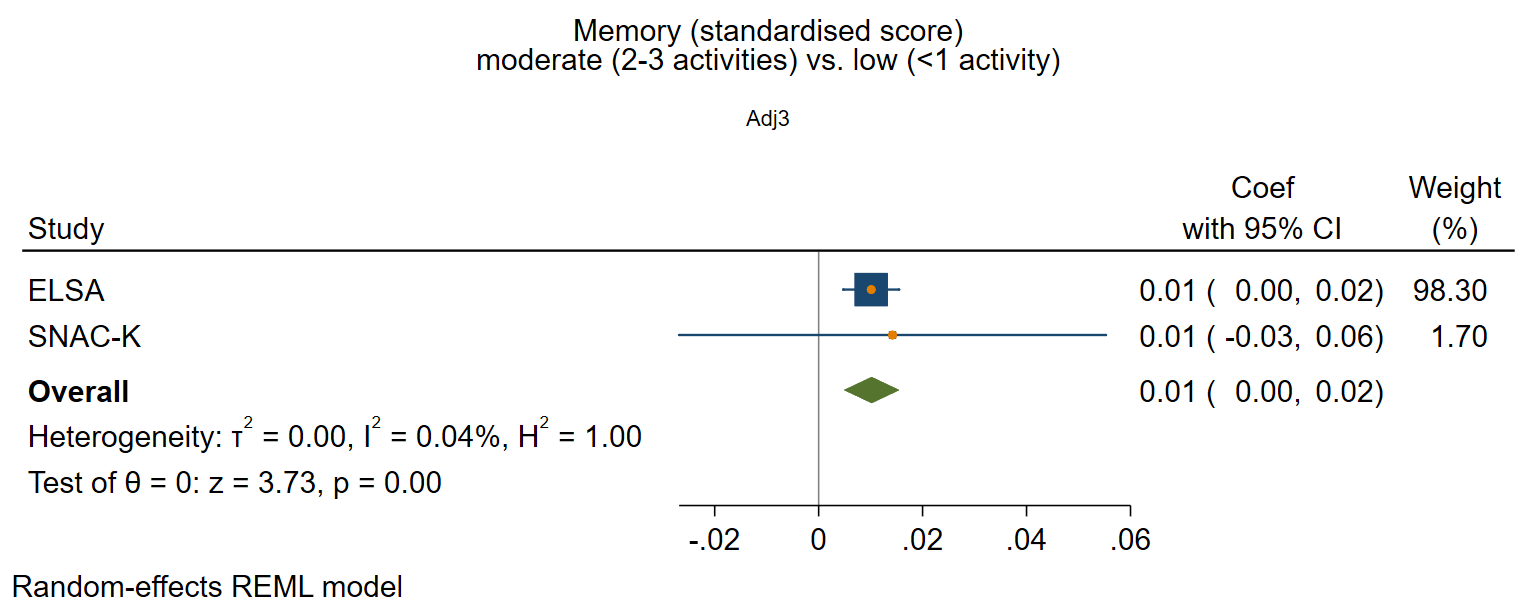

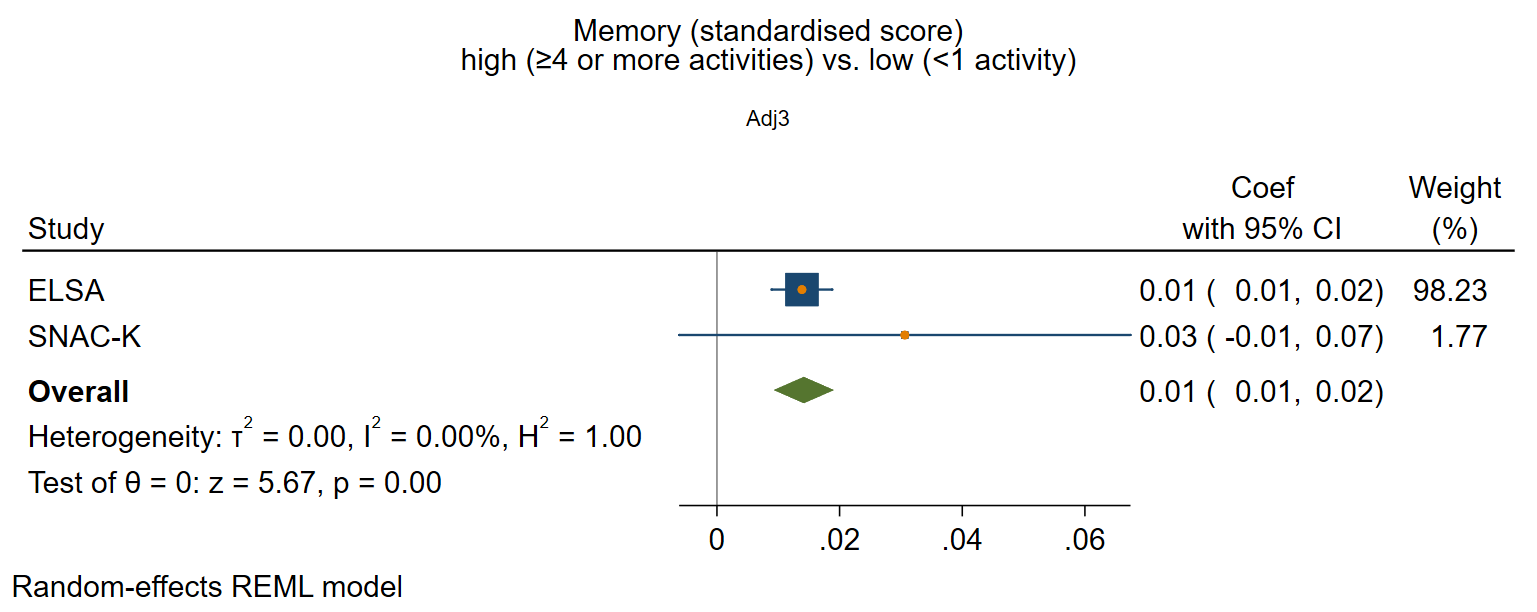

#### Perceived positive social support

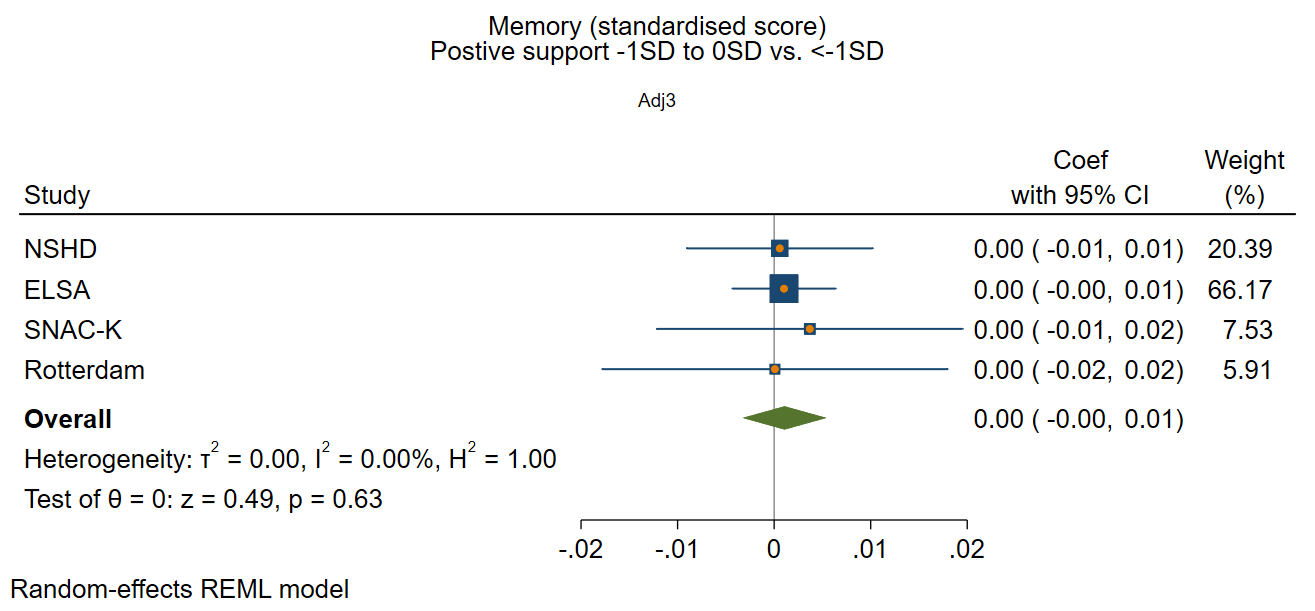

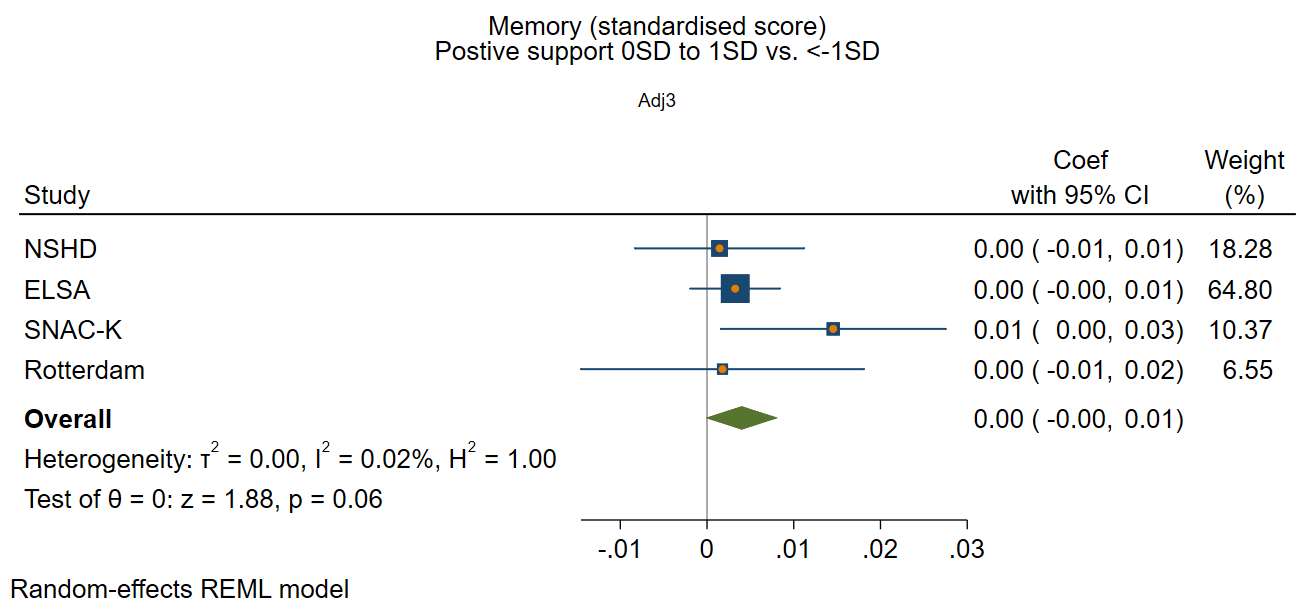

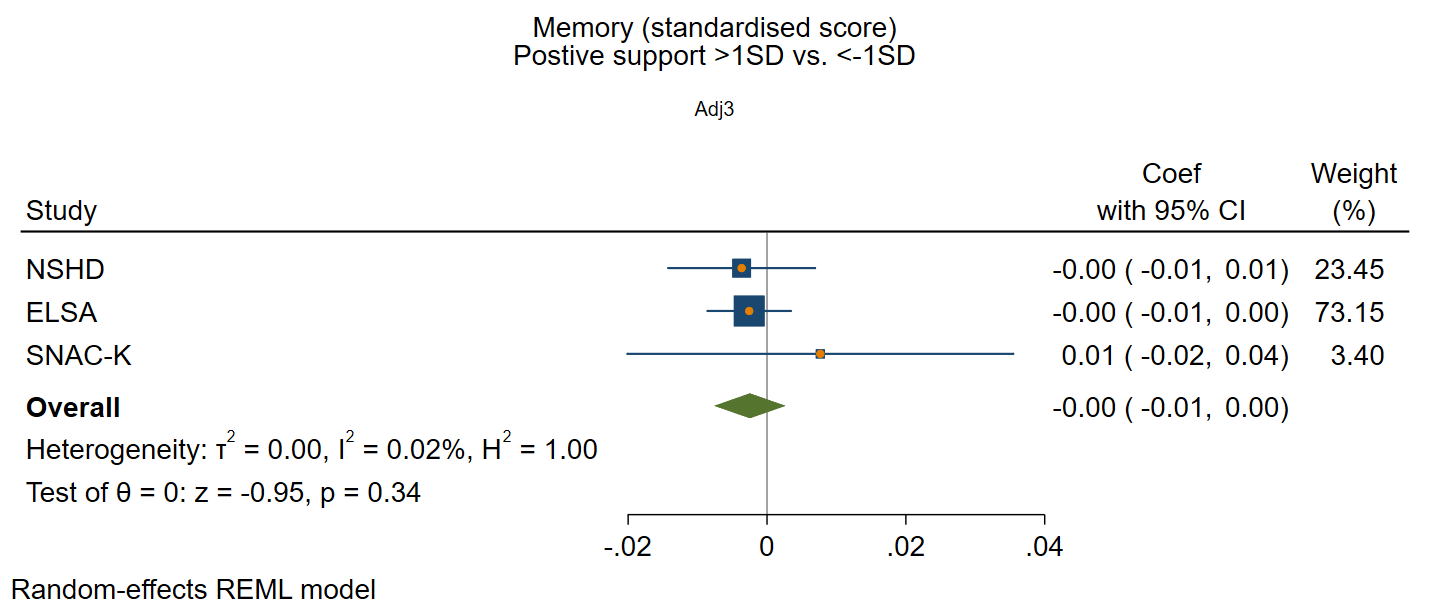

#### Perceive negative social support

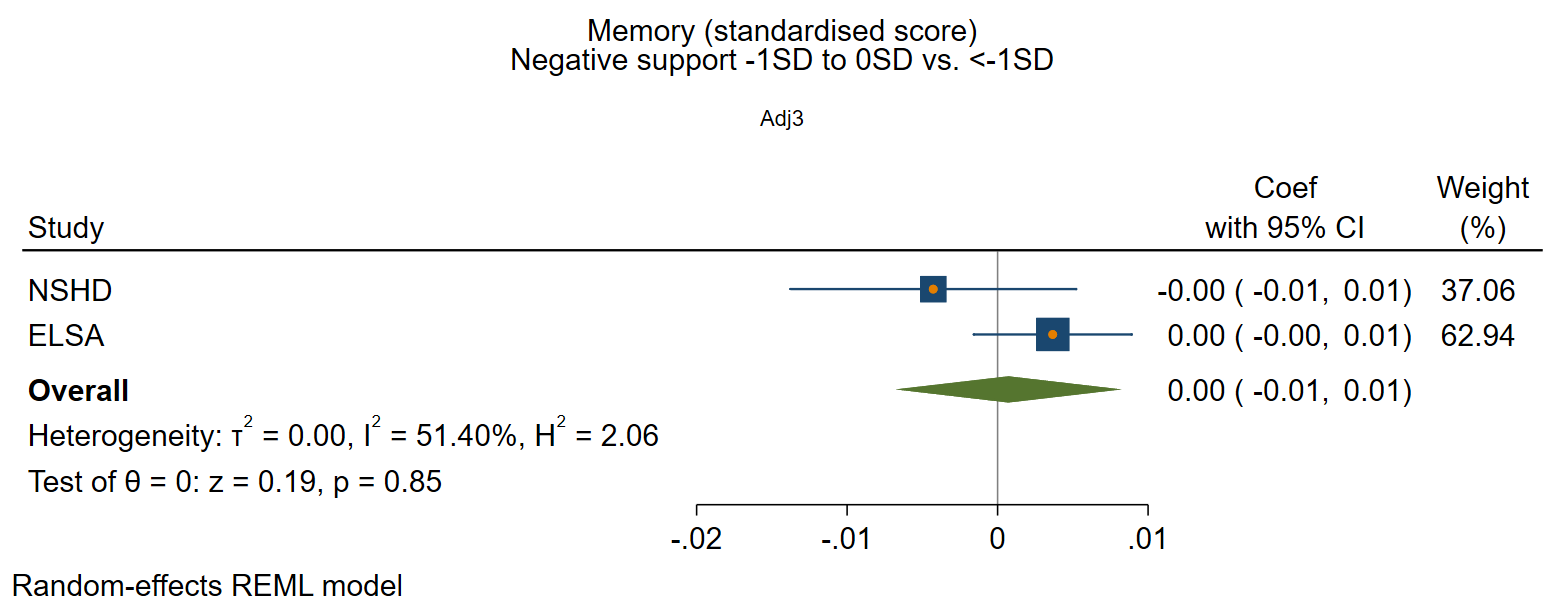

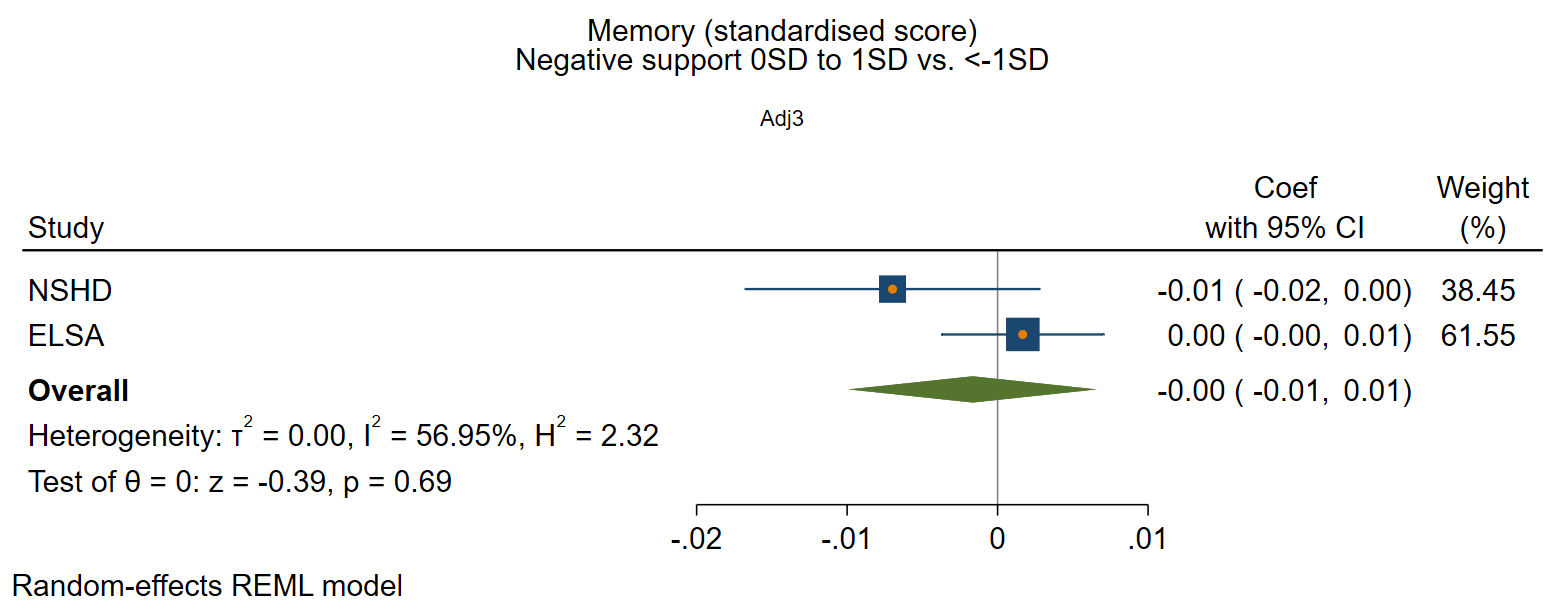

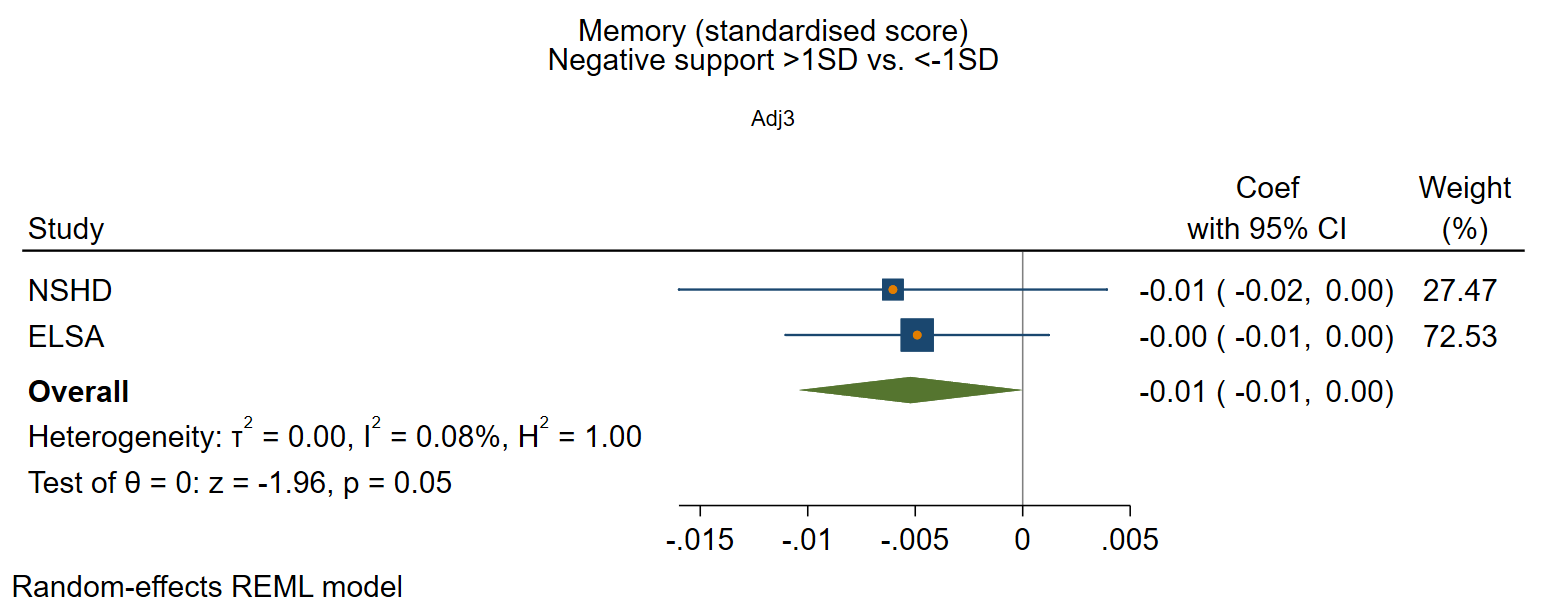

### Supplementary figure 4: Structural social health at baseline and rate of decline in executive function

*Adjusted for sex and age at baseline, social class, education, IADL, and vascular-related health conditions.*

#### Marital/cohabitation status

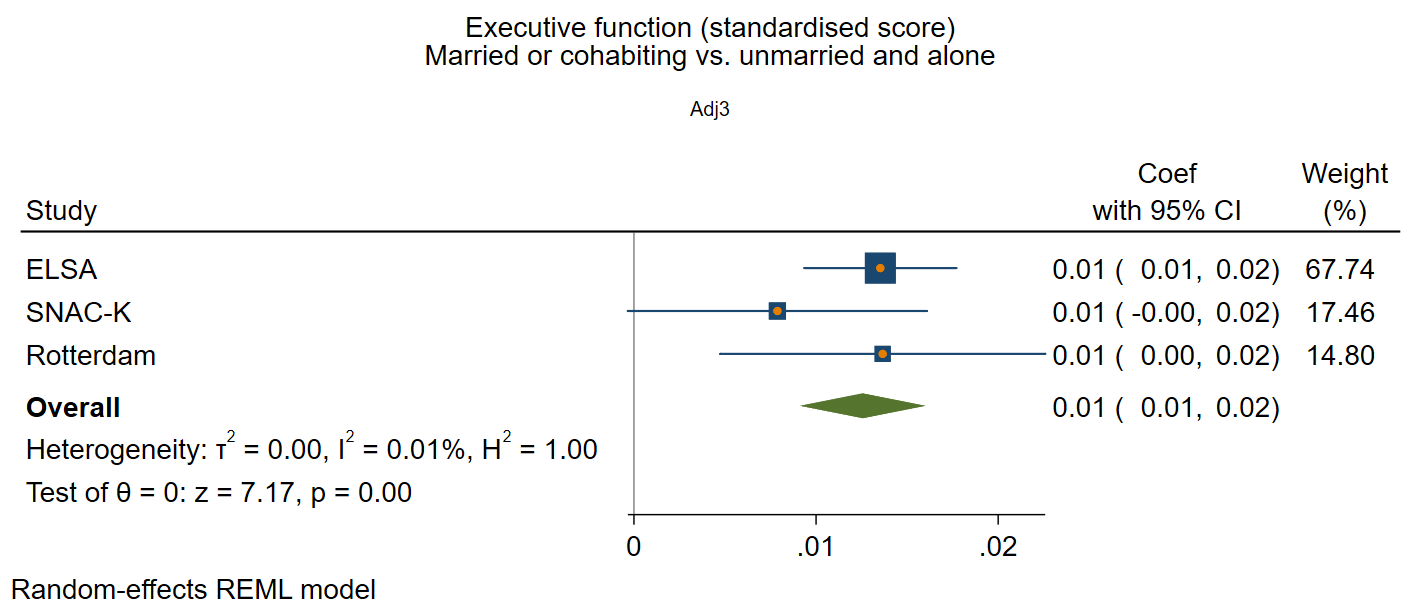

#### Network size

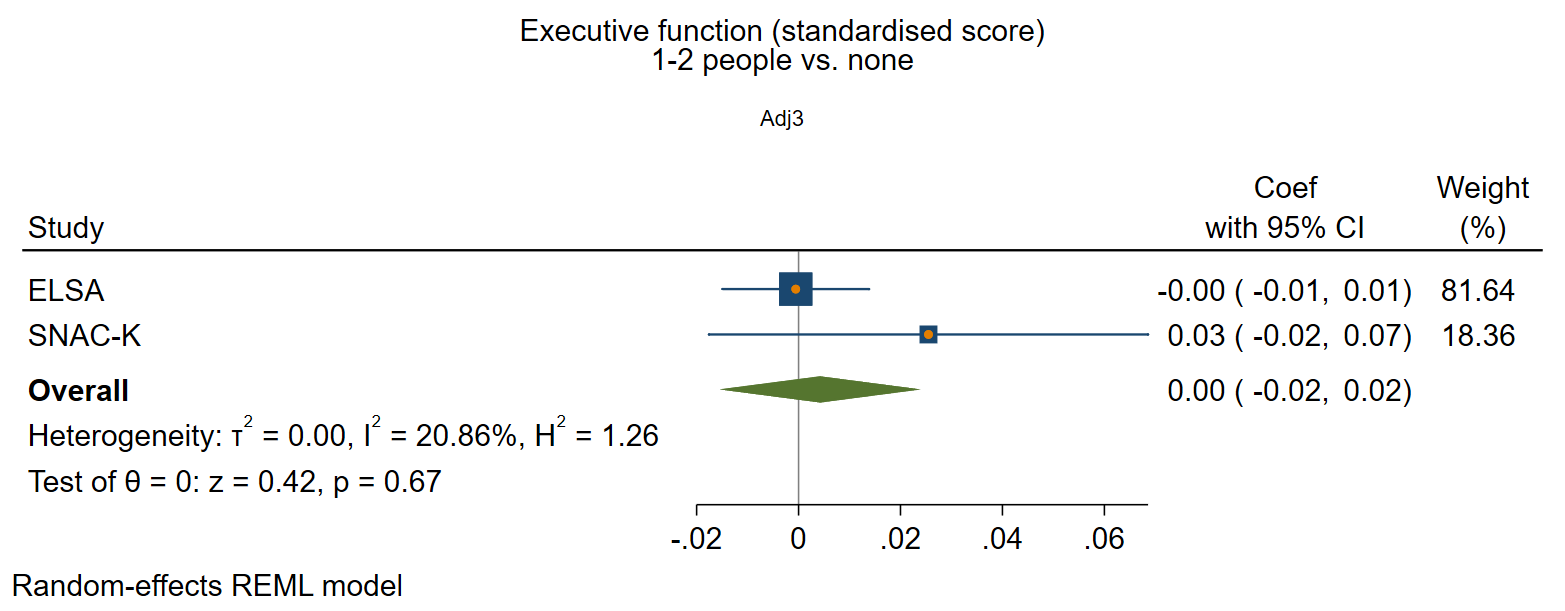

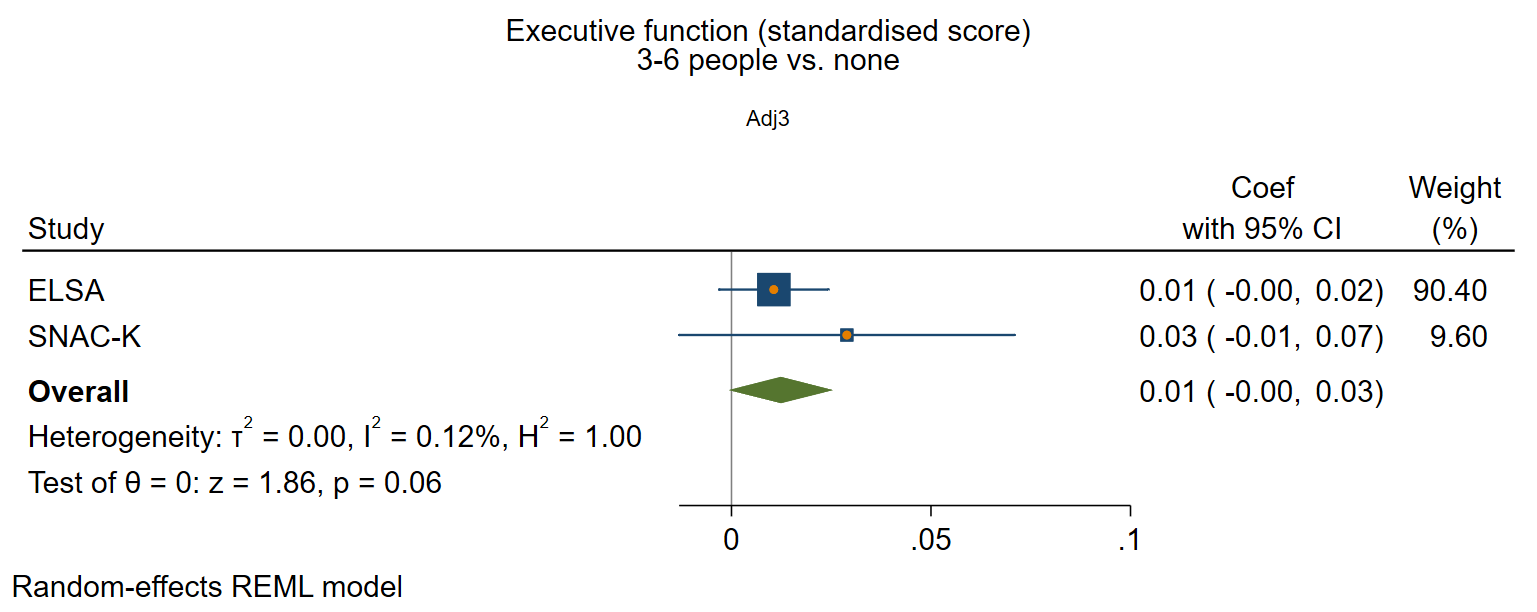

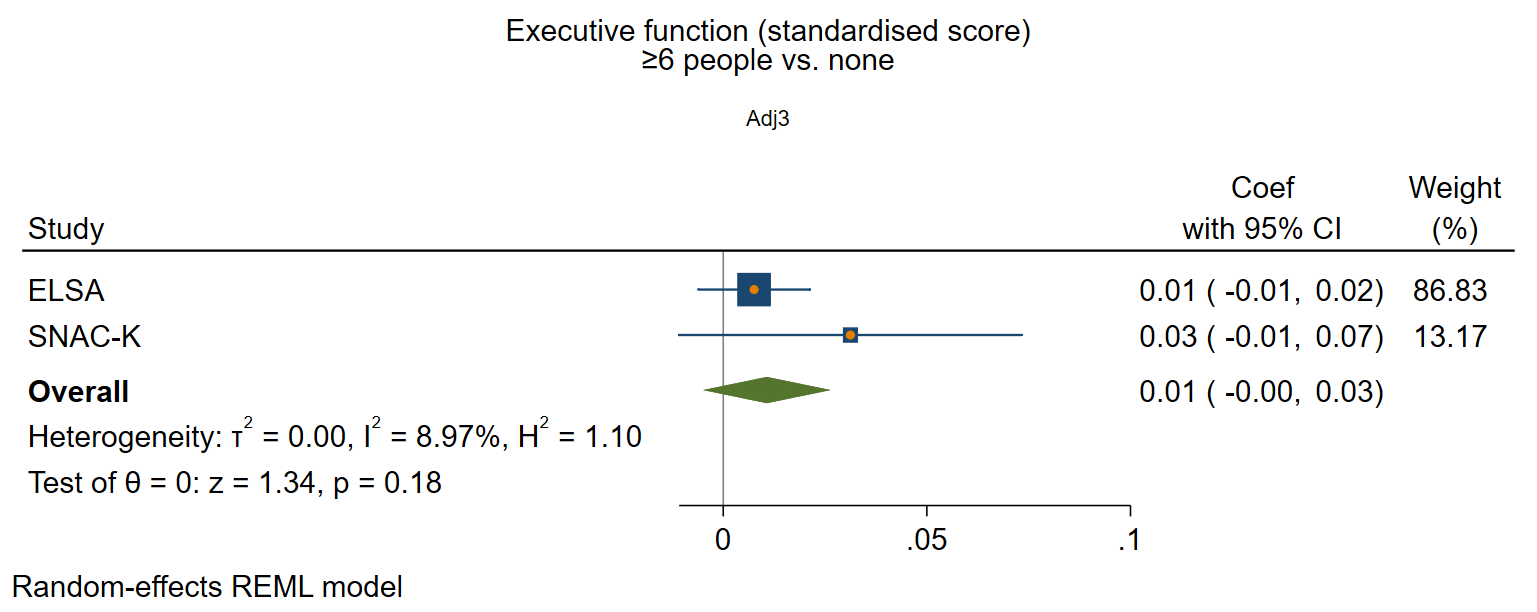

#### Frequency of contact

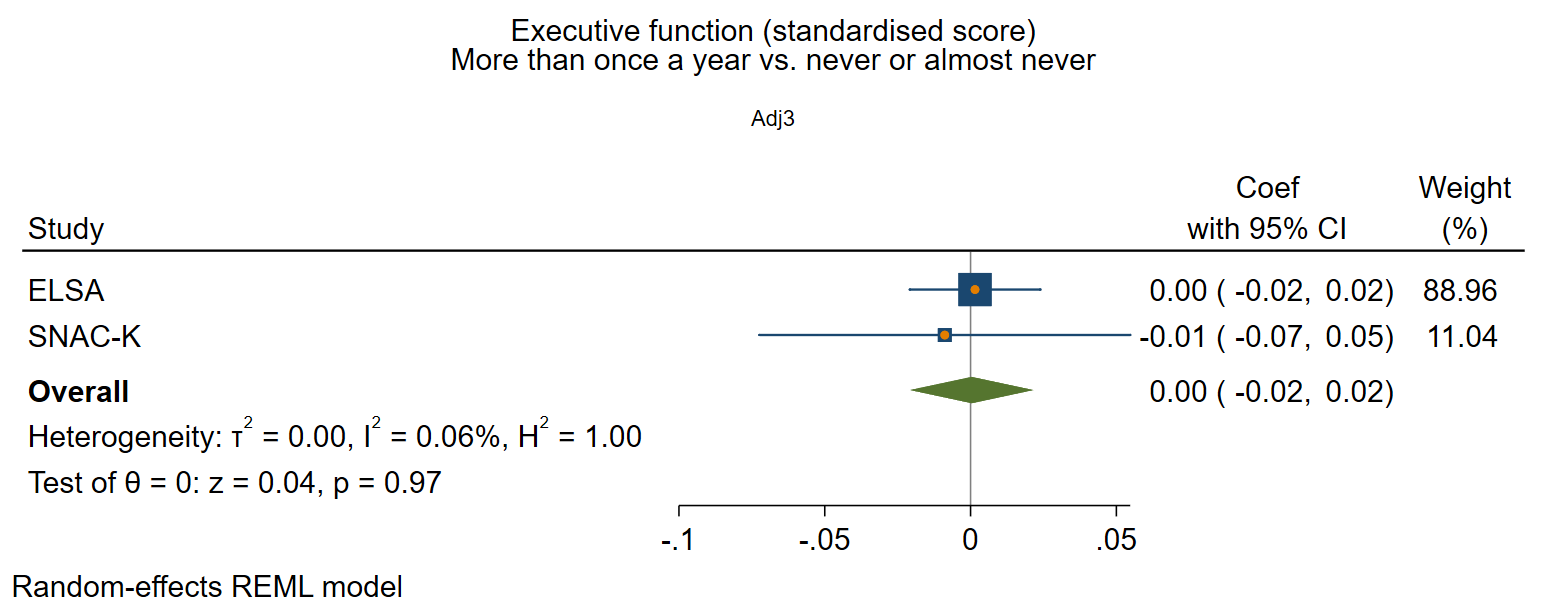

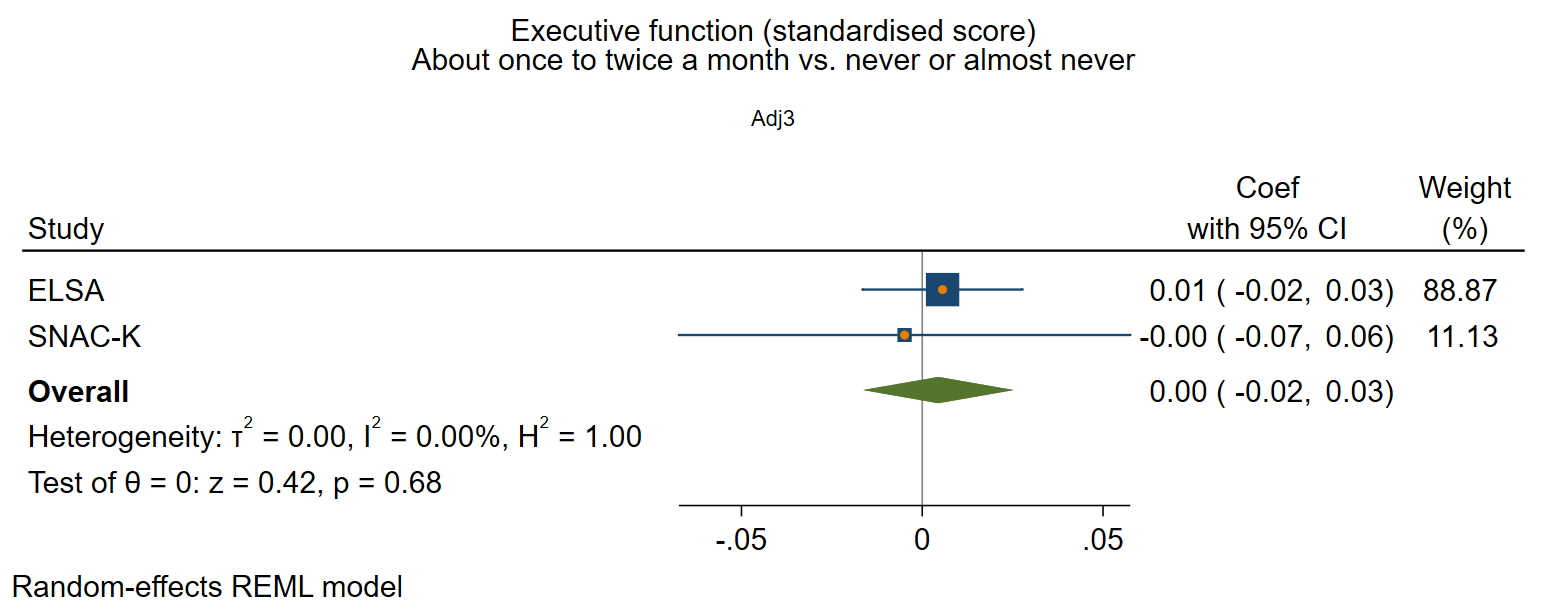

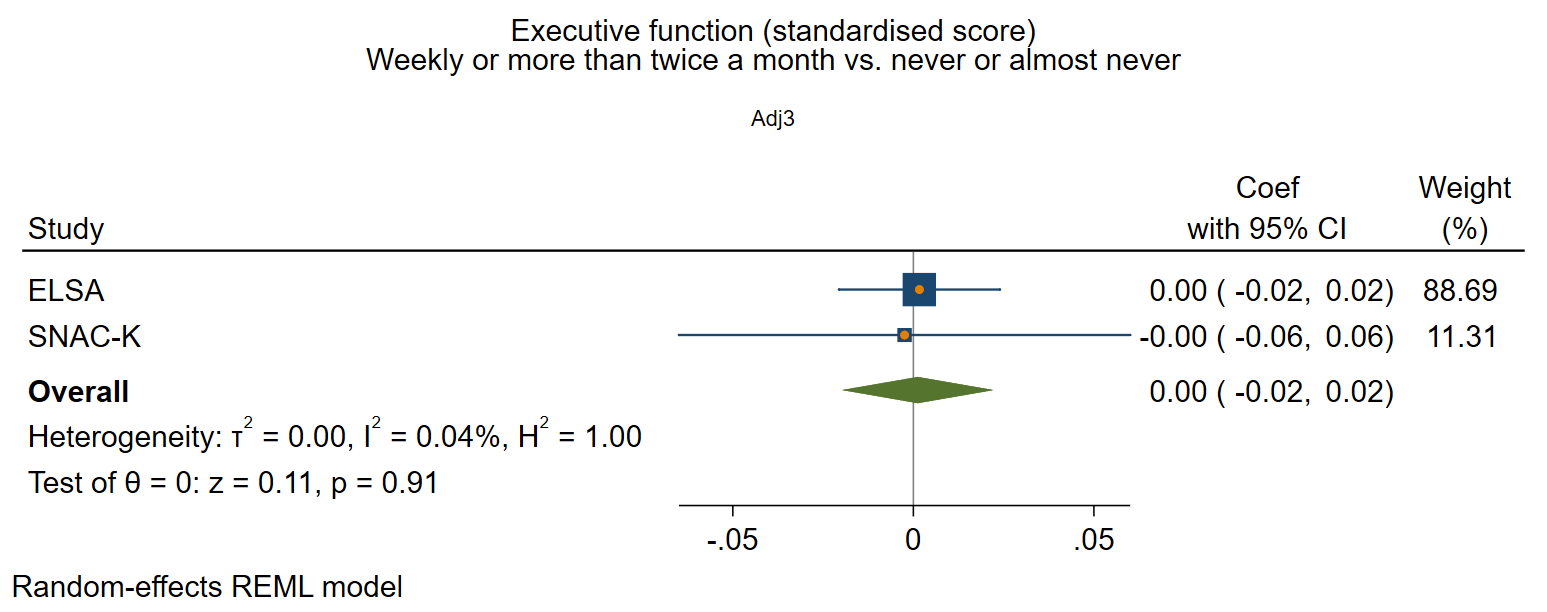

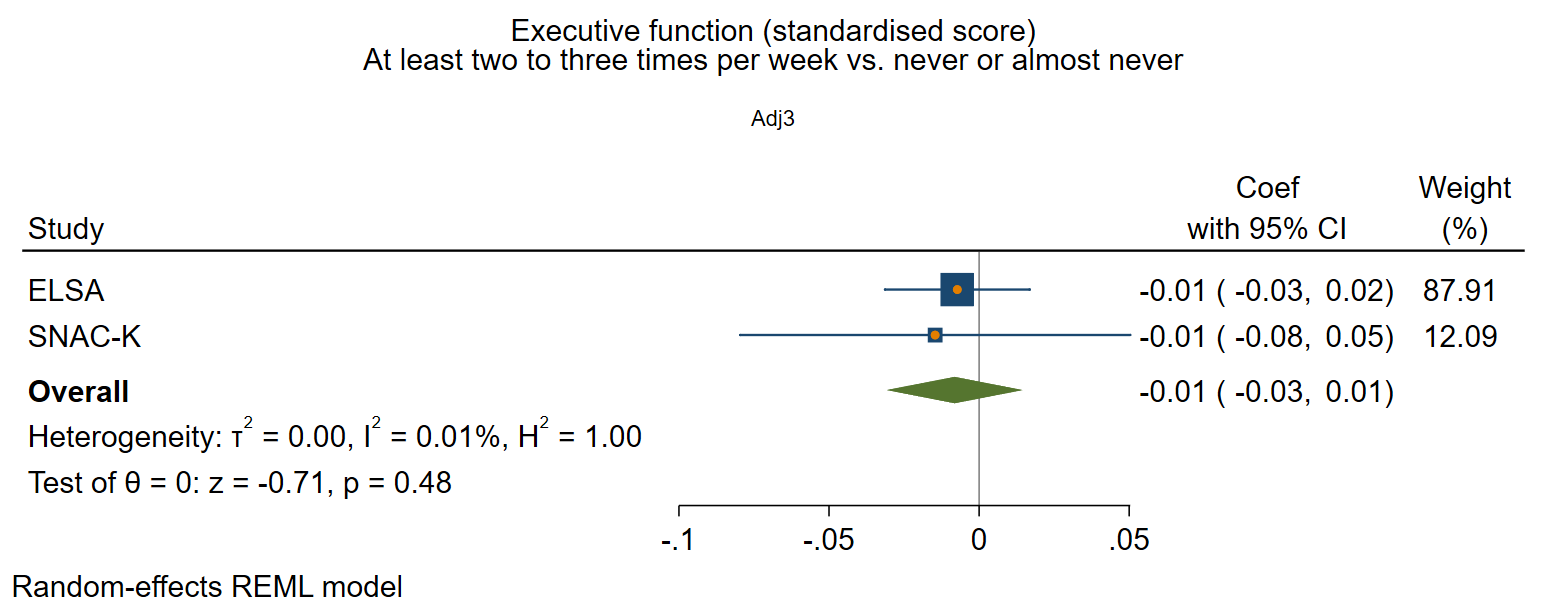

### Supplementary figure 5: Functional social health at baseline and rate of decline in executive function

*Adjusted for sex and age at baseline, social class, education, IADL, and vascular-related health conditions.*

#### Participation in social activities

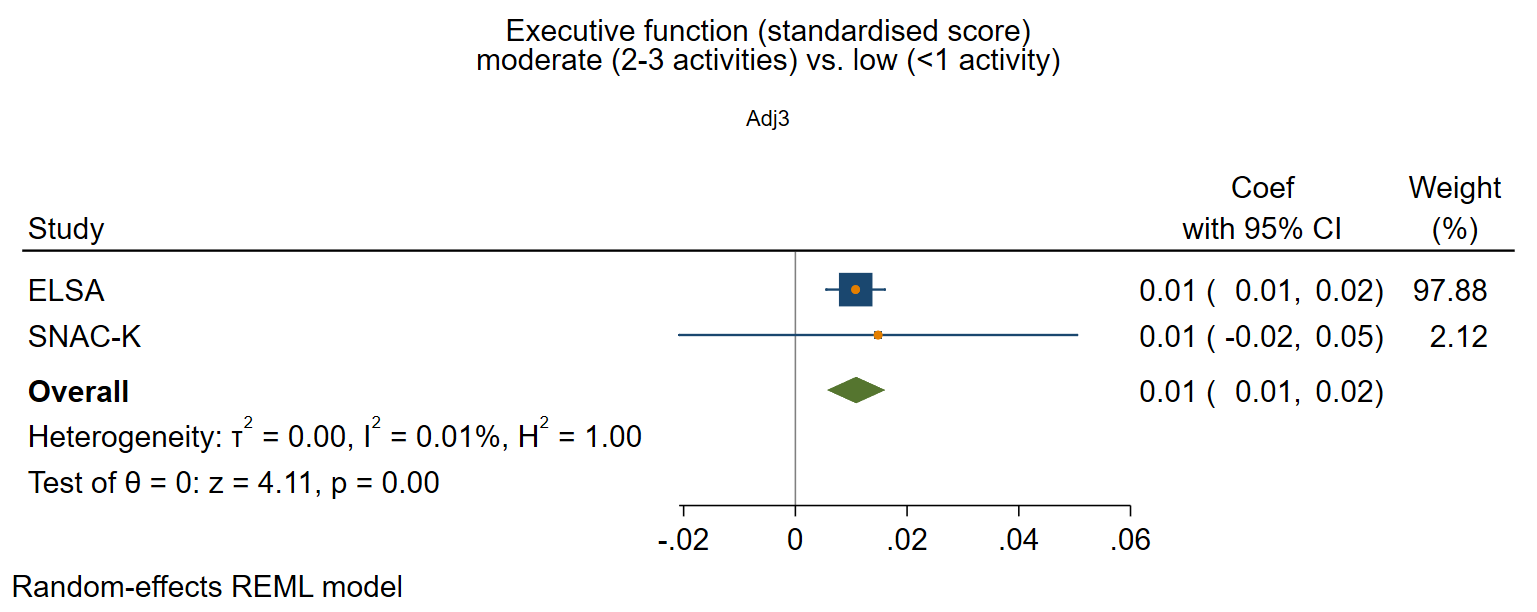

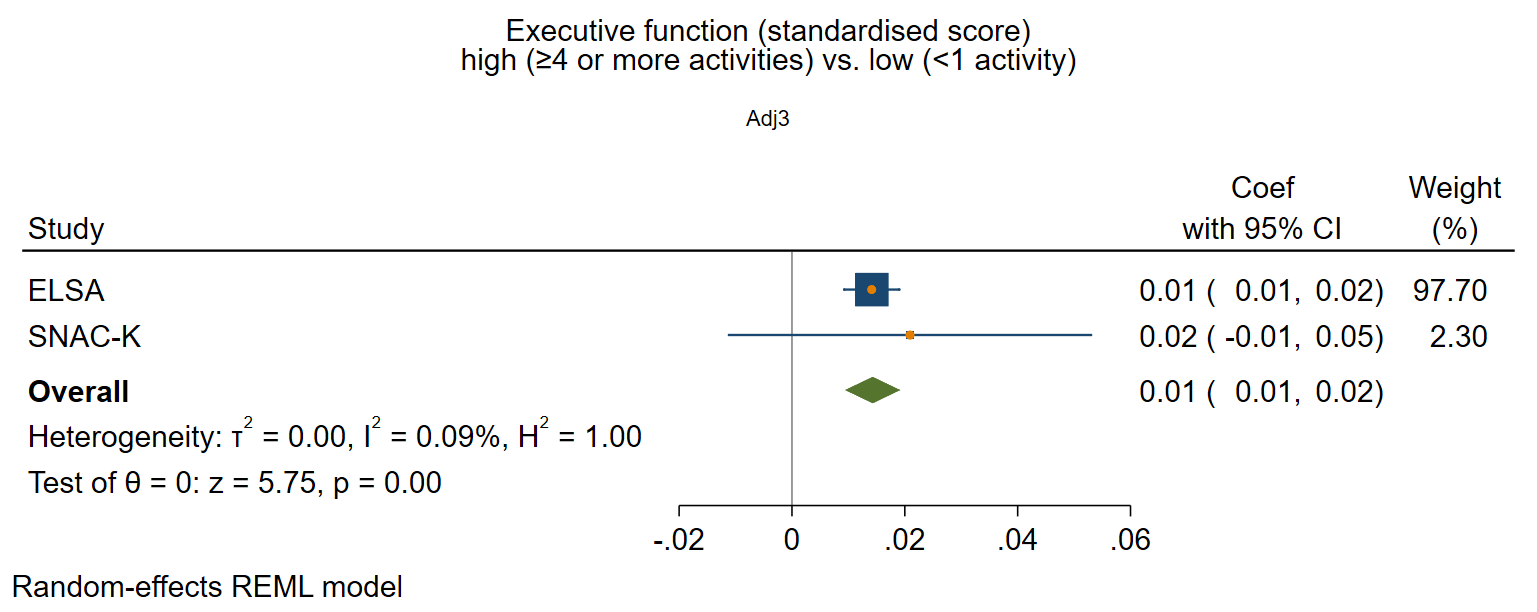

#### Perceived positive social support

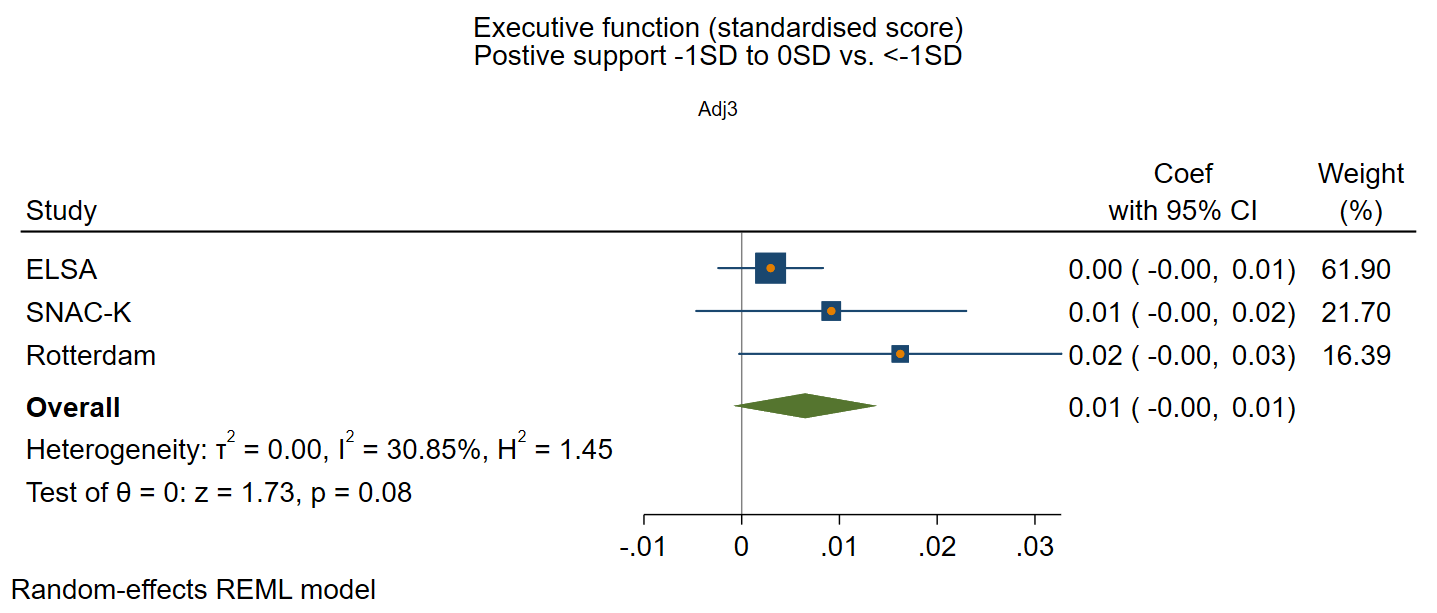

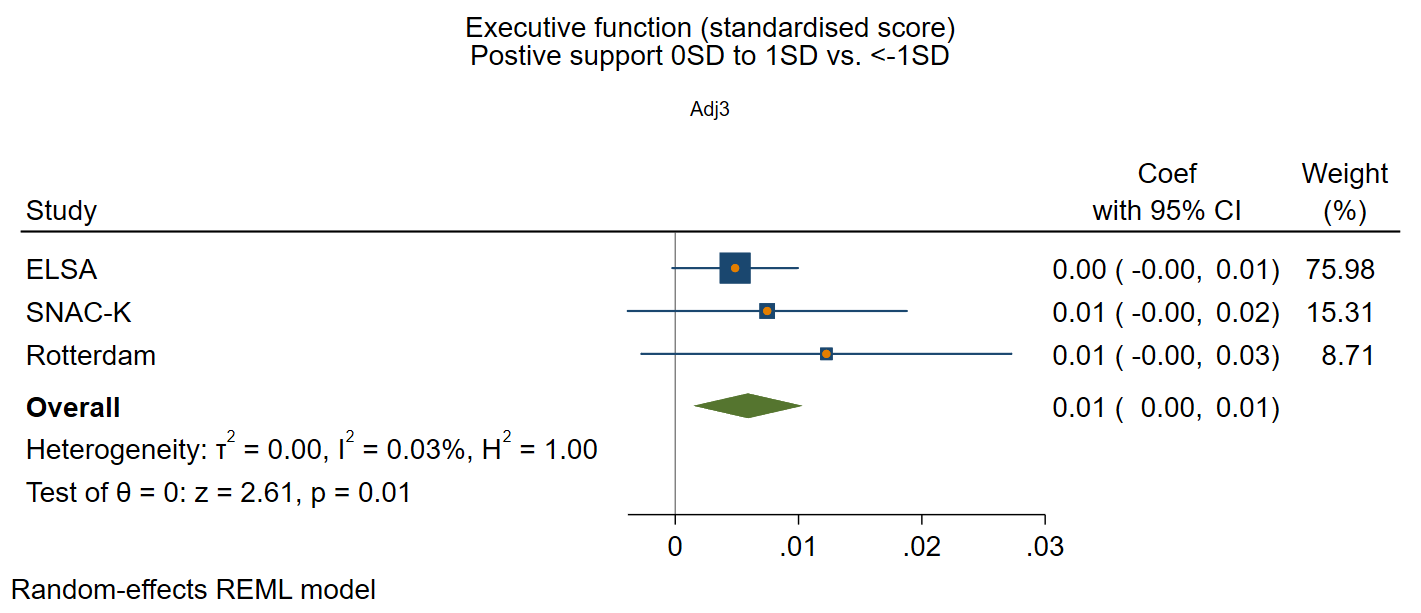

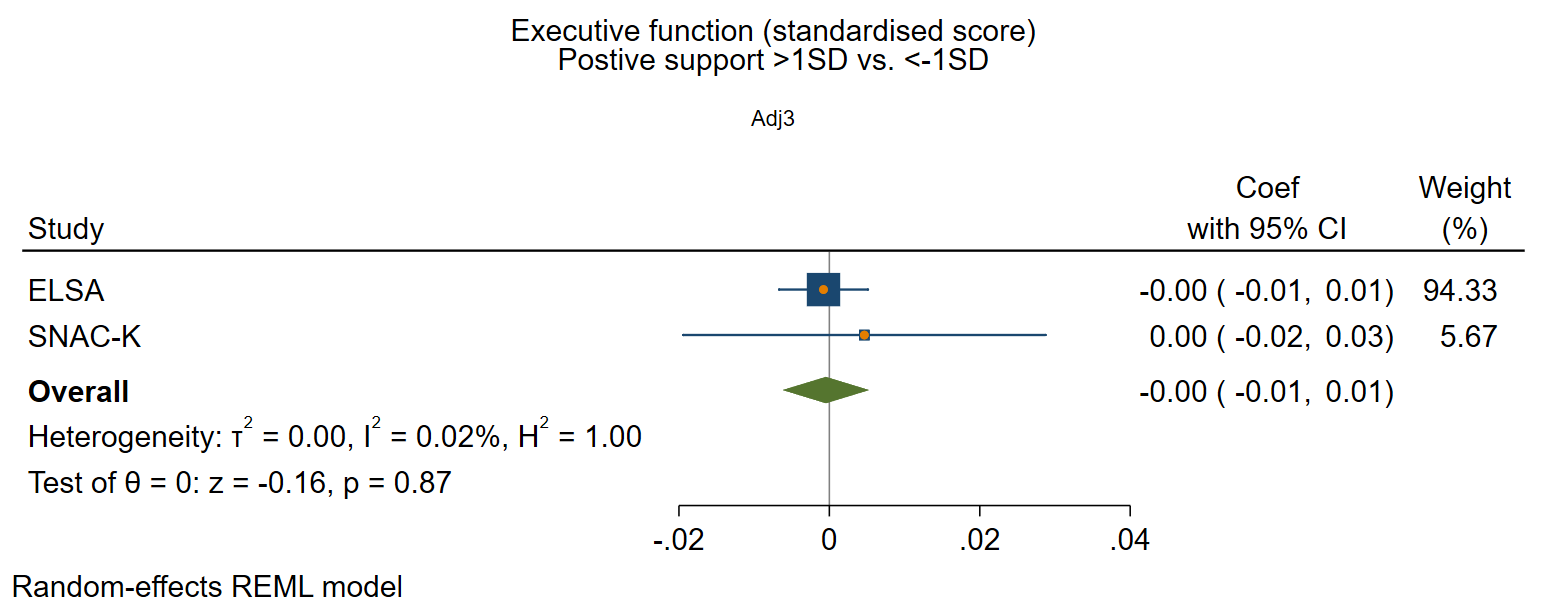

#### Perceived negative social support

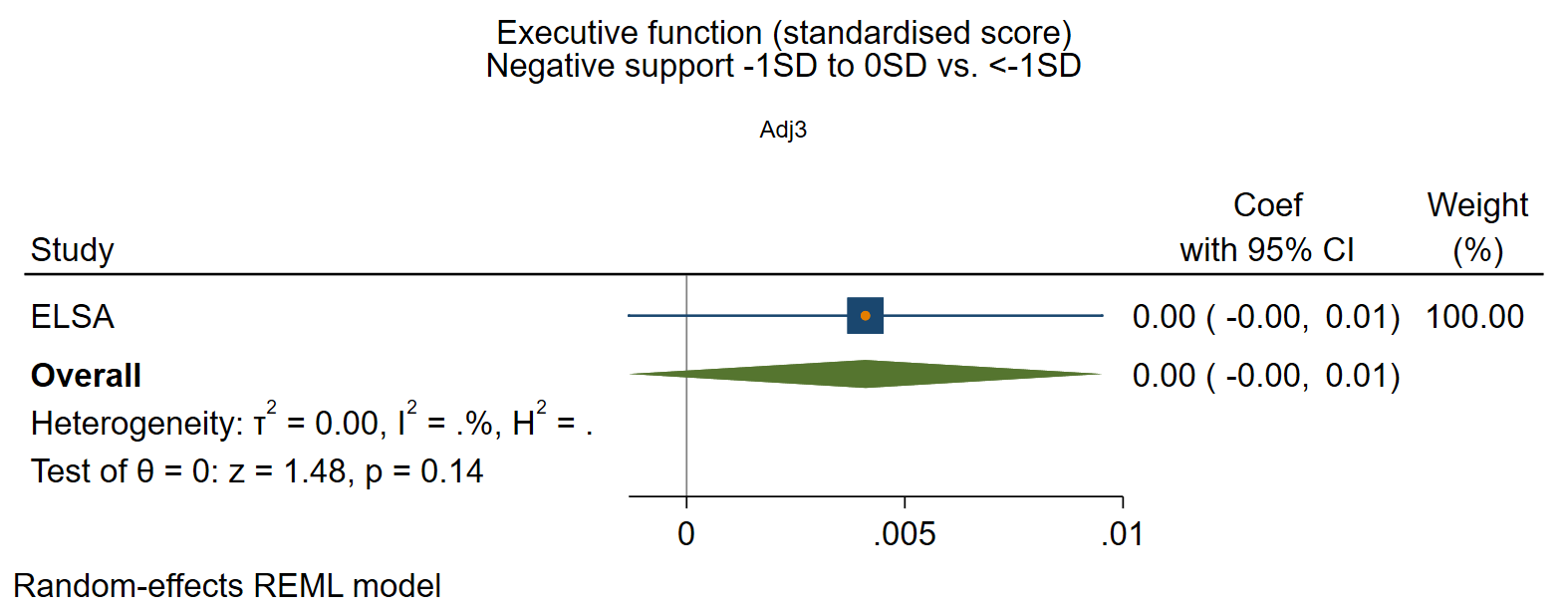

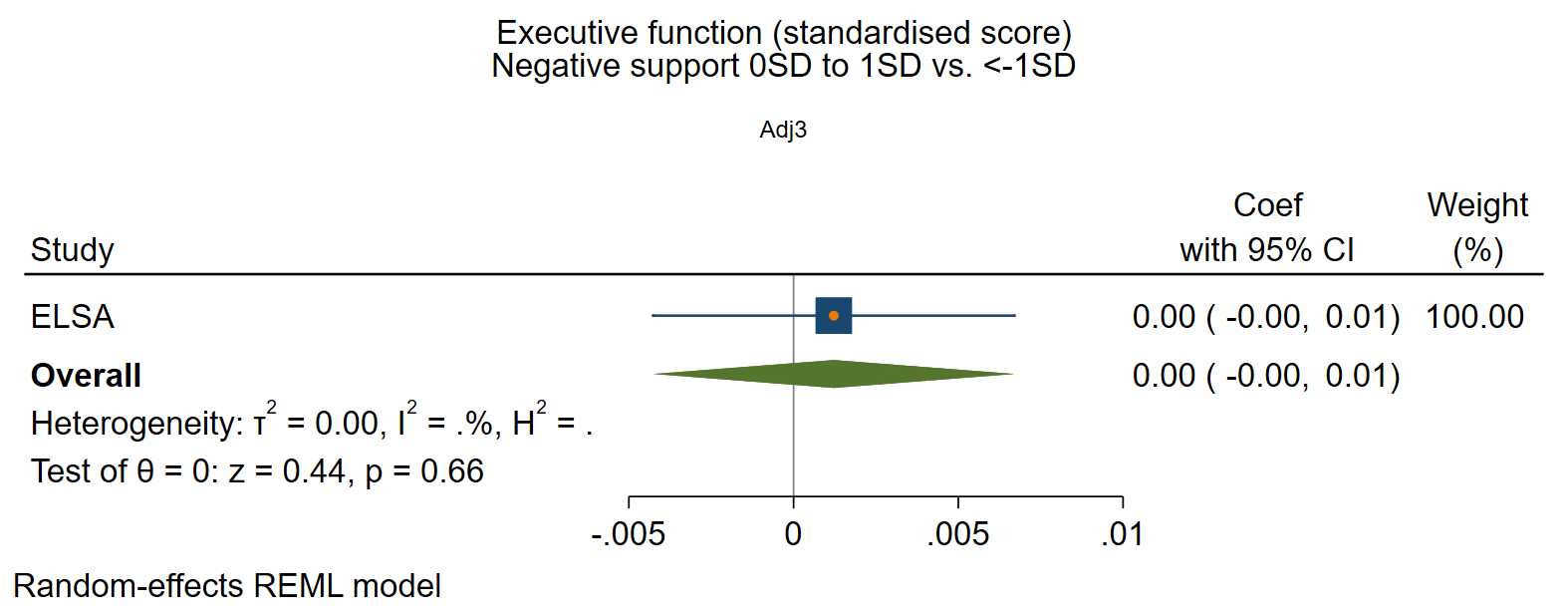

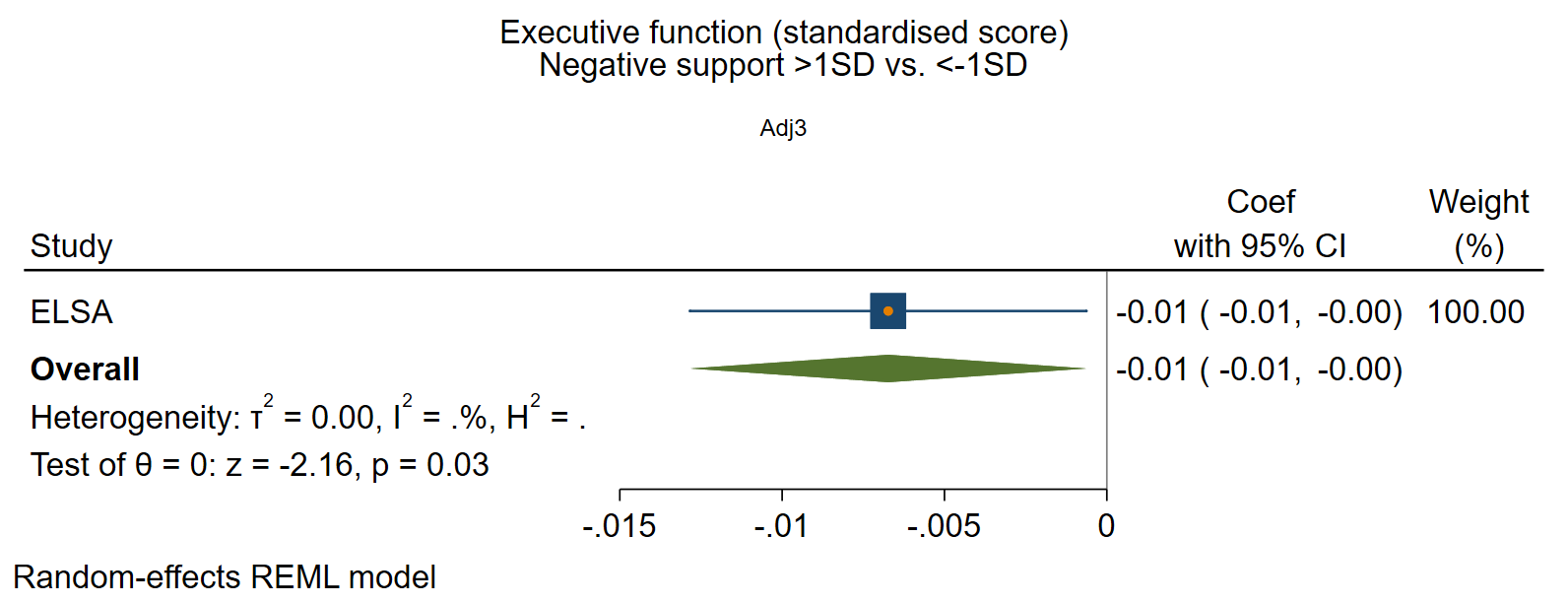

### Supplementary figure 6: Structural social health at baseline and rate of decline in processing speed

*Adjusted for sex and age at baseline, social class, education, IADL, and vascular-related health conditions.*

#### Marital/cohabitation status

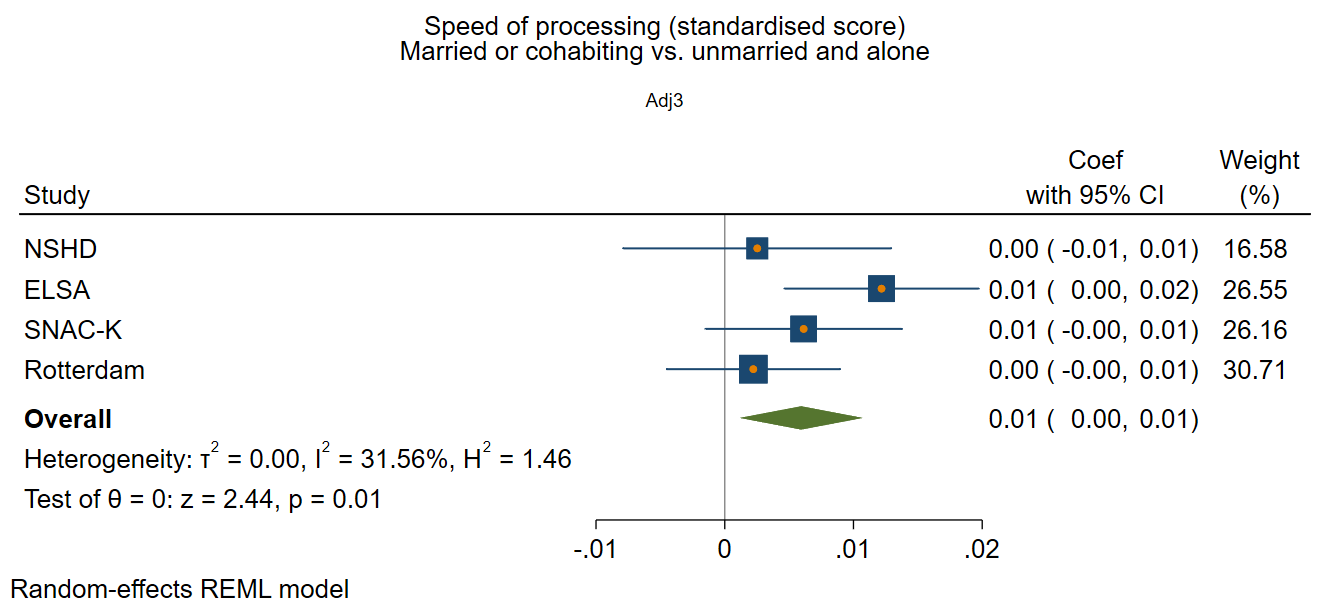

#### Network size

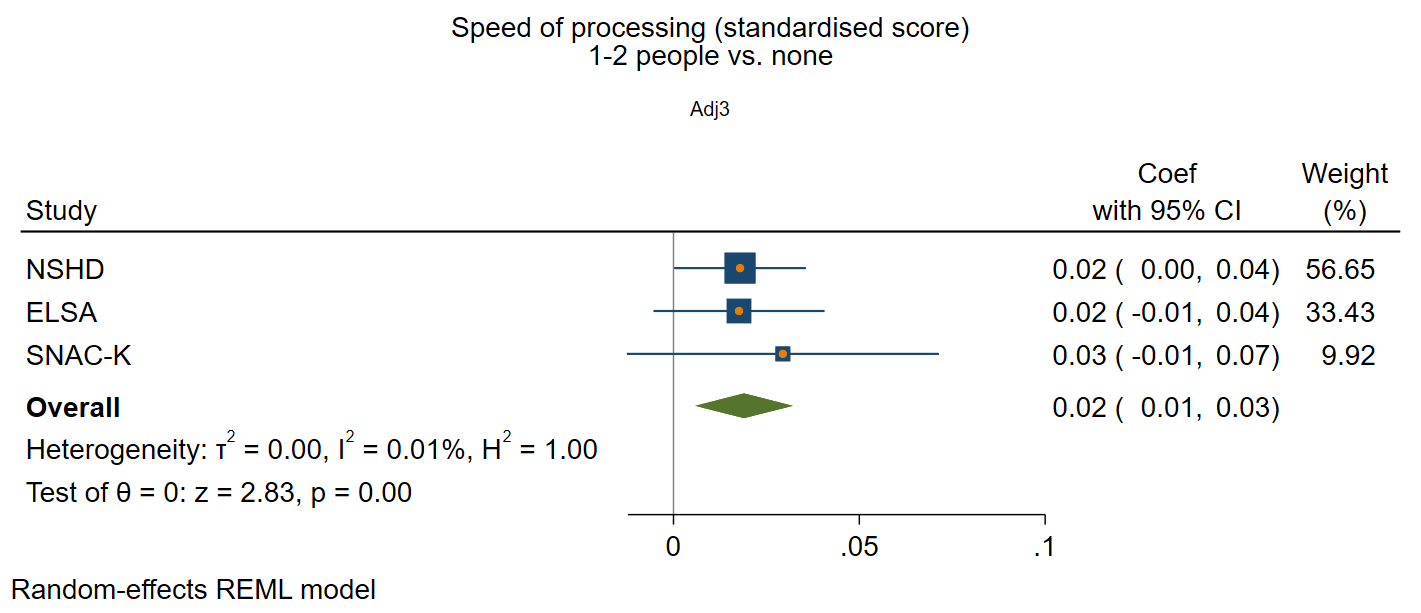

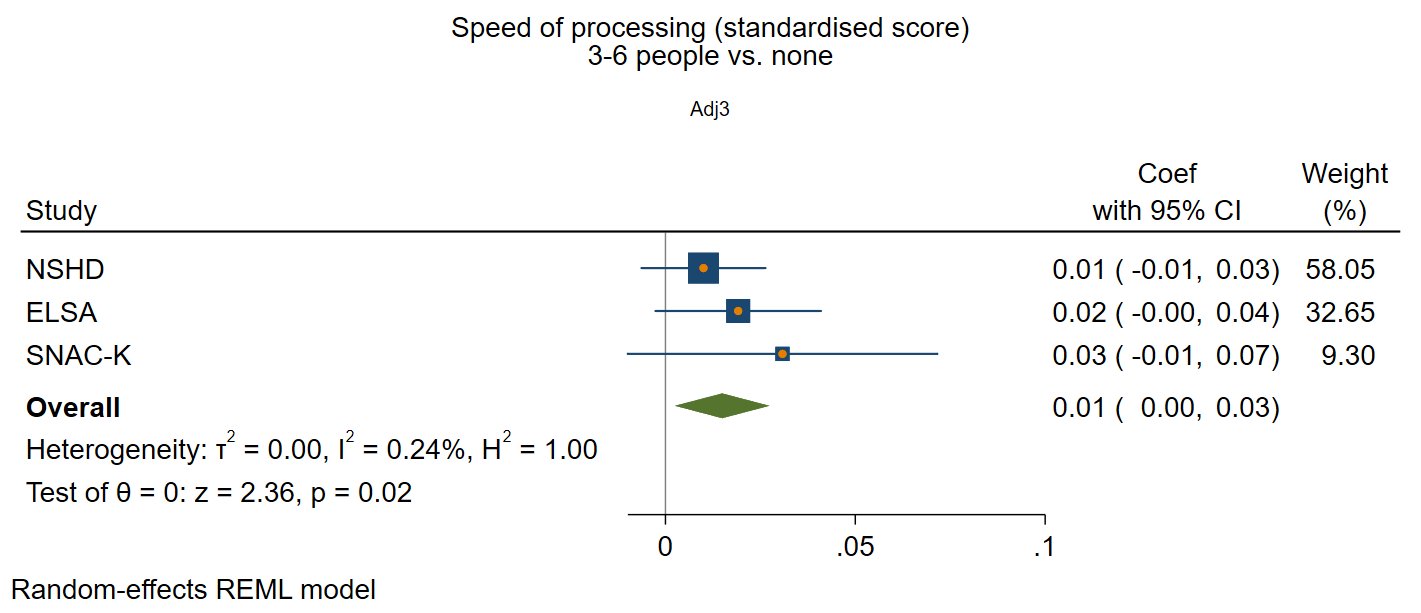

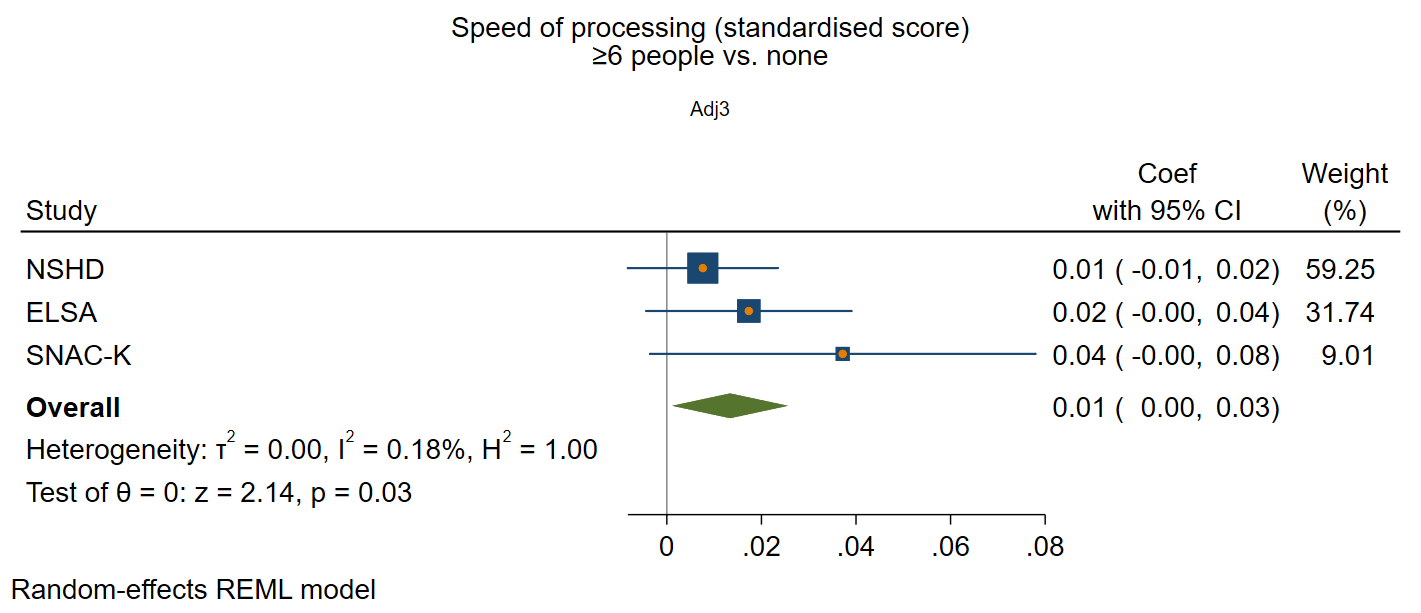

#### Frequency of contact

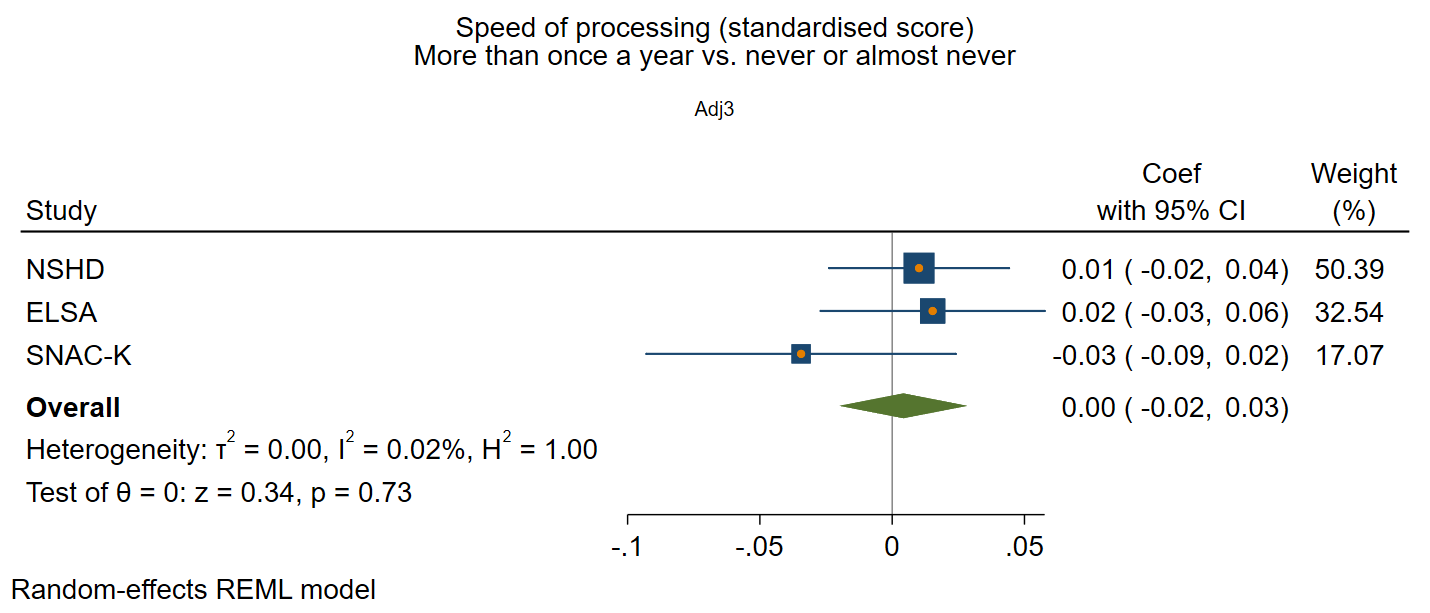

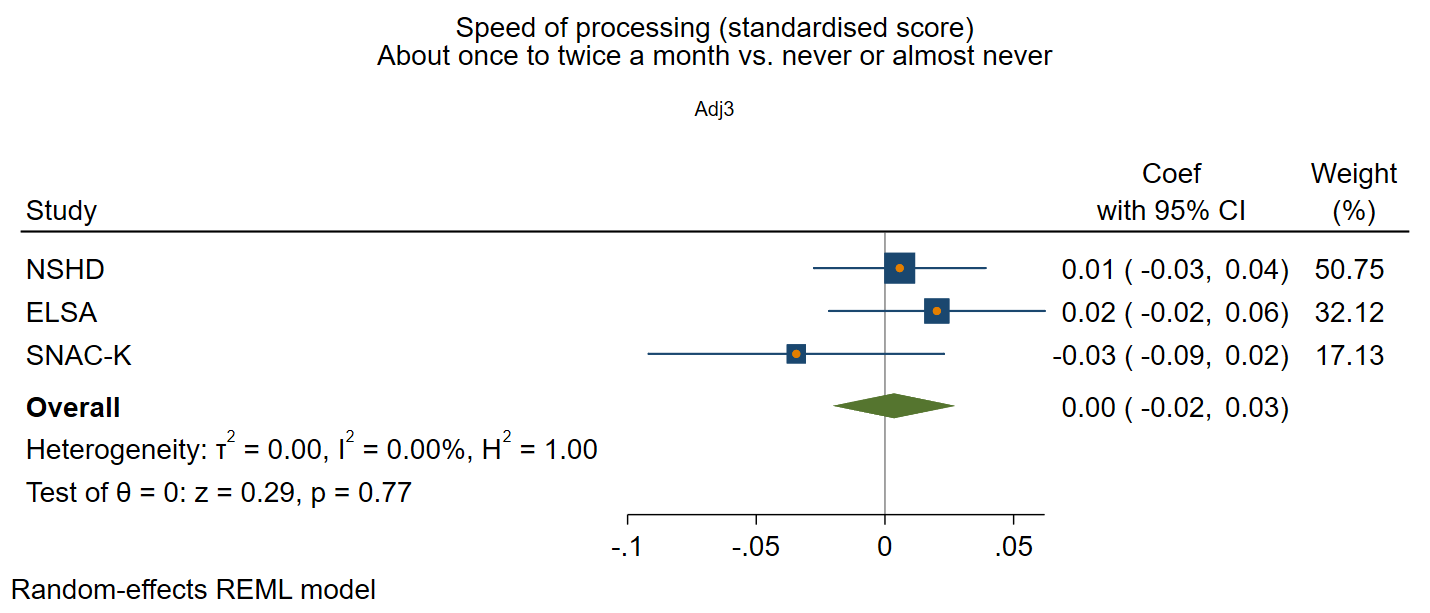

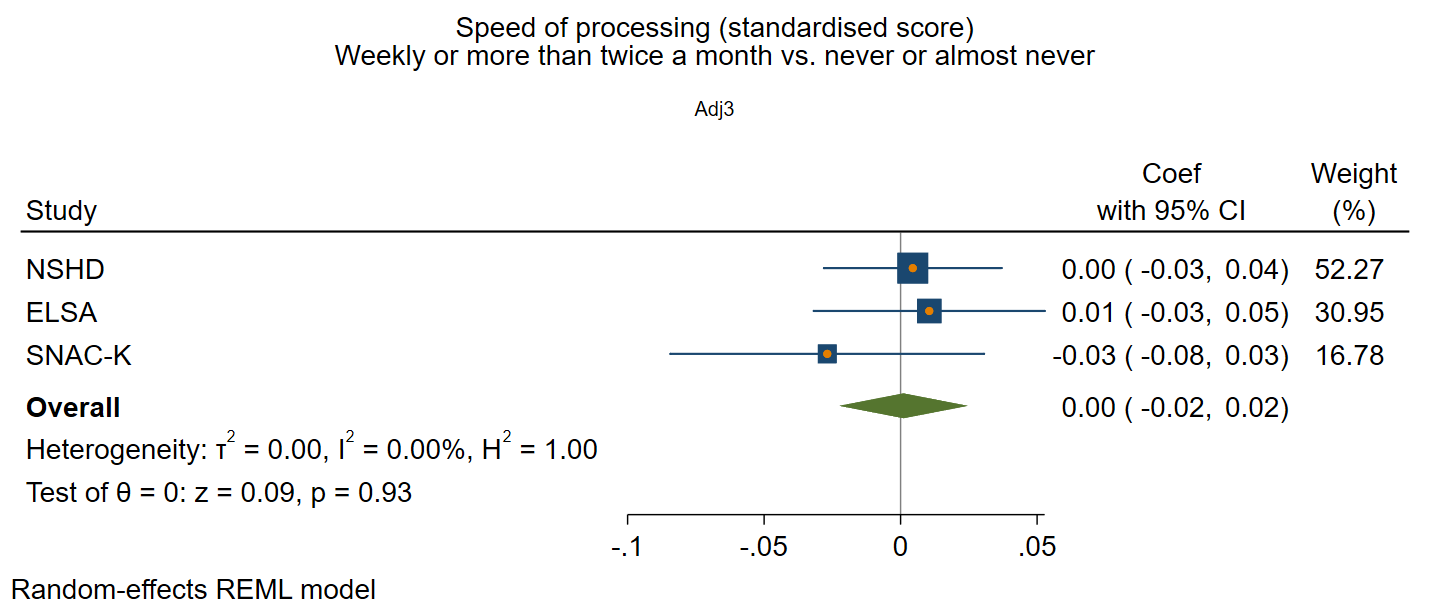

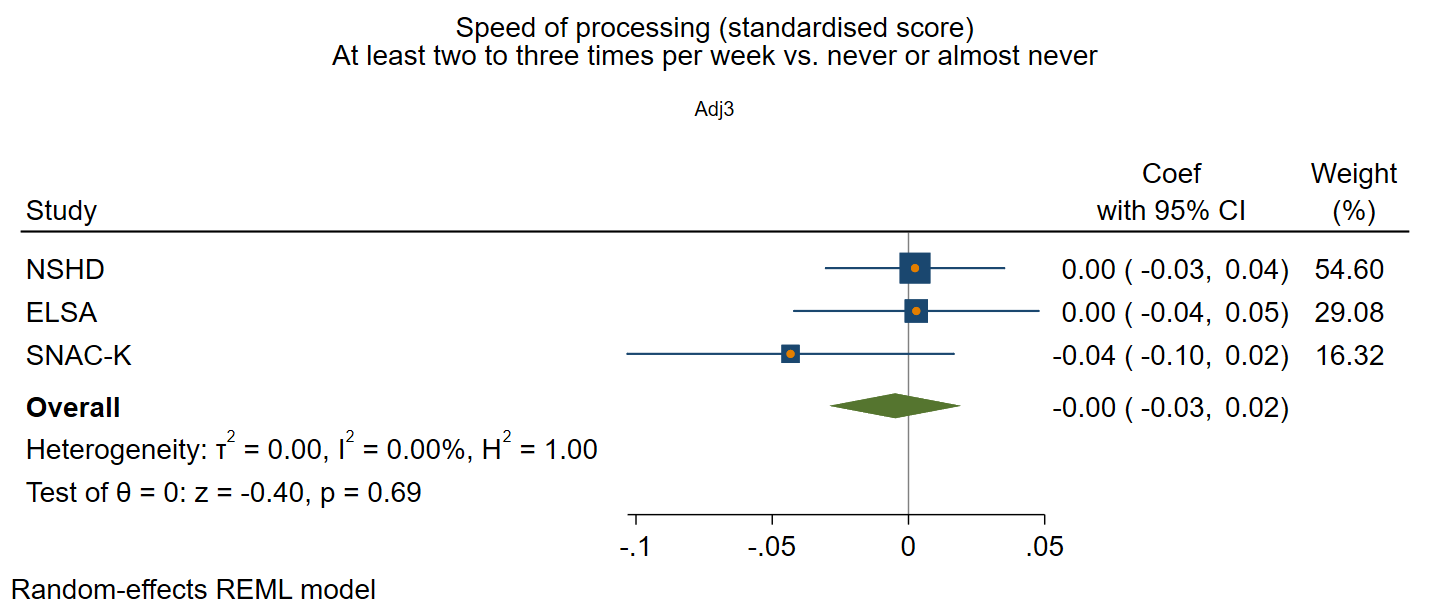

### Supplementary figure 7: Functional social health at baseline and rate of decline in processing speed

*Adjusted for sex and age at baseline, social class, education, IADL, and vascular-related health conditions.*

#### Participation in social activities

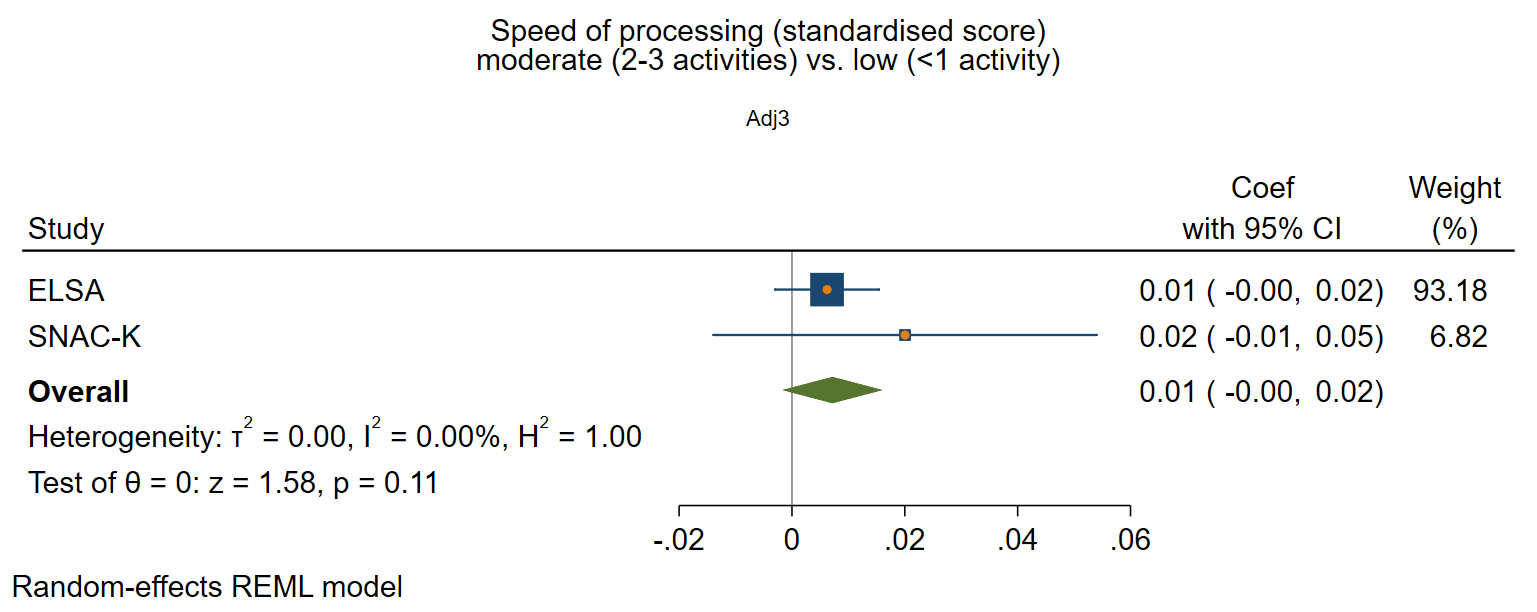

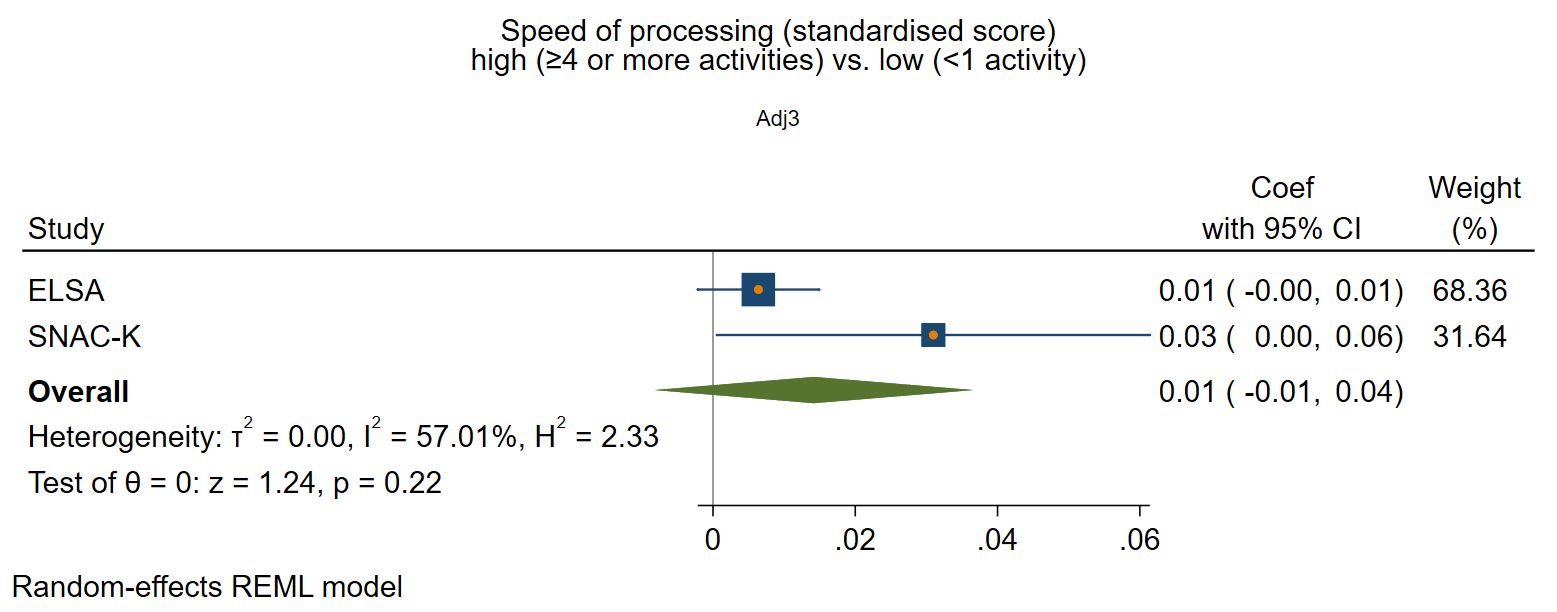

#### Perceived positive social support

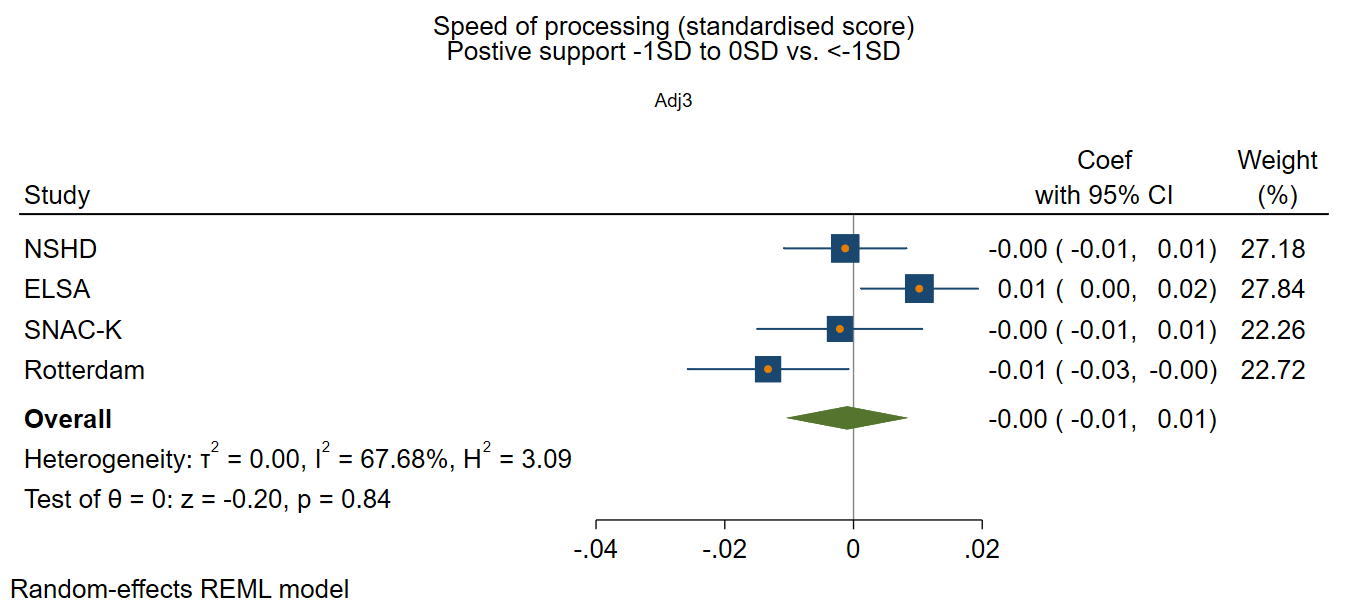

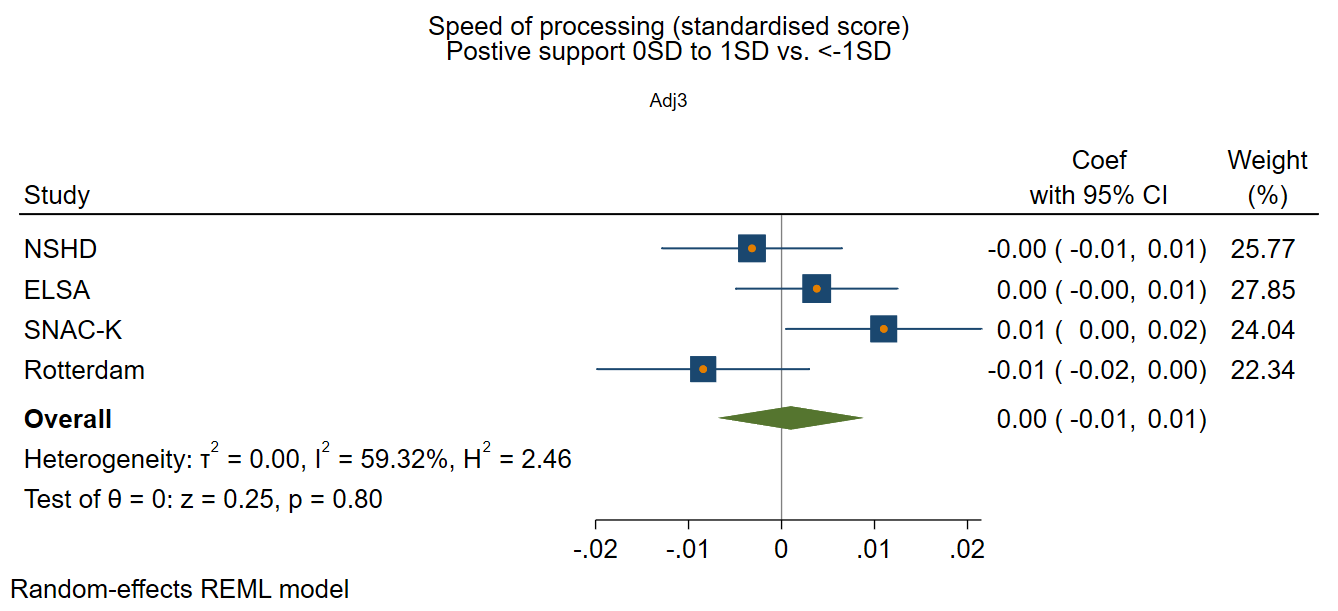

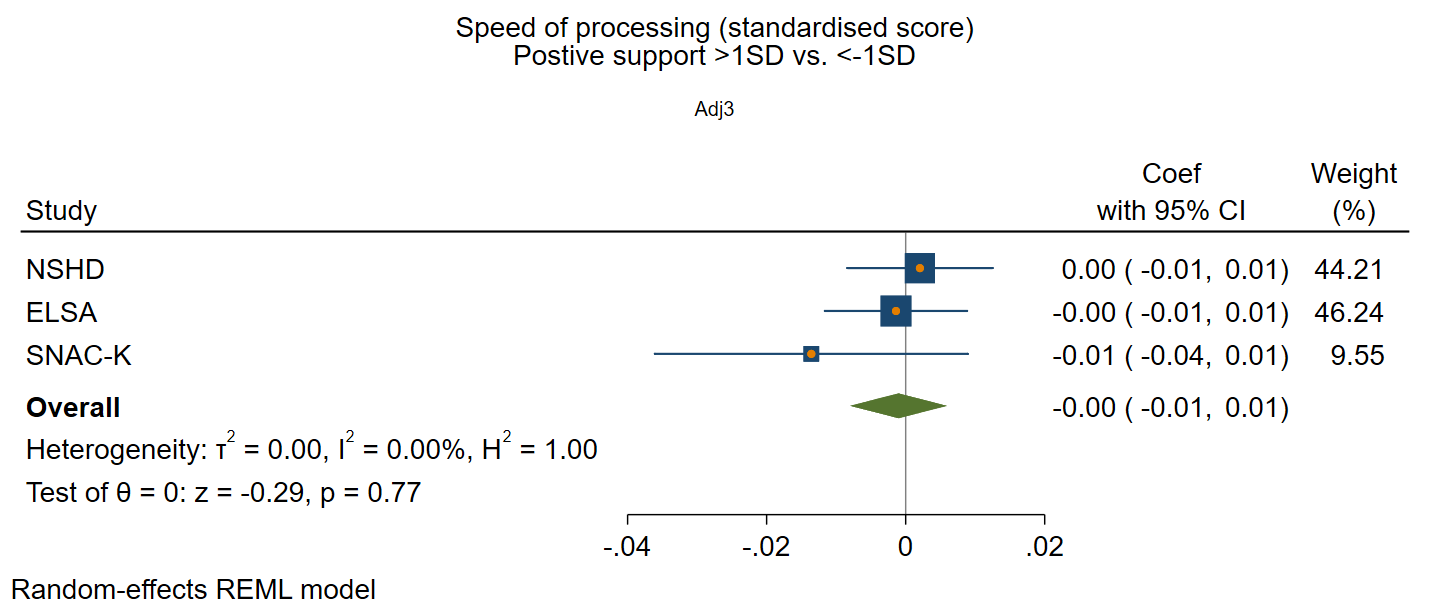

#### Perceived negative social support

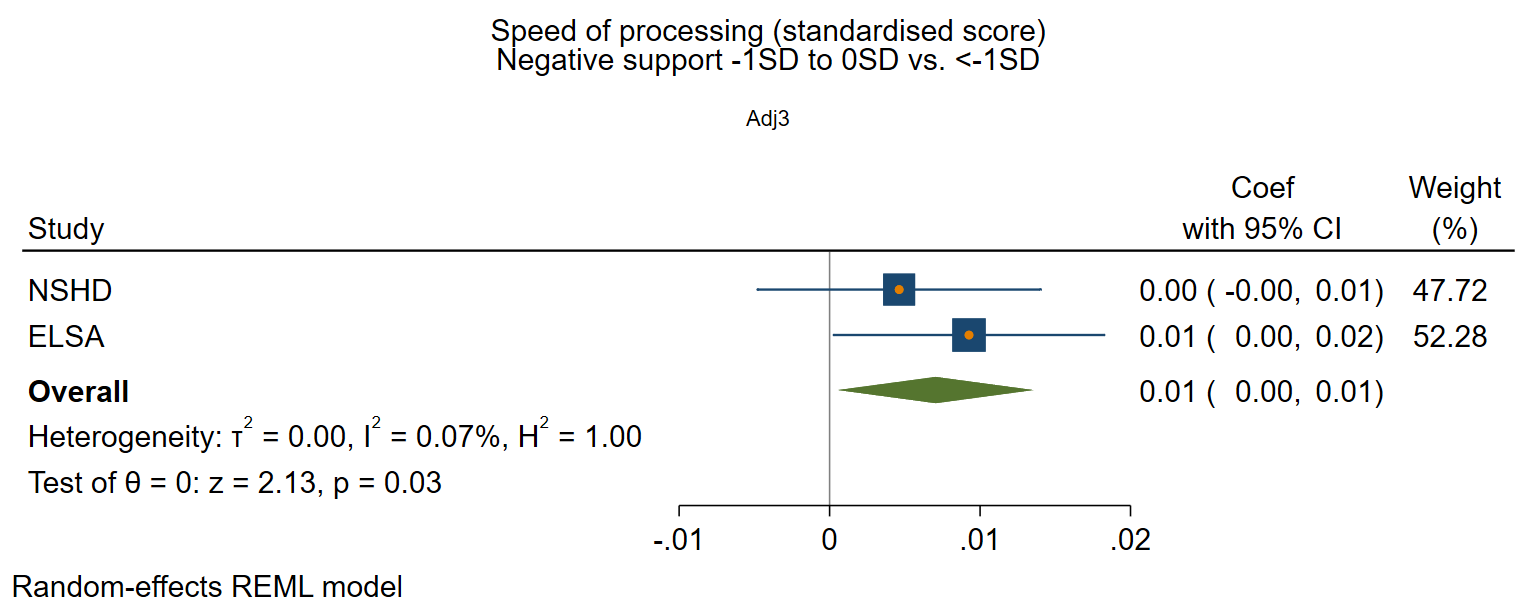

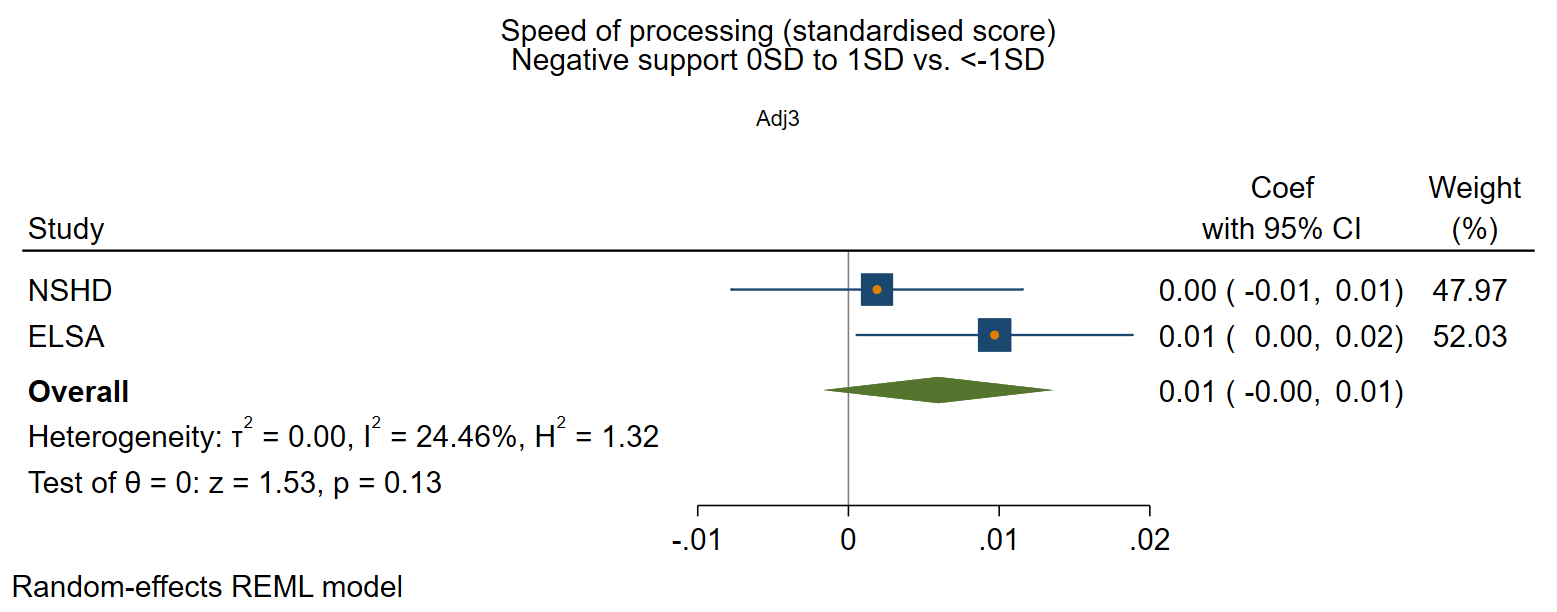

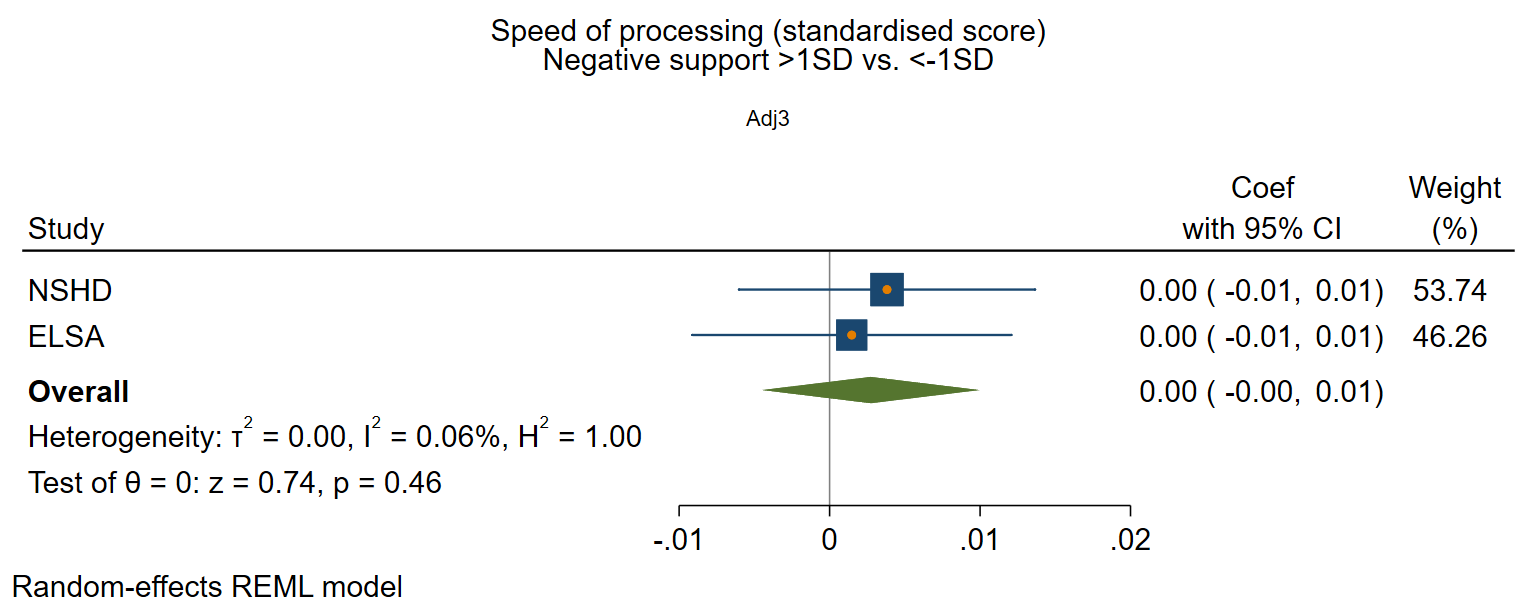

### Supplementary figure 8: Structural social health at baseline and rate of decline in global/composite cognition

*Adjusted for sex and age at baseline, social class, education, IADL, and vascular-related health conditions.*

#### Marital/cohabitation status

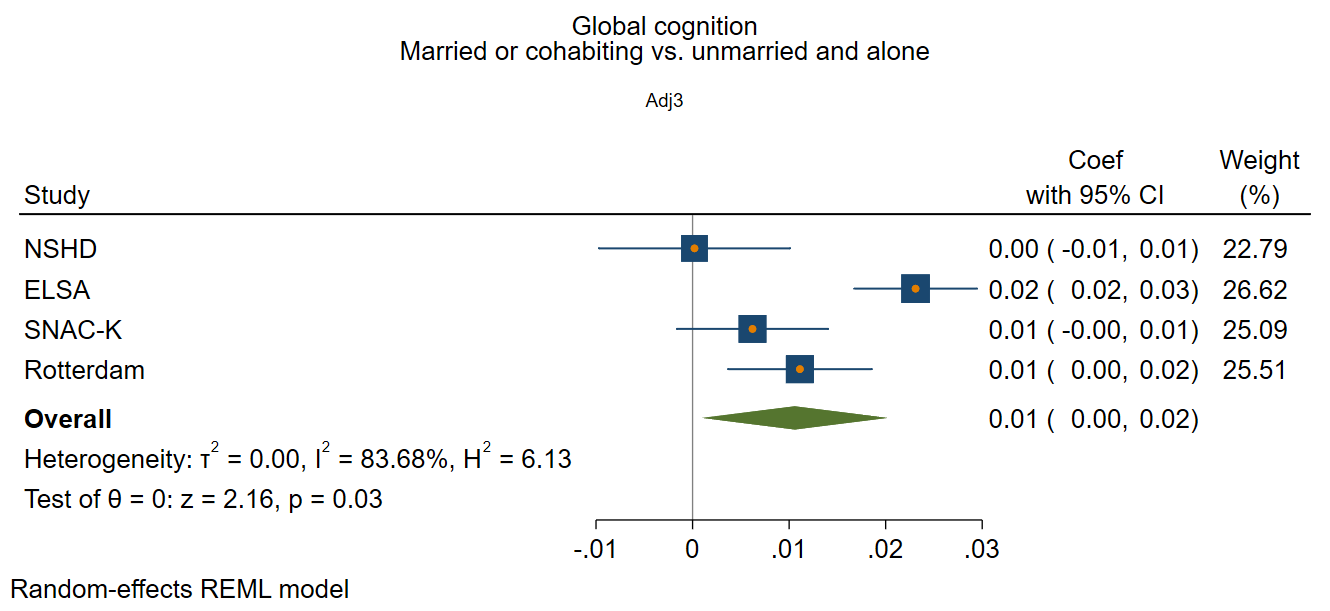

#### Network size

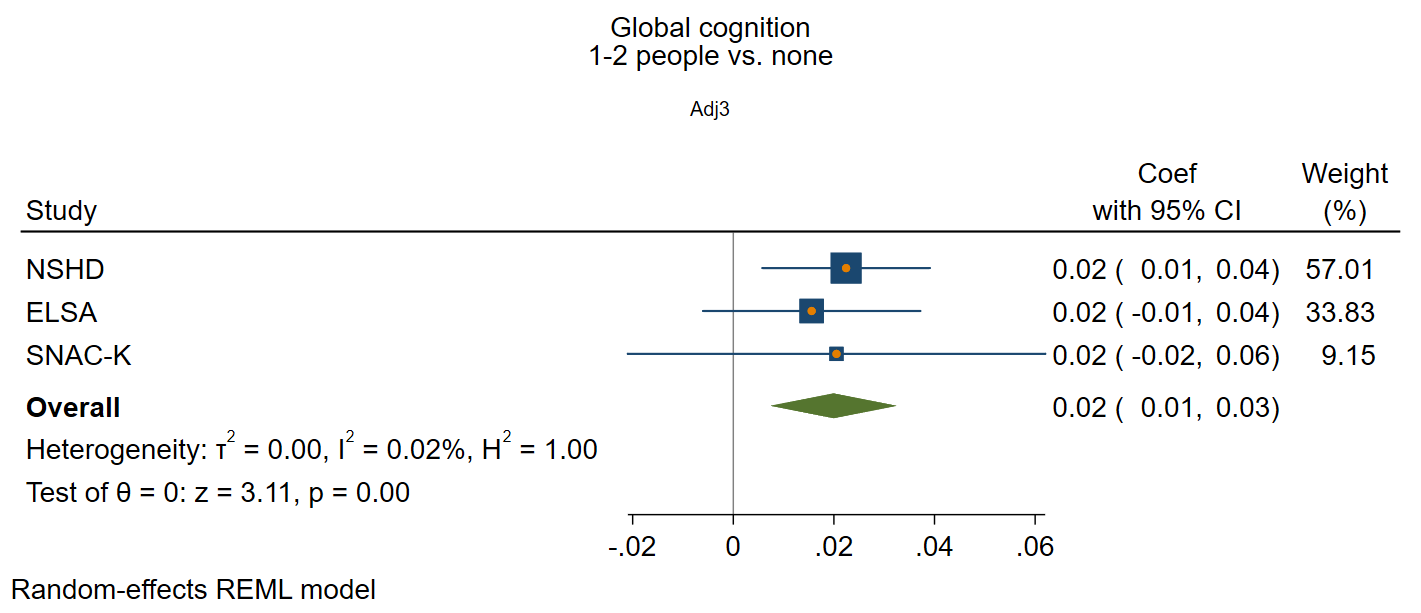

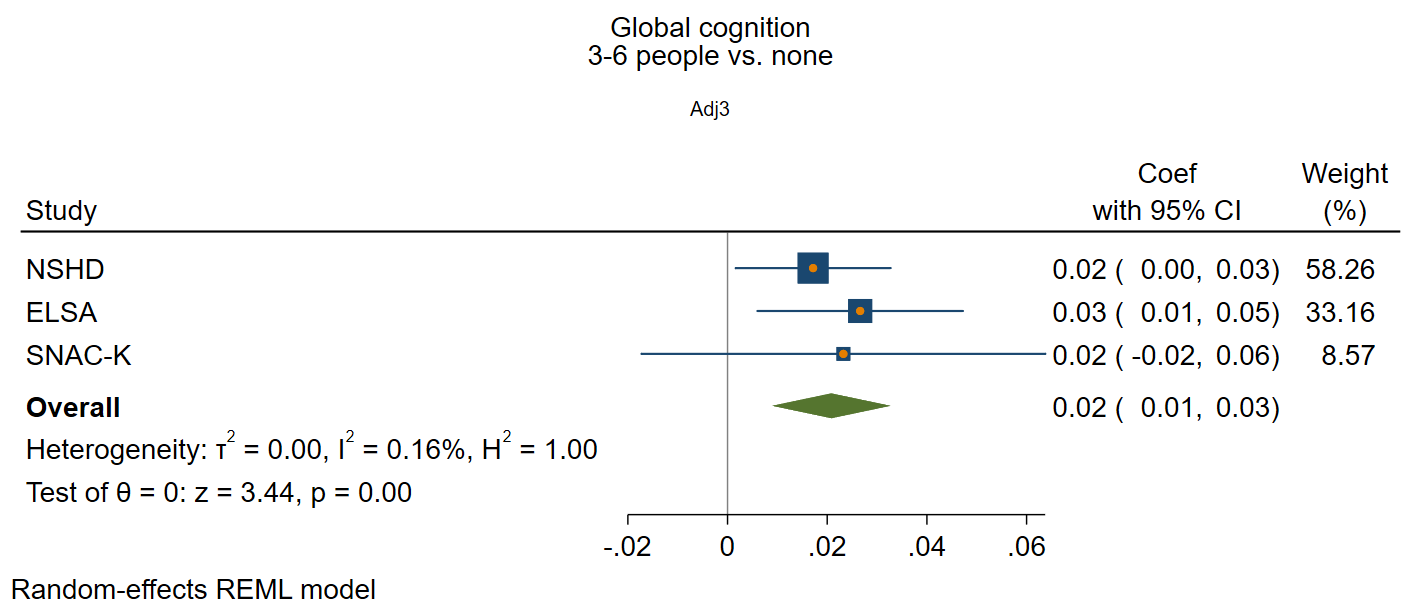

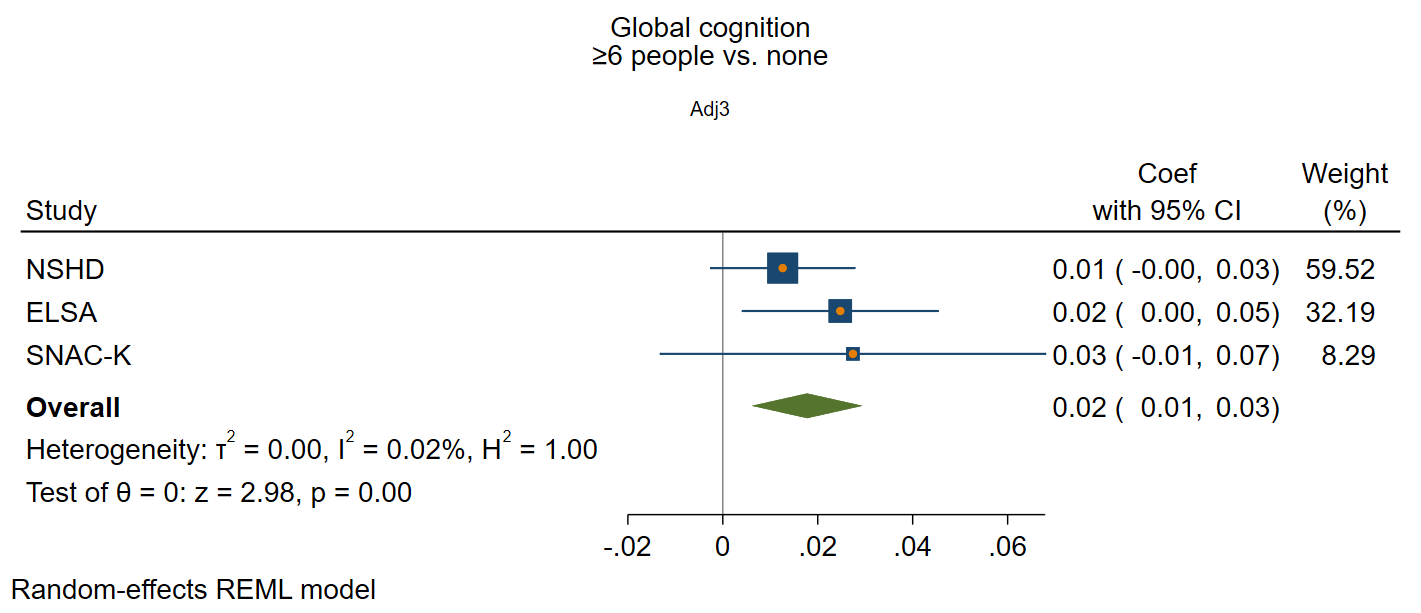

#### Frequency of contact

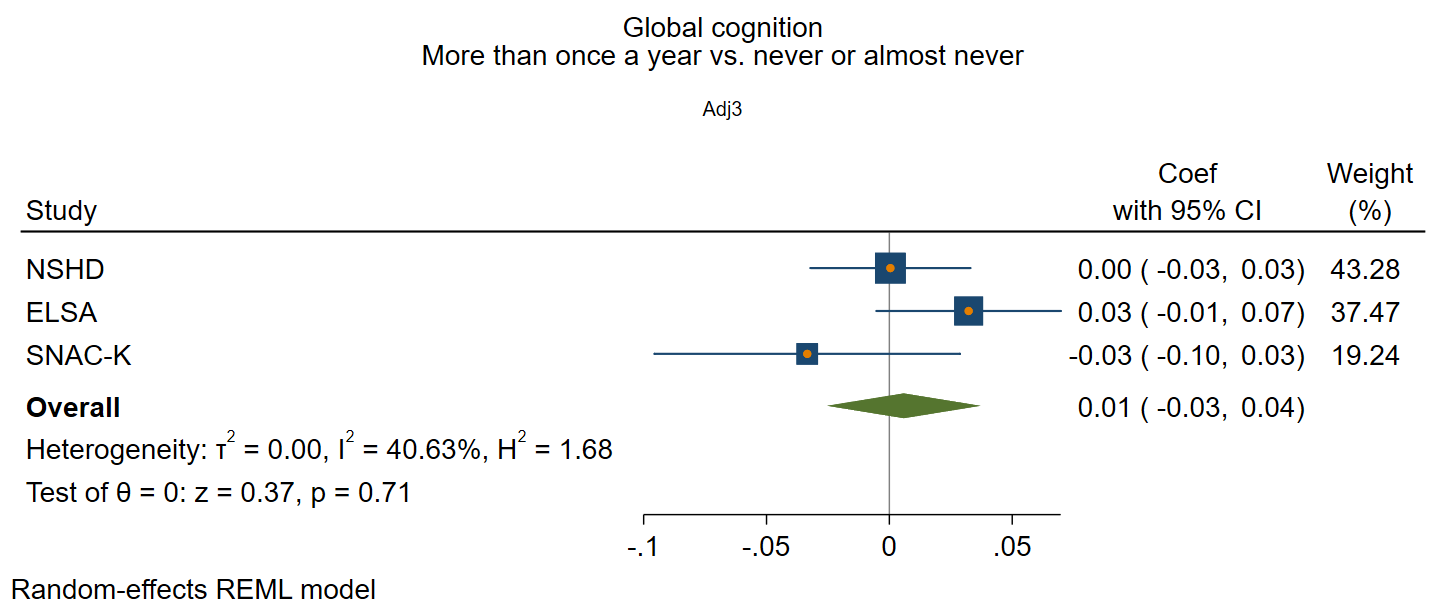

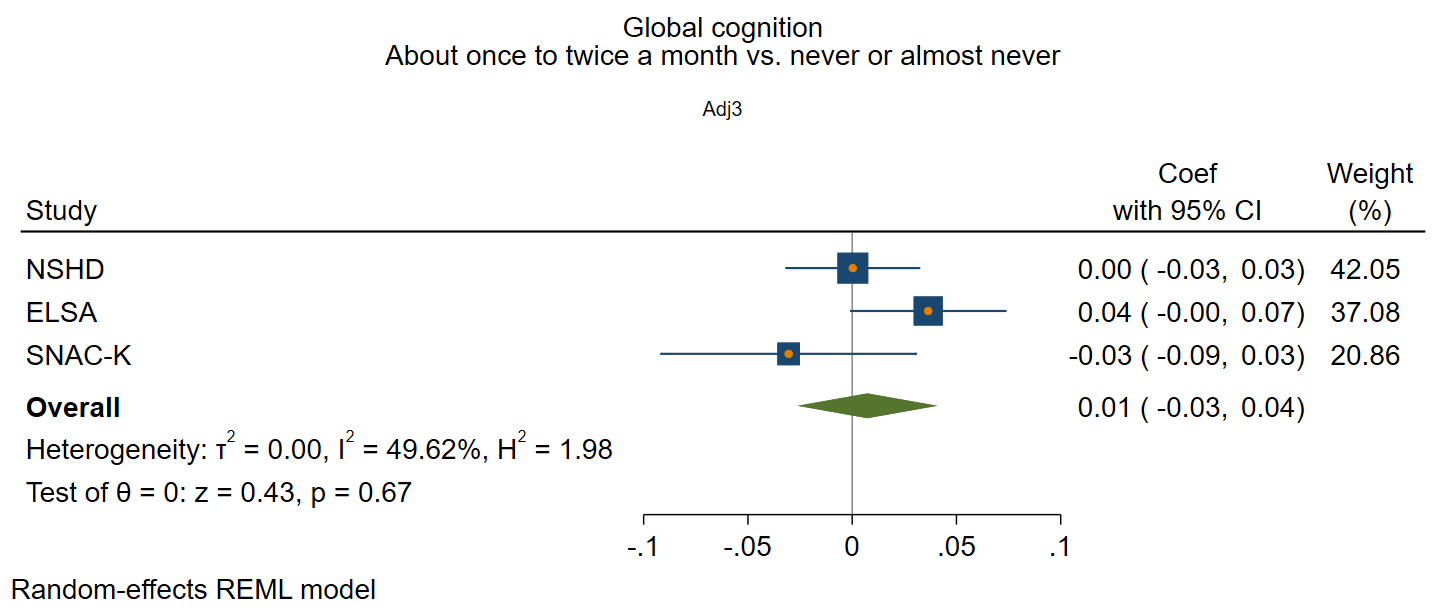

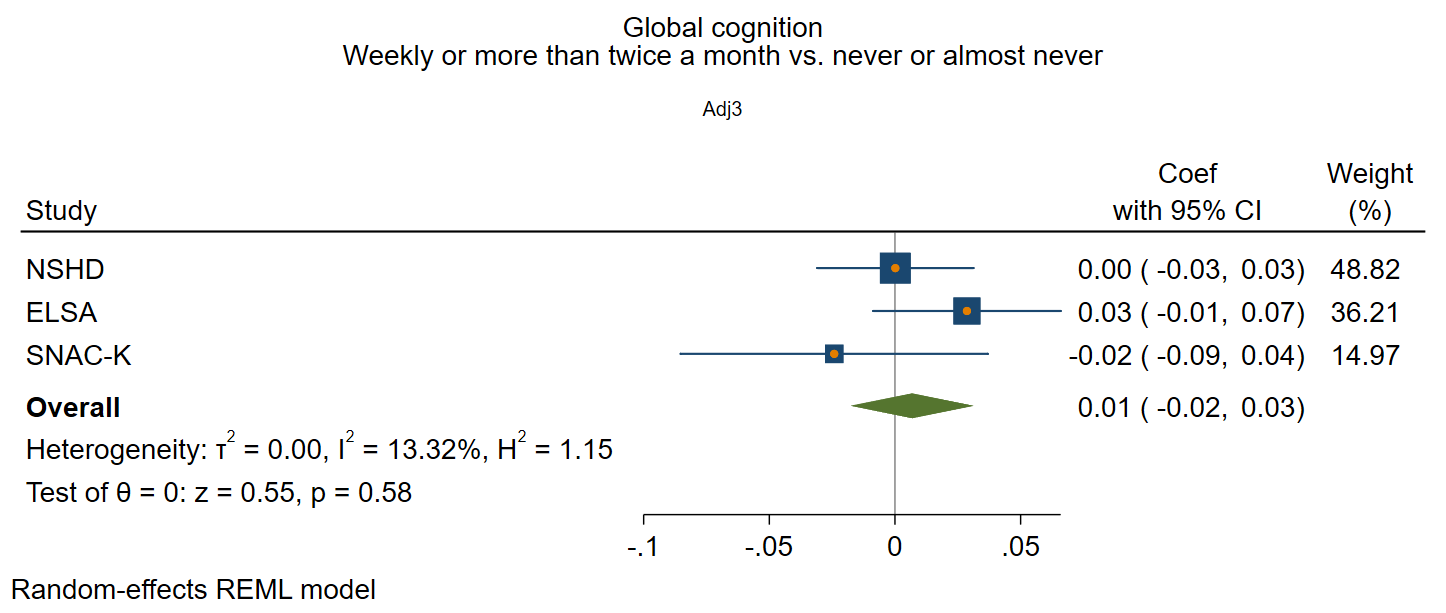

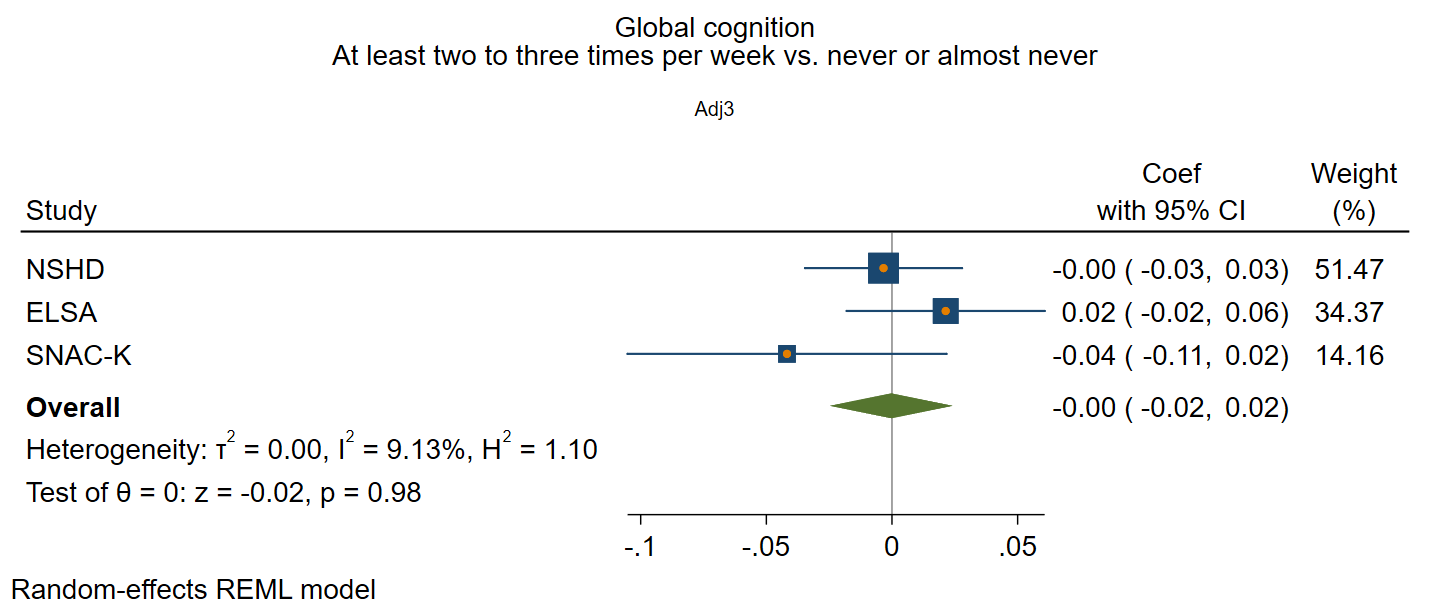

### Supplementary figure 9: Functional social health at baseline and rate of decline in global/composite cognition

*Adjusted for sex and age at baseline, social class, education, IADL, and vascular-related health conditions*

#### Participation in social activities

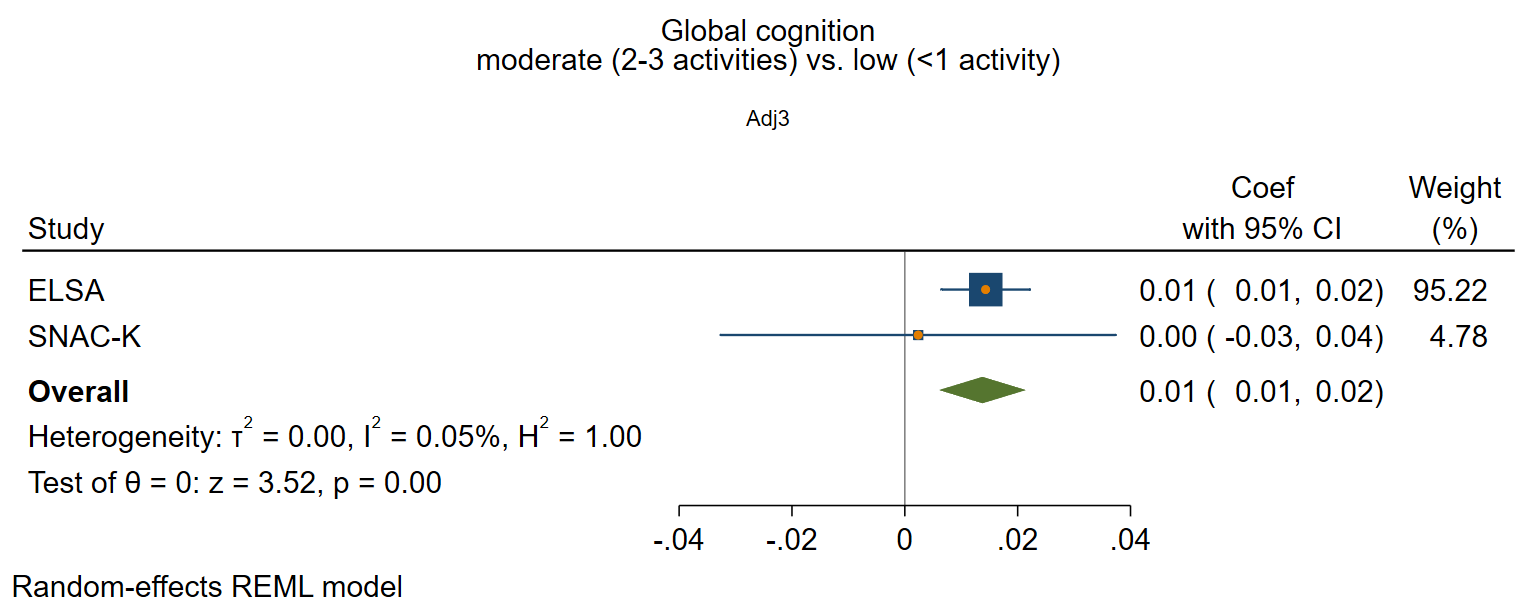

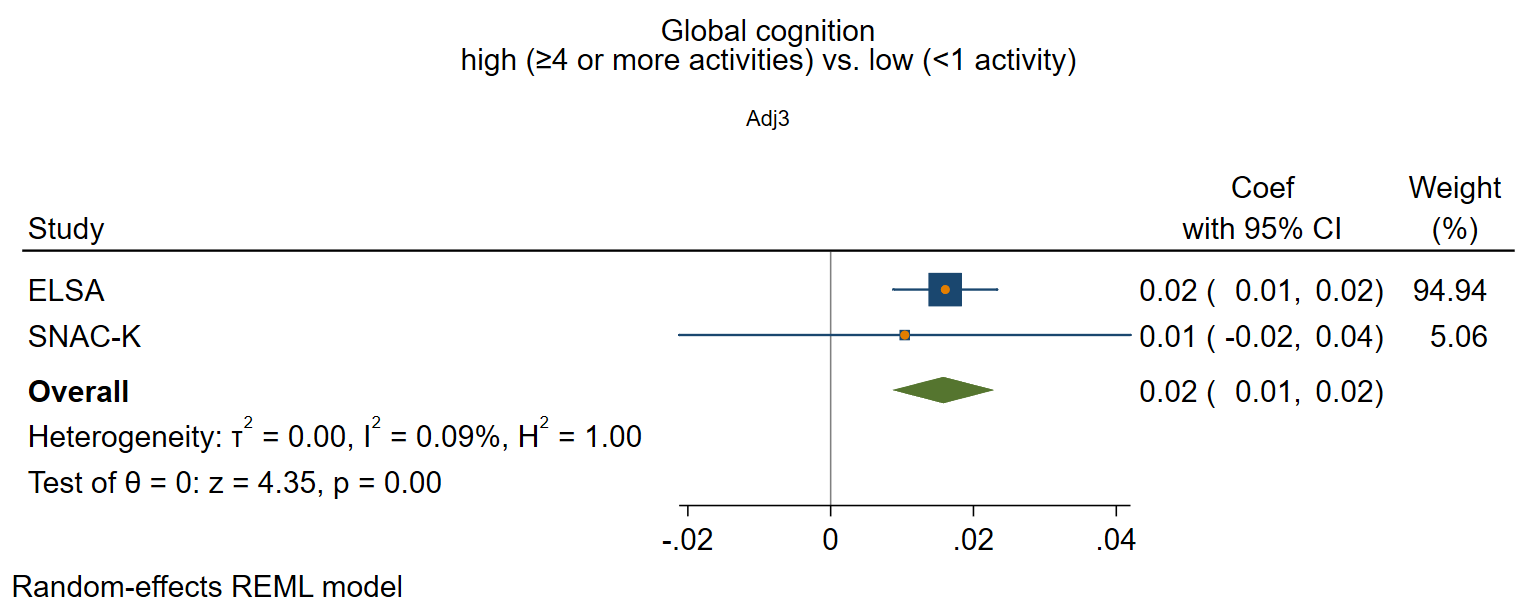

#### Perceived positive social support

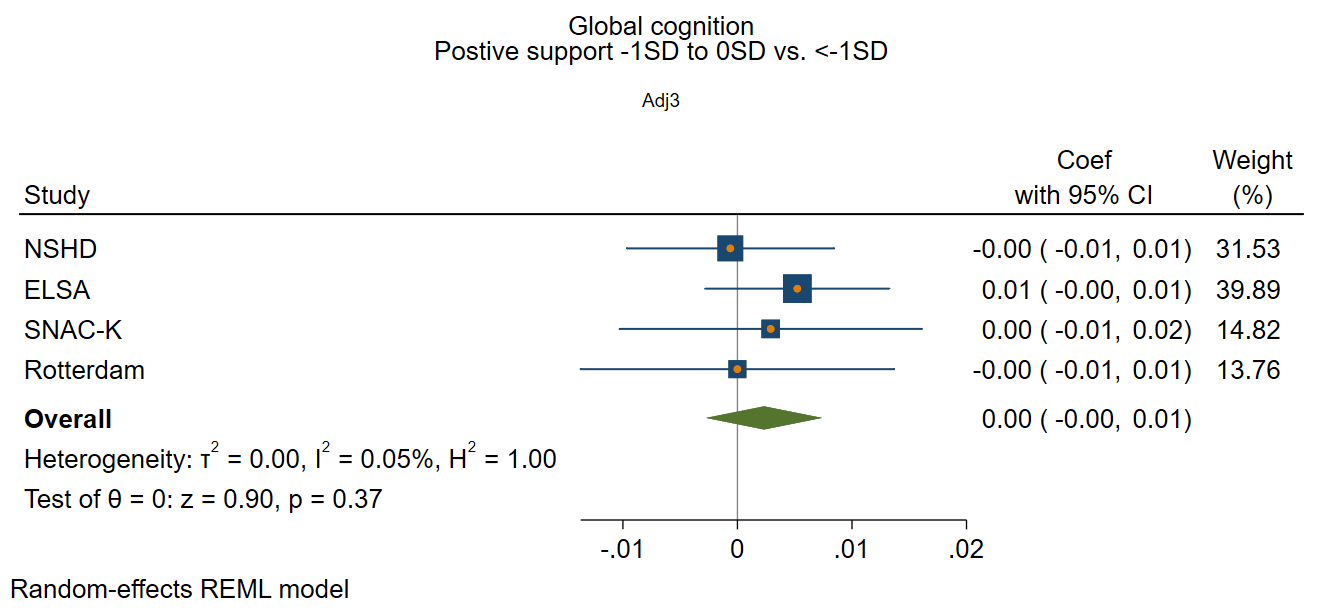

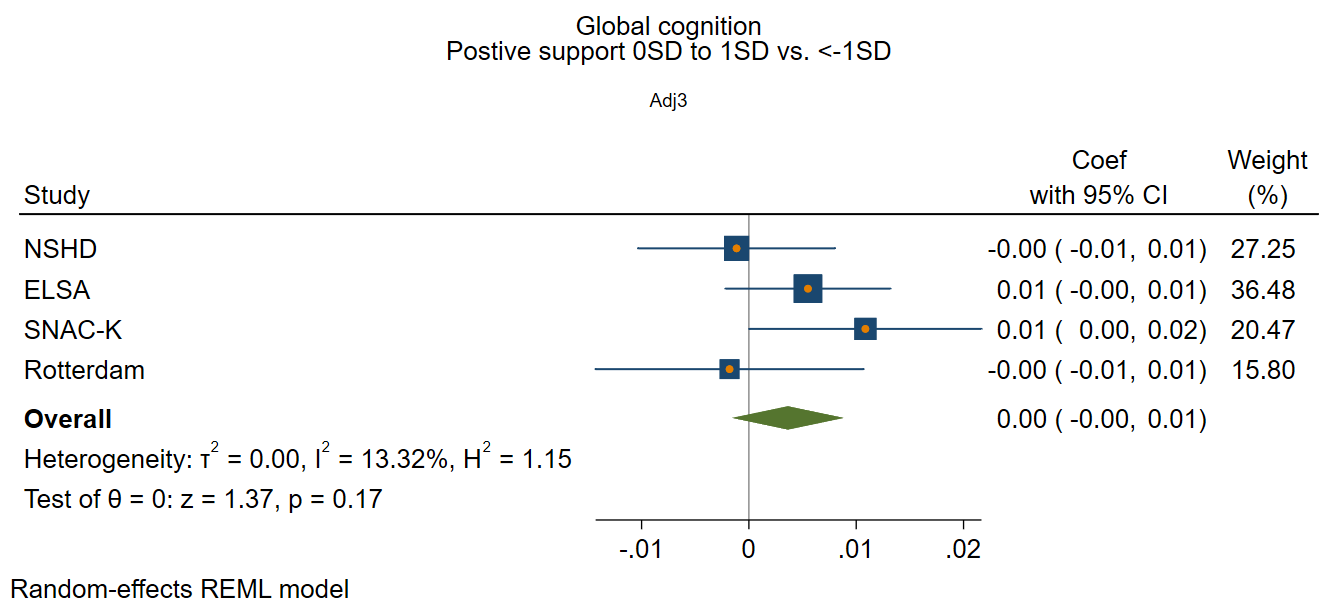

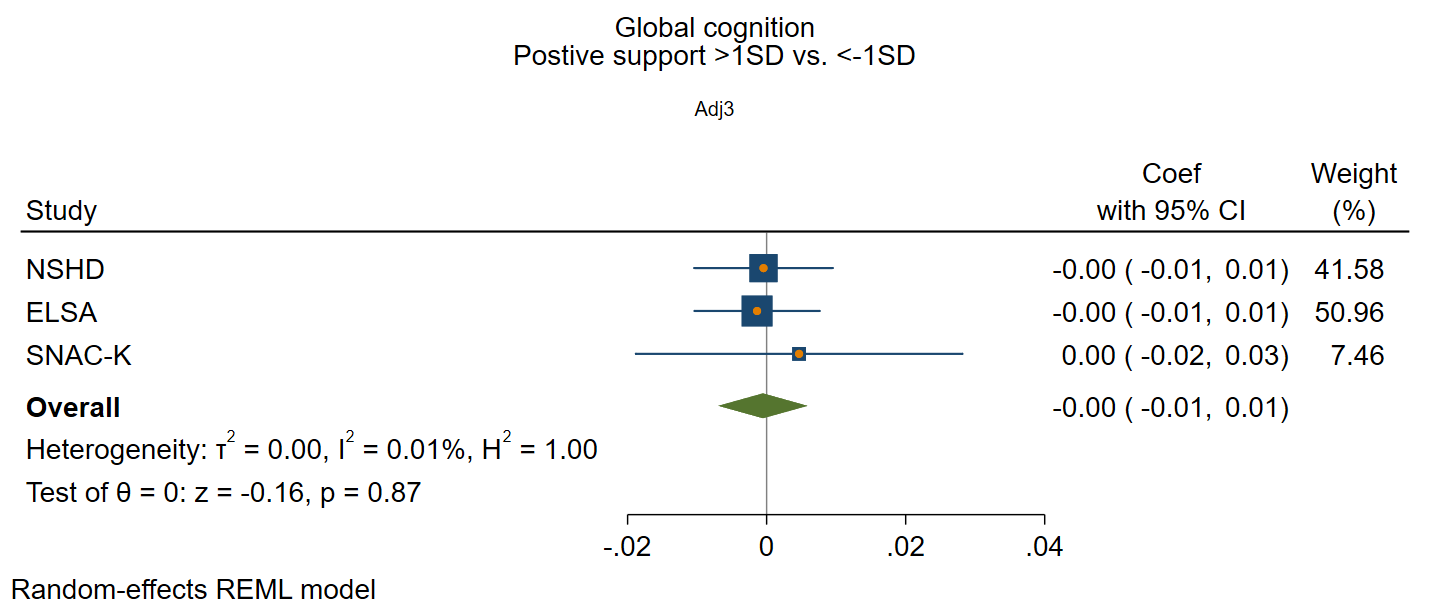

#### Perceived negative social support

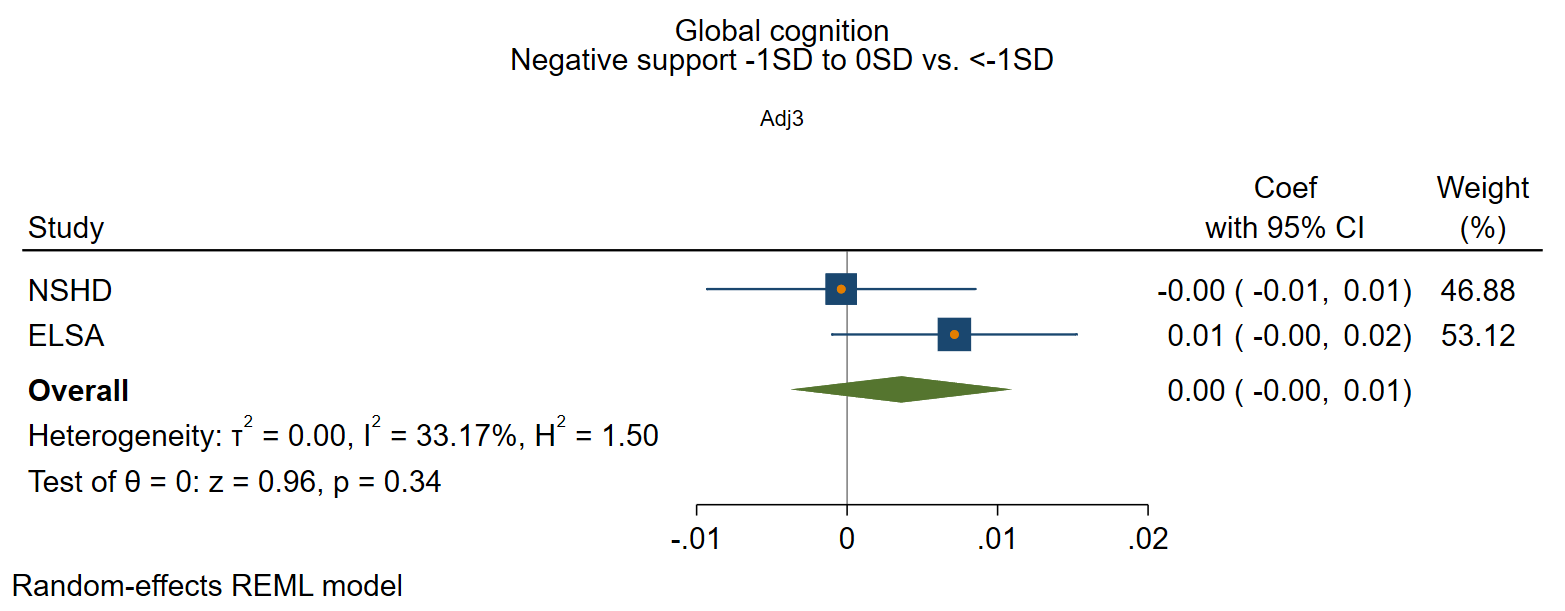

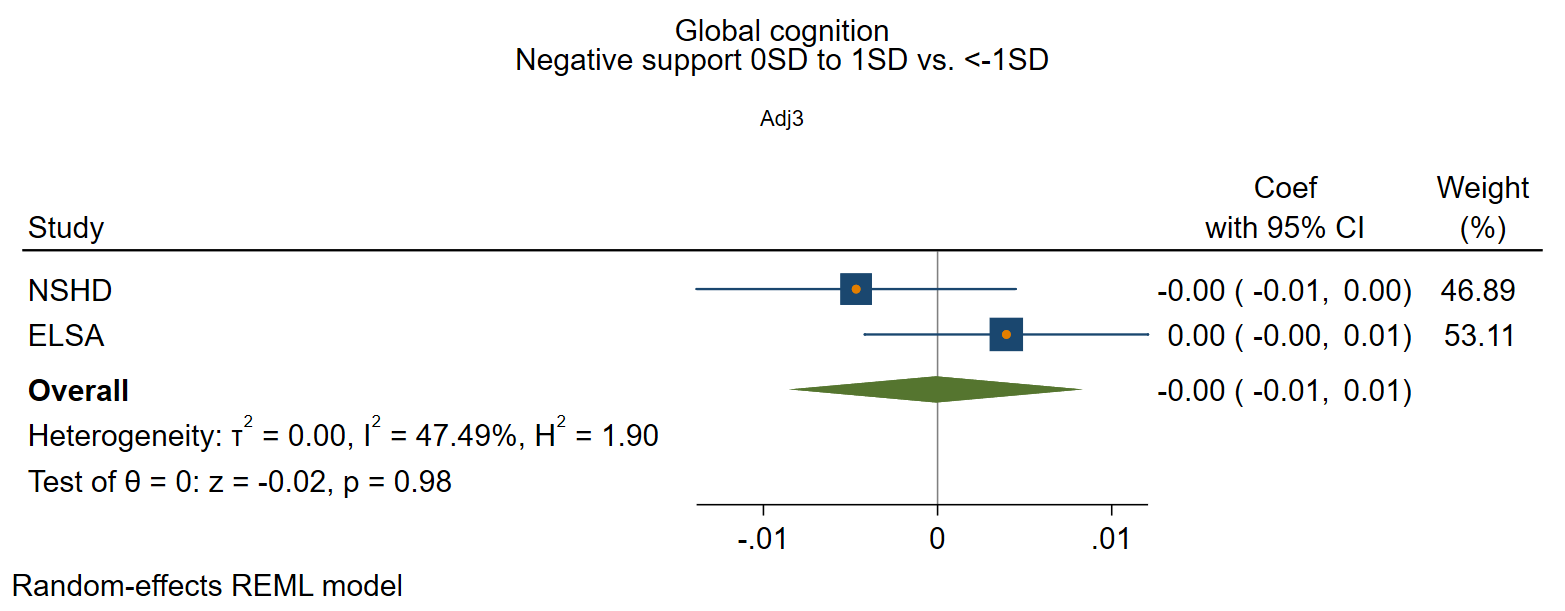

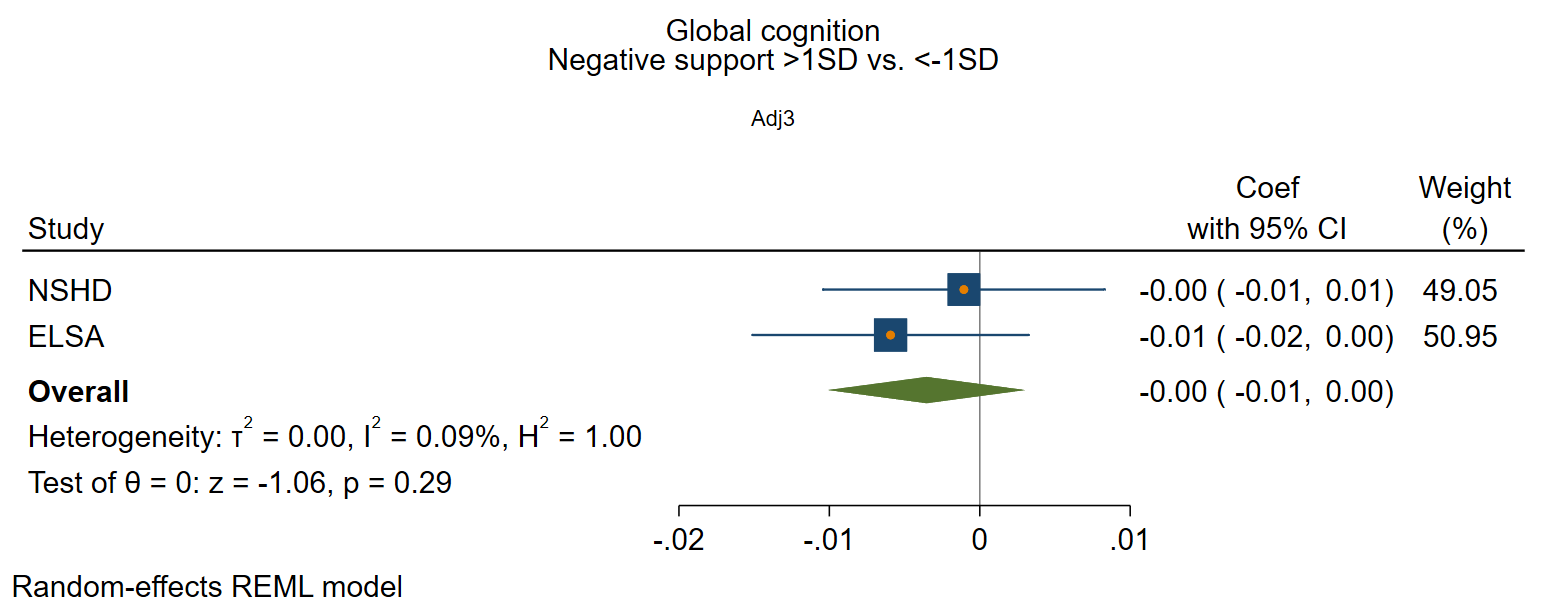
